# Dietary intake of plant bioactives among European adults

**DOI:** 10.64898/2026.04.14.26350848

**Authors:** Costanza Michelini, Federica Bergamo, Alice Rosi, Daniele Del Rio, Pedro Mena

**Author notes:** Corresponding author: Pedro Mena, Human Nutrition Unit, Department of Food and Drug, University of Parma, Via Volturno, 39, 43125 Parma, Italy.

## Abstract

This work explores the dietary intake of plant bioactives in the European adult population. The information available in the scientific literature is quite fragmented, with only partial knowledge of dietary bioactive intake and their health effects, and without harmonised figures across populations and phytochemical families. In this context, we comprehensively evaluated the intake of (poly)phenols, terpenoids, *N*-containing compounds, and miscellaneous phytochemicals in the European adult population, using public data from 26 countries reporting on 38,944 individuals. Further research was conducted to investigate the contributions of classes, subclasses, and individual compounds, as well as their relationships. Main food sources of each class and subclass of phytochemicals were also identified. Finally, variability in phytochemical intake across European countries was evaluated. This work significantly advances the current knowledge of plant bioactive intake and sets the stage for future research in nutrition and health fields.

## Introduction

The close interconnection between diet and health is widely documented by decades of scientific research, providing evidence-based knowledge on the pivotal role of dietary patterns in non-communicable diseases^1,2^. Indeed, the impact of diet on morbidity and mortality risk is superior to other factors, such as alcohol and tobacco^3^. Eating healthy, by consuming largely fruit and vegetables, whole grains, pulses, and other plant-based foods and drinks (such as coffee and tea), also results in a high intake of small bioactive molecules with beneficial effects against chronic diseases^4^. For instance, the regular consumption of (poly)phenol-rich foods (such as apples, tea, berries, and dark chocolate) and carotenoid-rich foods (such as carrots, tomatoes, and peppers) has been associated with a lower risk of cardiovascular disease, type 2 diabetes, and certain cancer^5–8^.

The chemical complexity of foods has hardly been recognised, as only a small portion of the over 26,000 molecules in food matrices have been studied: this leads to significant gaps in our overall understanding of our diet and its health effects. Moreover, the potential beneficial effects of these compounds on our health cannot be attributed to a single player or compound class but, instead, to the continuous interactions among phytochemicals, further complicating the current understanding^9^. Nevertheless, assessing the intake of dietary plant bioactives is particularly challenging. Most studies tend to focus on a limited number of phytochemicals or compound classes at a time^5,7,10^, while very few provide a broader-yet limited-molecular picture of the current diet^11–13^. Comparative studies considering phytochemical intake across multiple populations with different dietary habits are lacking. Considering the European population, the intake of (poly)phenols has been extensively assessed, as shown in the EPIC study^14^. On the contrary, a more selective and restricted approach is generally employed when estimating the intake of other phytochemicals, such as carotenoids, phytosterols, and glucosinolates^15–17^. This creates disparities in the global understanding of the plant bioactive intake through the diet. Furthermore, several uncertainties related to self-reported dietary assessment tools, such as food frequency questionnaires and dietary recalls, need to be considered^18^. The interplay of these factors ultimately leads to substantial variability in dietary phytochemical consumption across countries, posing additional challenges for a comprehensive understanding of our diet.

Within this framework, our aim was to comprehensively assess the intake of dietary phytochemicals in the European adult population, considering (poly)phenols, terpenoids, *N*-containing compounds, and miscellaneous phytochemicals as primary families of bioactive molecules. Food consumption data from the EFSA Comprehensive Food Consumption Database for 26 countries were used and linked to the Phytochemical Food Database (PhytoFooD)^19^, which provided bioactive composition data for plant-based foods. We investigated the phytochemical composition of Europeans’ diet, also providing information on the contribution of each compound class, and which foods and chemicals are the main drivers of the assessment. The variability in phytochemical intake across European countries was also evaluated, shedding light on the complex and multifaceted landscape of plant bioactive exposure.

## Results

### Intake of phytochemicals in Europe

The dietary exposure of European adults to plant bioactives was estimated considering food consumption data from 26 countries, matching the food items to the Phytochemical Food Database (PhytoFooD), which includes data on 1,410 foods and 1,067 compounds^19^. Figure 1 highlights the average intake of phytochemicals in the European population. Taken together, the average intake of dietary plant bioactives among Europeans amounted to 5.1 ± 1.1 g/day. Terpenoids, including tetra-, tri-, di-, sesqui- and monoterpenoids were the most consumed plant bioactive compounds from the diet, exceeding 3.7 g per day in one reported case (Fig. 1a). The (poly)phenol family, accounting for flavonoids, phenolic acids, tannins and miscellaneous (poly)phenols, and *N*-containing compounds, including alkaloids, glucosinolates & isothiocyanates and miscellaneous *N*-containing compounds, held the second and third positions, slightly exceeding 1 g/day each on average. Regarding major classes for each family, monoterpenoids represented 65% of the total terpenoids (∼1.3 g/day), followed by triterpenoids (27%) and sesquiterpenoids (5%). Phenolic acids (62%) and alkaloids (53%) were the main classes contributing to each family, averaging ∼800 and ∼600 mg/day, respectively. Other relevant (poly)phenols were tannins (18%) and flavonoids (16%). Finally, the miscellaneous phytochemical family accounted for the lowest average intake (approximately 700 mg/day), of which more than 600 mg/day was attributable to phytates. Detailed information on the average phytochemical intake in Europe is presented in Tables S1-S4.

**Fig. 1:**
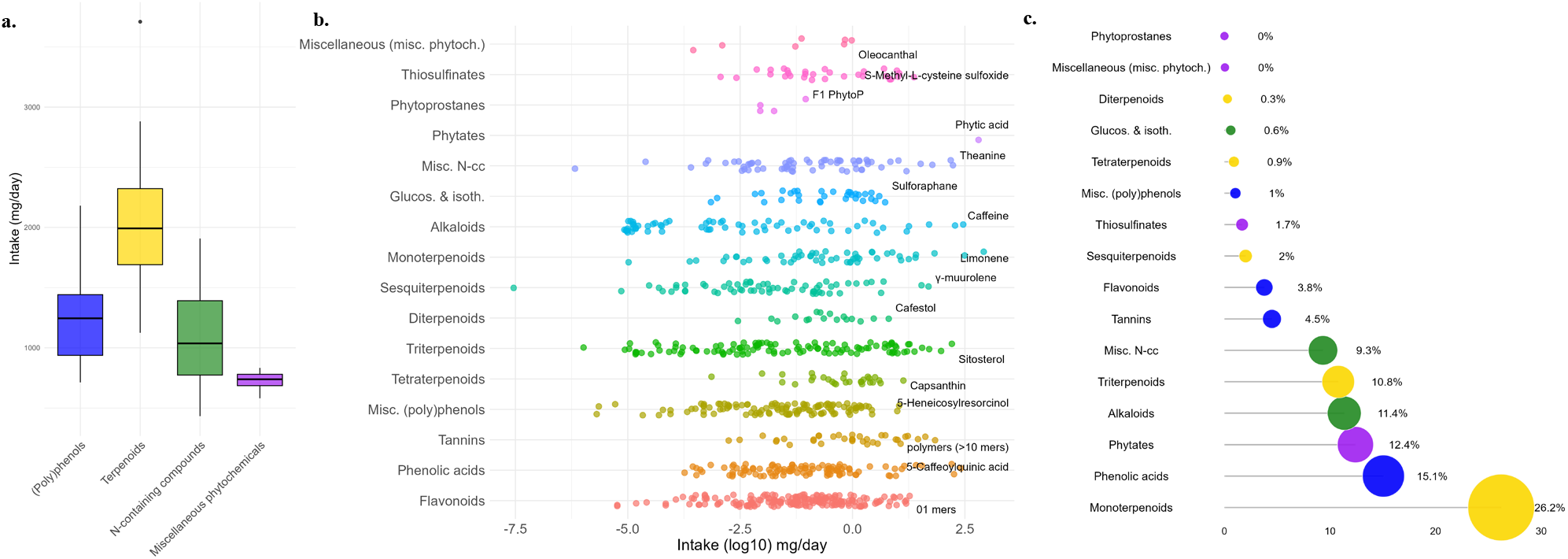
Average intake of phytochemicals in Europe. The average intake of bioactive compounds, divided into (poly)phenols, terpenoids, *N*-containing compounds, and miscellaneous phytochemicals, was evaluated in 26 European countries. These results were then grouped, to be representative of the European adult population. **a**. Distributions of phytochemical intakes in Europe considering average plant bioactive consumption of 26 countries. **b**. Most consumed plant bioactives (on average) for each phytochemical class in Europe; data is presented as a logarithmic scale. **c**. Contribution (%) of plant bioactive classes to the overall phytochemical intake.

We also identified top-contributing compounds for each phytochemical class (Fig. 1b): for instance, 5-caffeoylquinic acid, caffeine, sitosterol, and sulforaphane resulted as drivers of phenolic acid, alkaloid, triterpenoid, and glucosinolate & isothiocyanate intakes, respectively. While data for these plant bioactive classes were somewhat expected, this level of detail is frequently missing for lesser-studied phytochemical classes, such as thiosulfinates, phytoprostanes, mono-, sesqui- and diterpenoids. Here, we could evaluate that S-methyl-L-cysteine sulfoxide, F1-phytoprostane, limonene, γ-muurolene and cafestol were the most consumed compounds within their respective plant bioactive categories. It is worth noting that, in some cases, a distinct gap was observed between the leading bioactive and other chemicals of the same class, such as for *S*-methyl-L-cysteine sulfoxide, limonene, and capsanthin.

Primary food sources of phytochemicals for the European population were also determined. Figures 2a-d show the five main food contributors for each subclass of (poly)phenol, terpenoid, *N*-containing compound, and miscellaneous phytochemical families. We observed that, for some compound categories, a dominant food among all others emerged: for instance, red wine, bell peppers, potatoes, and fresh garlic were the principal suppliers of stilbenes, xanthophylls, steroidal alkaloids, and thiosulfinates. However, it is crucial to bear in mind that food commodities are composed of intricate networks of hundreds of molecules with diverse structures and bioactivities. This complexity was clearly evident in some foods that fall into multiple phytochemical classes, such as coffee, potatoes, carrots, and olive oil. On the contrary, some products can be considered exclusive sources of specific phytochemicals: this is particularly true for onions and garlic for thiosulfinates, and broccoli for isothiocyanates.

**Fig. 2:**
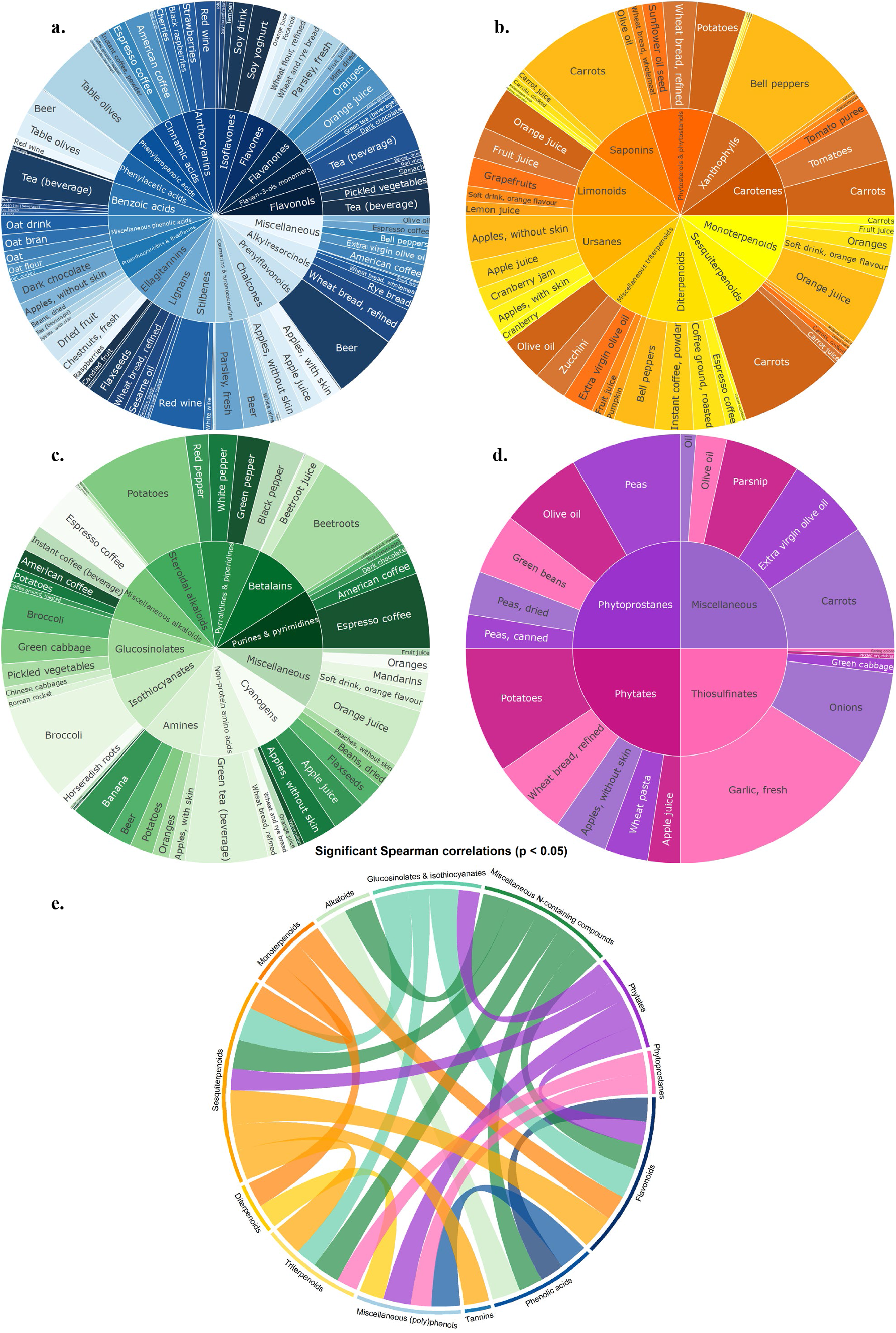
Associations between phytochemical classes and subclasses and their main food sources. Foods are complex matrixes, where hundreds of structurally different bioactive molecules are closely interconnected. Figures **a, b, c**, and **d** rank the five principal food sources for each bioactive subclass of (poly)phenol, terpenoid, *N*-containing compound, and miscellaneous phytochemical families, respectively. **e**, Significant Spearman’s correlations (p-value < 0.05) among phytochemical classes.

To further explore the relationships among different phytochemical classes, we performed Spearman’s correlations at the plant bioactive class level (p-value < 0.05). Two main patterns were observed: the first one-the most predictable-was composed of classes of chemicals belonging to the same family that are correlated, such as those belonging to the terpenoid family. The second type comprises phytochemical classes that, although members of different families, exhibit statistically significant correlations. For instance, the miscellaneous *N*-containing compound class (from the *N*-containing compound family) is strongly associated with sesqui- and triterpenoids (from the terpenoid family). This phenomenon can be briefly explained by the presence of common food sources, such as apples and citrus fruits, that simultaneously provide chemically and structurally diverse plant bioactives.

### Phytochemical intake variability within European countries

As part of the dietary intake assessment, a moderate variability in food consumption data across Europe needs to be accounted for. Therefore, we investigated the effects of differences in food consumption across European countries on daily phytochemical intake. Figure 3 shows the phytochemical intake distributions of total (poly)phenols, terpenoids, *N*-containing compounds, and miscellaneous phytochemicals. A pronounced variation was observed due to quantitative differences in food consumption.

**Fig. 3:**
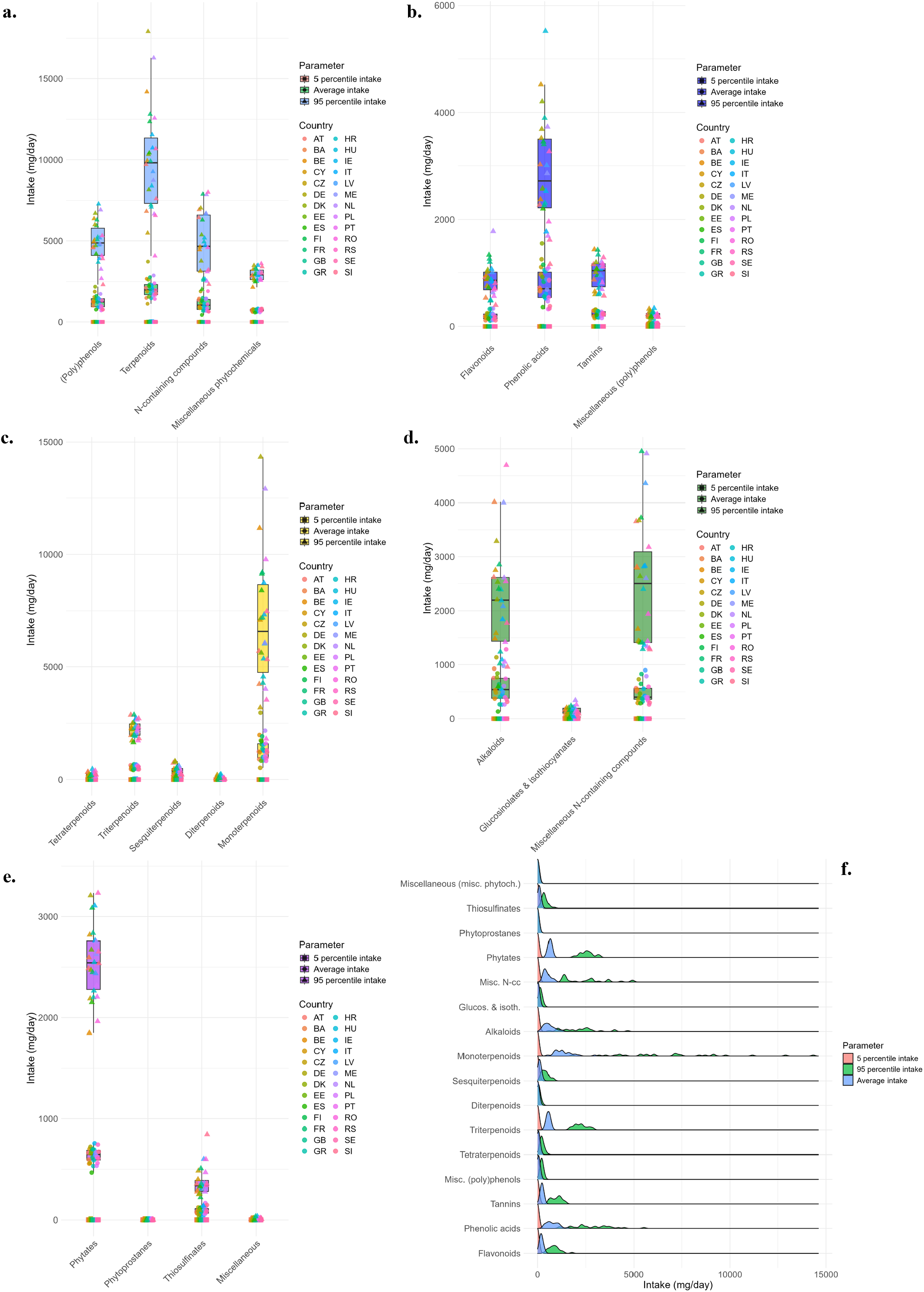
Phtytochemical intake distributions in Europe and European countries. Dietary habits represent powerful drivers of bioactive intake assessment. We investigated the phytochemical exposure differences, considering food consumption data of the 5^th^ percentile, the average, and the 95^th^ percentile of each European country. **a**. Phytochemical intake distributions of bioactive families among European countries. Figure **b, c, d** and **e** display the intake distributions among European countries of different classes of compounds, divided into (poly)phenols, terpenoids, *N*-containing compounds, and miscellaneous phytochemicals, respectively. **f**. Intake distributions of plant bioactive classes in Europe considering food consumption data of the 5^th^ percentile, the average, and the 95^th^ percentile of the population.

Terpenoids (Fig. 3c) were the bioactive family with the widest distribution, even when focusing on the same parameter. For instance, considering the 95^th^ percentile, Romania showed the lowest intake of terpenoids (4.1 g/day), while Germany had the highest, almost reaching 18 g/day. This is principally due to the elevated consumption of terpenoid-rich foods, such as citrus juices and multigrain breads. Nevertheless, these high values, beyond the extreme behaviour of some individuals, might be related to the lack of more accurate data, especially regarding the type of citrus juices consumed by these populations and the actual monoterpenoid concentrations within them. A similar pattern was observed in other phytochemical classes, such as alkaloids, miscellaneous N-containing compounds, and phytates (Fig. 3d-e). Informative data on the intake of different phytochemical families and classes for each country are provided in Table S5. These extended fluctuations appeared consistently in many cases (Fig. 3f and Fig. S1), highlighting the non-normal feature of plant bioactive intake distributions in the European population: for instance, in Fig. 3f, the monoterpenoid distribution of the 95^th^ percentile of the population visibly displays multiple peaks, nearly allowing us to identify the intake of individual countries. This peculiarity is particularly relevant when studying bioactive compounds that may have different implications for human health depending on dietary intake, such as caffeine. Indeed, our results showed that the average intake of caffeine in the European population is around 300 mg/day. This value is in line with EFSA recommendations not to exceed the safe threshold of 400 mg/day for caffeine intake^20^. However, looking at the caffeine consumption of the 95^th^ percentile of the population, it was observed that this safe limit was exceeded by 96% of the European countries considered in this assessment, particularly by Bosnia and Herzegovina, Montenegro, and Serbia (Fig. 4). Detailed data on country-specific caffeine intake are reported in Table S6.

**Fig. 4:**
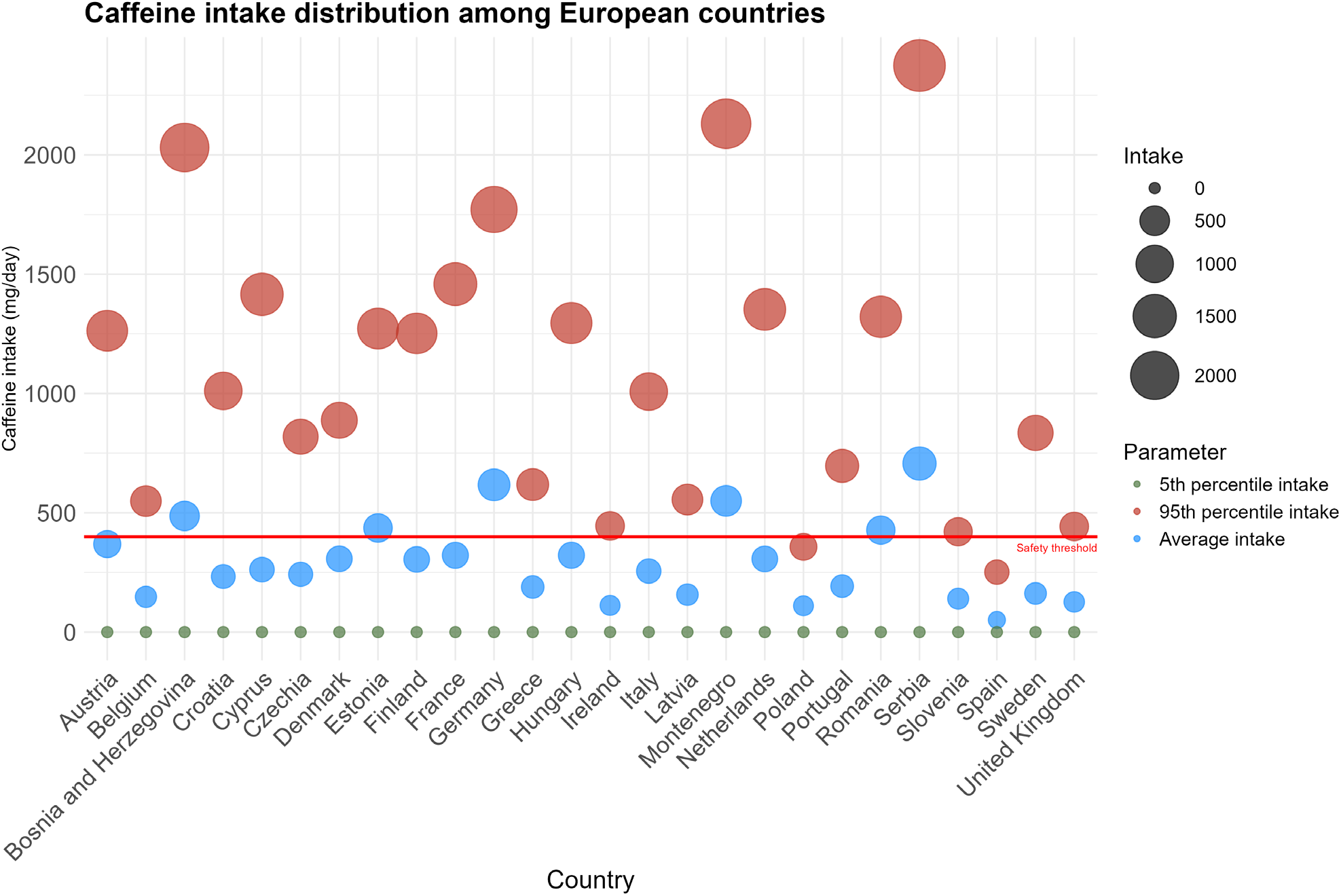
Caffeine intake differences in European countries. The consumption of specific phytochemicals, such as caffeine, can exert different health effects based on the amount consumed. We investigated the caffeine intake differences between the 5^th^ percentile, the average and the 95^th^ percentile of the population of each European country. The red line indicates a safety threshold of 400 mg/day proposed by EFSA upon chronic caffeine consumption for adults^20^.

## Discussion

In the current work, the intake of (poly)phenols, terpenoids, *N*-containing compounds, and miscellaneous phytochemicals in the European adult population was assessed, using the EFSA Comprehensive European Food Consumption Database and the PhytoFooD database. The daily exposure to more than 1,000 bioactive molecules in about 700 different foods consumed by the European adult population was evaluated. To our knowledge, this is the first attempt to simultaneously assess the intake of all dietary plant bioactives in the European population. Further analyses were performed to evaluate the contributions of different phytochemical classes, subclasses, and individual phytochemicals, and to explore their potential interconnections. Main food sources for each subclass of bioactive compounds were also identified. Insights into the variability of plant bioactive intake within European countries, considering multiple food consumption parameters, were provided. Finally, special consideration was given to differences in caffeine intake across and within European countries, emphasising the value of these results for safety evaluations and public health.

Our findings showed that plant bioactive intake in Europe is characterised by extensive variability: from a chemical perspective, remarkable differences were observed across phytochemical families and classes. Major divergences, reflecting their respective dietary habits, can also be observed among European countries. Nevertheless, our data align well with those reported for the EPIC cohort for (poly)phenols^14^, while some differences can be observed for carotenoid intake^16^. However, different dietary habits and carotenoid concentrations in foods may ultimately result in substantial variability in carotenoid intake^13^. Moreover, this work provides new figures on the daily consumption of the less studied bioactives in a comprehensive manner, and it also updates EPIC values, related mainly to the dietary habits of some countries from the late 90’s, to include new countries like Austria, Hungary and Portugal, using data from a period referred mainly from 2003 to 2021.

We shed light on the molecular complexity of the human diet and, consequently, on the disparities in the scientific literature regarding evidence for bioactive molecules. This is relevant, as (poly)phenols are often associated with the beneficial properties of many diet^21,22^, while the same diets may contain even higher amounts of other well-known phytochemicals with proven bioactivities. For instance, monoterpenoids were the phytochemical class with the highest intake, being about 1.3 g/day. However, their daily consumption and health effects on humans are poorly documented, and this lack of knowledge could be explained by the wide array of chemical structures, which also complicates current analytical challenge^23^. These observations can be extended to other classes of phytochemicals and to several highly consumed compounds, such as theanine, γ-muurolene, and *S*-Methyl-L-cysteine sulfoxide.

Our results further confirmed the intricate chemical nature of foods as complex matrices of structurally distinct bioactive molecules, which are, however, strongly correlated with one another. Additionally, the majority of these chemicals remain unexplored, and therefore their health implications are unknown^24^. These combined and interactive effects of food molecules will need to be considered in future research.

We also investigated how influential dietary patterns are in assessing phytochemical intake: the significant variability in plant bioactive exposure mirrored the diverse dietary habits within and among European countries. Moreover, in several cases, clear distinctions could be drawn between the plant bioactive intake of the average and the 95^th^ percentile of the population, revealing non-Gaussian traits of bioactive intake distributions. These data are usually considered when performing safety evaluations, like that of caffeine: indeed, our results regarding average caffeine intake are well aligned with EFSA recommendations^20^, but not when considering the 95^th^ percentile of the population. On the other hand, very low consumption of other health-relevant phytochemicals might be of significant importance for safety evaluations: for instance, low intakes of vitamin A precursors, such as α-carotene and β-carotene, can be associated with lower cognitive performance^25^. This information is also of greatest value when considering multi-exposure to chemicals within the same subclass, as their bioactivity can be similar.

Although this work represents a pioneering effort to comprehensively assess the phytochemical intake in the European adult population and, in general, of a large population, some limitations should be acknowledged. Firstly, the accuracy of dietary assessment tools, such as food records and dietary recalls, can be biased by their constitutive subjective nature^18^. To minimize these discrepancies and harmonize dietary data, EFSA provided precise guidelines for collecting food consumption information, enabling each country to reduce misreporting linked to subjective factors^26^. Secondly, for some countries, food consumption data at the 5^th^ and/or 95^th^ percentile of the population were not available, preventing us from properly evaluating their phytochemical intake distributions. Thirdly, the chemical composition of foods is widely influenced by multiple factors, such as cultivar type and the surrounding environment^27^. This intrinsic variability was not considered in this assessment, since only the average concentration of bioactives in foods was used. Lastly, the concentrations of chemicals in thermally processed foods were obtained using food yield factors; the absence of compound-specific retention factors could have led to inaccuracies in the global phytochemical assessment. These limitations associated with dietary assessment and composition tables should be embraced in the future with novel experimental designs, allowing for the characterisation of the dietary intake of each population more efficiently. To do so, efforts should focus on collecting standardised analytical data and dietary information across countries. Furthermore, incorporating dietary biomarkers into the assessment of phytochemical intake may provide a more objective and reliable evaluation^28^. In conclusion, this work provides the most comprehensive description of phytochemical intake in the diet of European adults, simultaneously considering multiple bioactive families and food sources. These results represent an extraordinary achievement in the global understanding of our diet, and they lay the groundwork for future evaluations of the role of dietary plant bioactives in human health.

## Methods

### Study population and food consumption data

The information on the dietary habits of the European adult population (aged 18-65 years) was extracted from the EFSA Comprehensive European Food Consumption Database (https://www.efsa.europa.eu/en/data-report/food-consumption-data), updated to December 2024.

The most recent surveys for each country were selected and used for this assessment. General characteristics of each European country survey, such as the name of the study considered, the number of individuals recruited and the food intake assessment method used are reported in Table S7. Different dietary assessment tools, mainly 24-hour dietary recalls and food records, were used to collect food consumption data: results were then harmonized and reported in the online database as g/day, as currently visible on the EFSA website.

Consumption data at the highest level of specificity (Exposure Hierarchy L7 based on FoodEx2 classification^29^) still reporting the name of food categories, for instance ‘*Leafy vegetables*’, were split into their constituent food items, based on the EFSA Catalogue browser structure^29^. Composite foods, such as “*Aioli or garlic sauce”*, were broken down into separate ingredients, aiming to reflect the original recipe as closely as possible.

### Phytochemical Food Database (PhytoFooD)

Data on the concentration of plant bioactive molecules in foods was obtained from the Phytochemical Food Database (PhytoFooD). Briefly, it collects data on the content of 1,067 phytochemicals in 1,410 different foods, divided into four main compound families: (poly)phenols, terpenoids, *N*-containing compounds, and miscellaneous phytochemicals. Each family is composed of several classes and subclasses: (poly)phenols are divided into 4 main classes, namely flavonoids, phenolic acids, tannins and miscellaneous (poly)phenols. Terpenoids are composed of 5 classes: tetra-, tri-, di-, sesqui- and monoterpenoids. *N*-containing compound family comprises 3 classes: alkaloids, glucosinolates & isothiocyanates, and miscellaneous *N*-containing compounds. These three families are subsequently divided into multiple subclasses of bioactives. Lastly, phytates, phytoprostanes, thiosulfinates, and miscellaneous compounds are the 4 main classes of the miscellaneous phytochemical family. PhytoFooD collects chemical information of both fresh and cooked foods from 13 national databases and targeted literature searches, providing average values after outlier assessment and removal. When data on cooked foods were not available, yield factors from Bognar’s tables^30^ and CREA^31^ were applied to convert food weight from raw to processed equivalents.

### Linkage between dietary data and PhytoFooD

Food items reported in the Exposure Hierarchy L7 from the EFSA Comprehensive European Food Consumption Database were univocally matched to our PhytoFooD food list, based on food names and macronutrient content. Otherwise, the most similar food from the catalogue was used. This step enabled us to link food consumption data to food composition data. The intake of individual bioactives was obtained by proportioning the phytochemical concentration of a specific food (expressed as mg/100g) to the amount of that food consumed. Intake of phytochemical subclasses, classes, and, finally, families was calculated by summing the individual bioactive intakes from different food sources, according to their classification. Multiple parameters were used to evaluate the extent of bioactive intake variability across Europe and its countries; particularly, food intake data at the 5^th^ percentile, the mean, and the 95^th^ percentile of the population were considered and matched to the mean concentration of plant bioactive compounds in each food. The complete phytochemical intake assessment was performed using Microsoft Excel spreadsheets. Statistical analysis and figures were carried out using R version 4.5.1 software (R Core Team 2025) (https://www.r-project.org/)^32^ and RStudio version 418 (Posit Software, 2025) (https://posit.co/). Graphical representations were created using following packages: tidyr^33^, dplyr^34^, ggplot2^35^, circlize^36^, ggbeeswarm^37^, ggridges^38^, scales^39^ and countrycode^40^.

## Data Availability

All data produced in the present study are available upon reasonable request to the authors

## Acknowledgements

This work was supported by the European Research Council (ERC) under the European Union’s Horizon 2020 research and innovation program (PREDICT-CARE project, grant agreement No 950050) and it has received funding from the Italian Ministry for Universities and Research (MUR) under the FARE program (CARE-DIET, R20MPBW4FM).

## Author contributions

C.Mic., A.R., D.D.R, and P.M. conceptualized the study. C.Mic., A.R. and F.B. collected, assembled and analysed the composition data for the development of the phytochemical food composition database. C.Mic. and F.B. analysed the data. C.Mic. drafted the paper. P.M. obtained funding for the study. All authors interpreted the results, revised the paper for important intellectual content, and read and approved the final paper.

## Supplementary materials

**Table S1: average intake (mg/day) of (poly)phenol family, classes, subclasses and individual compounds in the European adult population**.

**Table S2: average intake (mg/day) of terpenoid family, classes, subclasses and individual compounds in the European adult population**.

**Table S3: average intake (mg/day) of *N*-containing compound family, classes, subclasses and individual compounds in the European adult population**.

**Table S4: average intake (mg/day) of miscellaneous phytochemical family, classes, subclasses and individual compounds in the European adult population**.

**Table S5: phytochemical intake distributions (mg/day) of (poly)phenols, terpenoids, *N*-containing compounds and miscellaneous phytochemicals in 26 European countries**.

**Table S6: intake distribution (mg/day) of caffeine in Europe**. Missing values are attributable to unavailable food consumption data.

**Table S7: descriptive characteristics of European country surveys**.

---

**Fig. S1:**
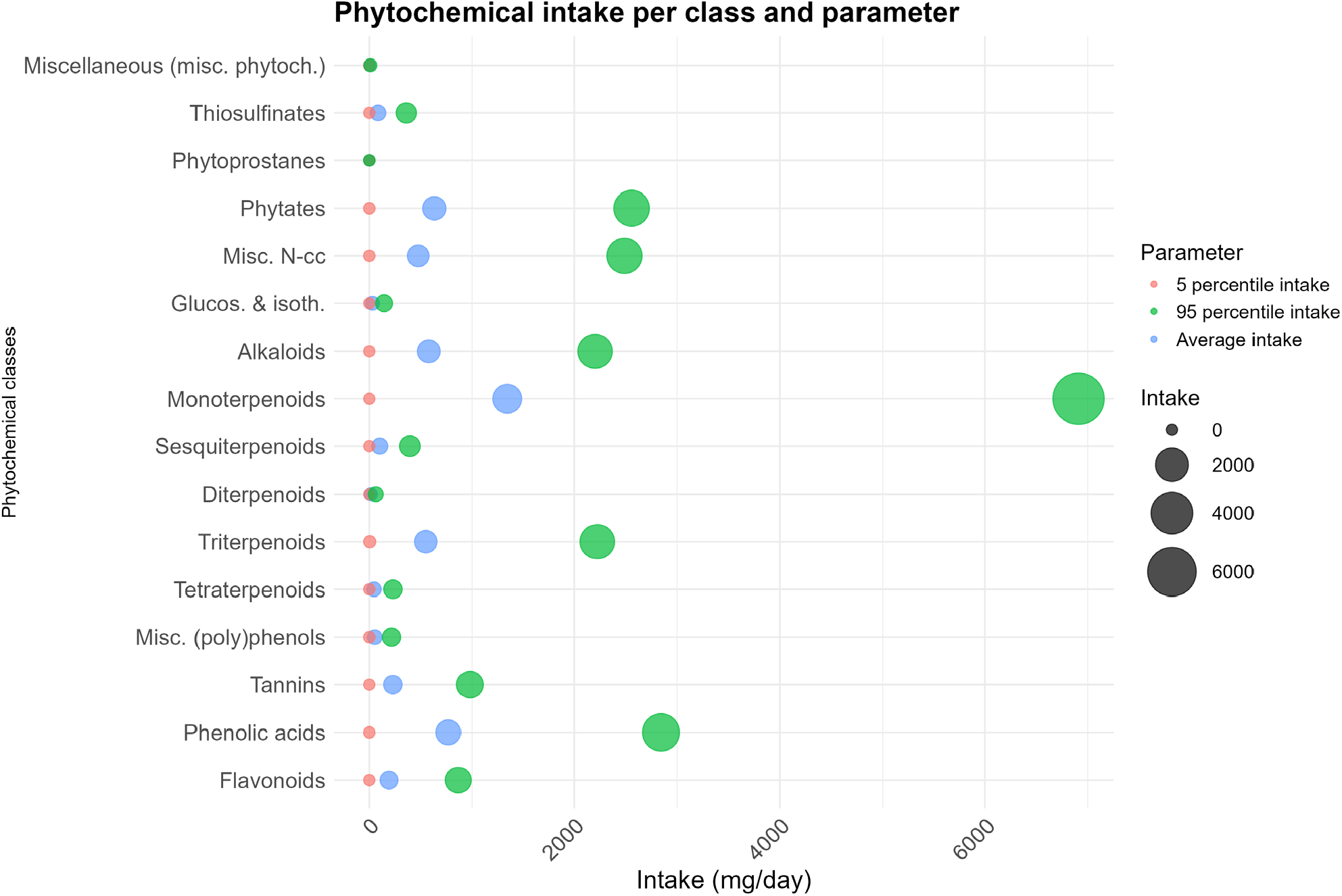
Intake distributions of plant bioactive classes in Europe. Food consumption data of the 5^th^ percentile, the average, and the 95^th^ percentile of the population of each European country were used to estimate the intake of dietary phytochemical classes in the European population.

