## Supplementary material for "Dietary intake of plant bioactives among European adults": Tables S1-S7

**Table S1: average intake of (poly)phenol family, classes, subclasses and individual compounds in the European adult population.**

| Family | Class | Subclass | Compound | Intake (mg/day) Mean ± SD |
|---|---|---|---|---|
| (Poly)phenols | Total | - | - | 1250.3 ± 367.9 |
| (Poly)phenols | Flavonoids | Total | - | 193.8 ± 67.7 |
| (Poly)phenols | Flavonoids | Flavonols | Total | 34.8 ± 10.5 |
| (Poly)phenols | Flavonoids | Flavonols | Dihydromyricetin 3-*O*-rhamnoside | 0.8 ± 0.7 |
| (Poly)phenols | Flavonoids | Flavonols | Dihydroquercetin 3-*O*-rhamnoside | 0.2 ± 0.2 |
| (Poly)phenols | Flavonoids | Flavonols | 6,8-Dihydroxykaempferol | 0 ± 0 |
| (Poly)phenols | Flavonoids | Flavonols | Galangin | 0 ± 0 |
| (Poly)phenols | Flavonoids | Flavonols | Kaempferol | 1.1 ± 0.9 |
| (Poly)phenols | Flavonoids | Flavonols | Kaempferol 3,7,4'-*O*-triglucoside | 0 ± 0 |
| (Poly)phenols | Flavonoids | Flavonols | Kaempferol 3,7-*O*-diglucoside | 0 ± 0 |
| (Poly)phenols | Flavonoids | Flavonols | Kaempferol 3-*O*-(2"-rhamnosyl-6"-acetyl-galactoside) 7-*O*-rhamnoside | 0 ± 0 |
| (Poly)phenols | Flavonoids | Flavonols | Kaempferol 3-*O*-(2"-rhamnosyl-galactoside) 7-*O*-rhamnoside | 0 ± 0 |
| (Poly)phenols | Flavonoids | Flavonols | Kaempferol 3-*O*-(6"-malonyl-glucoside) | 0.1 ± 0.1 |
| (Poly)phenols | Flavonoids | Flavonols | Kaempferol 3-*O*-(6"-acetyl-galactoside) 7-*O*-rhamnoside | 0 ± 0 |
| (Poly)phenols | Flavonoids | Flavonols | Kaempferol 3-*O*-acetyl-glucoside | 0.8 ± 0.7 |
| (Poly)phenols | Flavonoids | Flavonols | Kaempferol 3-*O*-galactoside | 0.5 ± 0.4 |
| (Poly)phenols | Flavonoids | Flavonols | Kaempferol 3-*O*-galactoside 7-*O*-rhamnoside | 0 ± 0 |
| (Poly)phenols | Flavonoids | Flavonols | Kaempferol 3-*O*-glucoside | 3.5 ± 2 |
| (Poly)phenols | Flavonoids | Flavonols | Kaempferol 3-*O*-glucosyl-rhamnosyl-galactoside | 0 ± 0 |
| (Poly)phenols | Flavonoids | Flavonols | Kaempferol 3-*O*-glucosyl-rhamnosyl-glucoside | 0 ± 0 |
| (Poly)phenols | Flavonoids | Flavonols | Kaempferol 3-*O*-glucuronide | 0.4 ± 0.6 |
| (Poly)phenols | Flavonoids | Flavonols | Kaempferol 3-*O*-rhamnoside | 0 ± 0 |
| (Poly)phenols | Flavonoids | Flavonols | Kaempferol 3-*O*-rhamnosyl-rhamnosyl-glucoside | 0.2 ± 0.2 |
| (Poly)phenols | Flavonoids | Flavonols | Kaempferol 3-*O*-rutinoside | 3.2 ± 2 |
| (Poly)phenols | Flavonoids | Flavonols | Kaempferol 3-*O*-sophoroside | 0.6 ± 0.3 |
| (Poly)phenols | Flavonoids | Flavonols | Kaempferol 3-*O*-sophoroside 7-*O*-glucoside | 0 ± 0 |
| (Poly)phenols | Flavonoids | Flavonols | Kaempferol 3-*O*-xylosyl-glucoside | 0.6 ± 0.5 |
| (Poly)phenols | Flavonoids | Flavonols | Kaempferol 3-*O*-xylosyl-rutinoside | 0 ± 0 |
| (Poly)phenols | Flavonoids | Flavonols | Kaempferol 7-*O*-glucoside | 0 ± 0 |
| (Poly)phenols | Flavonoids | Flavonols | Morin | 0 ± 0 |
| (Poly)phenols | Flavonoids | Flavonols | Myricetin | 0.2 ± 0.1 |
| (Poly)phenols | Flavonoids | Flavonols | Myricetin 3-*O*-arabinoside | 0 ± 0.1 |
| (Poly)phenols | Flavonoids | Flavonols | Myricetin 3-*O*-galactoside | 0 ± 0 |
| (Poly)phenols | Flavonoids | Flavonols | Myricetin 3-*O*-glucoside | 0 ± 0 |
| (Poly)phenols | Flavonoids | Flavonols | Myricetin 3-*O*-rhamnoside | 0 ± 0 |

| Family | Class | Subclass | Compound | Intake (mg/day) Mean ± SD |
|---|---|---|---|---|
| (Poly)phenols | Flavonoids | Flavonols | Myricetin 3-*O*-rutinoside | 0 ± 0 |
| (Poly)phenols | Flavonoids | Flavonols | Quercetin | 1.5 ± 0.6 |
| (Poly)phenols | Flavonoids | Flavonols | Quercetin 3,4'-*O*-diglucoside | 0.6 ± 0.3 |
| (Poly)phenols | Flavonoids | Flavonols | Quercetin 3-*O*-(6"-malonyl-glucoside) | 0.2 ± 0.1 |
| (Poly)phenols | Flavonoids | Flavonols | Quercetin 3-*O*-(6"-malonyl-glucoside) 7-*O*-glucoside | 0 ± 0 |
| (Poly)phenols | Flavonoids | Flavonols | Quercetin 3-*O*-(6"-acetyl-galactoside) 7-*O*-rhamnoside | 0 ± 0 |
| (Poly)phenols | Flavonoids | Flavonols | Quercetin 3-*O*-acetyl-rhamnoside | 0 ± 0 |
| (Poly)phenols | Flavonoids | Flavonols | Quercetin 3-*O*-arabinoside | 0.3 ± 0.1 |
| (Poly)phenols | Flavonoids | Flavonols | Quercetin 3-*O*-galactoside | 1.4 ± 0.8 |
| (Poly)phenols | Flavonoids | Flavonols | Quercetin 3-*O*-galactoside 7-*O*-rhamnoside | 0 ± 0 |
| (Poly)phenols | Flavonoids | Flavonols | Quercetin 3-*O*-glucoside | 1.9 ± 1.5 |
| (Poly)phenols | Flavonoids | Flavonols | Quercetin 3-*O*-glucosyl-rhamnosyl-galactoside | 0 ± 0 |
| (Poly)phenols | Flavonoids | Flavonols | Quercetin 3-*O*-glucosyl-rhamnosyl-glucoside | 1 ± 1.1 |
| (Poly)phenols | Flavonoids | Flavonols | Quercetin 3-*O*-glucosyl-xyloside | 0 ± 0 |
| (Poly)phenols | Flavonoids | Flavonols | Quercetin 3-*O*-glucuronide | 0.4 ± 0.2 |
| (Poly)phenols | Flavonoids | Flavonols | Quercetin 3-*O*-rhamnoside | 0.9 ± 0.3 |
| (Poly)phenols | Flavonoids | Flavonols | Quercetin 3-*O*-rhamnosyl-galactoside | 0 ± 0 |
| (Poly)phenols | Flavonoids | Flavonols | Quercetin 3-*O*-rhamnosyl-rhamnosyl-glucoside | 0 ± 0 |
| (Poly)phenols | Flavonoids | Flavonols | Quercetin 3-*O*-rutinoside | 6.8 ± 3.1 |
| (Poly)phenols | Flavonoids | Flavonols | Quercetin 3-*O*-sophoroside | 0.2 ± 0.1 |
| (Poly)phenols | Flavonoids | Flavonols | Quercetin 3-*O*-xyloside | 0.1 ± 0 |
| (Poly)phenols | Flavonoids | Flavonols | Quercetin 3-*O*-xylosyl-glucuronide | 0 ± 0 |
| (Poly)phenols | Flavonoids | Flavonols | Quercetin 3-*O*-xylosyl-rutinoside | 0 ± 0 |
| (Poly)phenols | Flavonoids | Flavonols | Quercetin 4'-*O*-glucoside | 0.4 ± 0.2 |
| (Poly)phenols | Flavonoids | Flavonols | Quercetin 7,4'-*O*-diglucoside | 0 ± 0 |
| (Poly)phenols | Flavonoids | Flavonols | 3,7-Dimethylquercetin | 0 ± 0 |
| (Poly)phenols | Flavonoids | Flavonols | 3-Methoxynobiletin | 0 ± 0 |
| (Poly)phenols | Flavonoids | Flavonols | 3-Methoxysinensetin | 0 ± 0 |
| (Poly)phenols | Flavonoids | Flavonols | 5,3',4'-Trihydroxy-3-methoxy-6:7-methylenedioxyflavone 4'-*O*-glucoside | 0.4 ± 0.3 |
| (Poly)phenols | Flavonoids | Flavonols | 5,4'-Dihydroxy-3,3'-dimethoxy-6:7-methylenedioxyflavone 4'-*O*-glucoside | 1 ± 0.9 |
| (Poly)phenols | Flavonoids | Flavonols | Isorhamnetin | 0.1 ± 0.1 |
| (Poly)phenols | Flavonoids | Flavonols | Isorhamnetin 3-*O*-galactoside | 0 ± 0 |
| (Poly)phenols | Flavonoids | Flavonols | Isorhamnetin 3-*O*-glucoside | 0.1 ± 0 |
| (Poly)phenols | Flavonoids | Flavonols | Isorhamnetin 3-*O*-glucoside 7-*O*-rhamnoside | 0 ± 0 |
| (Poly)phenols | Flavonoids | Flavonols | Isorhamnetin 3-O-glucuronide | 0 ± 0 |

| Family | Class | Subclass | Compound | Intake (mg/day) Mean ± SD |
|---|---|---|---|---|
| (Poly)phenols | Flavonoids | Flavonols | Isorhamnetin 3-*O*-rutinoside | 0 ± 0 |
| (Poly)phenols | Flavonoids | Flavonols | Isorhamnetin 4'-*O*-glucoside | 0 ± 0 |
| (Poly)phenols | Flavonoids | Flavonols | Isorhamnetin 7-*O*-rhamnoside | 0 ± 0 |
| (Poly)phenols | Flavonoids | Flavonols | Jaceidin 4'-*O*-glucuronide | 0.1 ± 0.1 |
| (Poly)phenols | Flavonoids | Flavonols | Kaempferide | 0 ± 0 |
| (Poly)phenols | Flavonoids | Flavonols | Methylgalangin | 0 ± 0 |
| (Poly)phenols | Flavonoids | Flavonols | Patuletin 3-*O*-(2"-feruloylglucosyl)(1->6)-apiosyl(1->2)-glucoside | 0.3 ± 0.2 |
| (Poly)phenols | Flavonoids | Flavonols | Patuletin 3-*O*-glucosyl-(1->6)-apiosyl(1->2)-glucoside | 0.5 ± 0.4 |
| (Poly)phenols | Flavonoids | Flavonols | Rhamnetin | 0 ± 0 |
| (Poly)phenols | Flavonoids | Flavonols | Spinacetin 3-*O*-(2"-feruloylglucosyl)(1->6)-apiosyl(1->2)-glucoside | 0.4 ± 0.3 |
| (Poly)phenols | Flavonoids | Flavonols | Spinacetin 3-*O*-(2"-p-coumaroylglucosyl)(1->6)-apiosyl(1->2)-glucoside | 0 ± 0 |
| (Poly)phenols | Flavonoids | Flavonols | Spinacetin 3-*O*-glucosyl-(1->6)-apiosyl(1->2)-glucoside | 0.3 ± 0.3 |
| (Poly)phenols | Flavonoids | Flavonols | Spinacetin 3-*O*-glucosyl-(1->6)-glucoside | 0.2 ± 0.2 |
| (Poly)phenols | Flavonoids | Flavan-3-ol monomers | Total | 100.3 ± 56.1 |
| (Poly)phenols | Flavonoids | Flavan-3-ol monomers | (+)-Catechin | 8.7 ± 3.2 |
| (Poly)phenols | Flavonoids | Flavan-3-ol monomers | (+)-Catechin 3-*O*-gallate | 5.1 ± 5.2 |
| (Poly)phenols | Flavonoids | Flavan-3-ol monomers | (+)-Catechin 3-*O*-glucose | 0.1 ± 0.1 |
| (Poly)phenols | Flavonoids | Flavan-3-ol monomers | (+)-Gallocatechin | 15.6 ± 15.5 |
| (Poly)phenols | Flavonoids | Flavan-3-ol monomers | (+)-Gallocatechin 3-*O*-gallate | 0.8 ± 0.7 |
| (Poly)phenols | Flavonoids | Flavan-3-ol monomers | (-)-Epicatechin | 15.6 ± 5.8 |
| (Poly)phenols | Flavonoids | Flavan-3-ol monomers | (-)-Epicatechin 3-*O*-gallate | 9.6 ± 8.2 |
| (Poly)phenols | Flavonoids | Flavan-3-ol monomers | (-)-Epigallocatechin | 11.4 ± 8.7 |
| (Poly)phenols | Flavonoids | Flavan-3-ol monomers | (-)-Epigallocatechin 3-*O*-gallate | 14.7 ± 11.3 |
| (Poly)phenols | Flavonoids | Flavan-3-ol monomers | 01 mers | 18.7 ± 7.3 |
| (Poly)phenols | Flavonoids | Flavanones | Total | 22.3 ± 11.3 |
| (Poly)phenols | Flavonoids | Flavanones | Didymin | 1.6 ± 0.9 |
| (Poly)phenols | Flavonoids | Flavanones | Eriocitrin | 3.4 ± 8.5 |
| (Poly)phenols | Flavonoids | Flavanones | Eriodictyol | 0 ± 0 |
| (Poly)phenols | Flavonoids | Flavanones | Eriodictyol 7-*O*-glucoside | 0 ± 0.1 |
| (Poly)phenols | Flavonoids | Flavanones | Hesperidin | 8.2 ± 4.3 |

| Family | Class | Subclass | Compound | Intake (mg/day) Mean ± SD |
|---|---|---|---|---|
| (Poly)phenols | Flavonoids | Flavanones | Naringenin | 0.1 ± 0 |
| (Poly)phenols | Flavonoids | Flavanones | Naringenin 7-*O*-glucoside | 0.1 ± 0 |
| (Poly)phenols | Flavonoids | Flavanones | Naringin | 1.1 ± 0.7 |
| (Poly)phenols | Flavonoids | Flavanones | Naringin 4'-*O*-glucoside | 0 ± 0 |
| (Poly)phenols | Flavonoids | Flavanones | Naringin 6'-malonate | 0 ± 0 |
| (Poly)phenols | Flavonoids | Flavanones | Narirutin | 1.8 ± 0.8 |
| (Poly)phenols | Flavonoids | Flavanones | Narirutin 4'-*O*-glucoside | 0 ± 0 |
| (Poly)phenols | Flavonoids | Flavanones | Neoeriocitrin | 0 ± 0 |
| (Poly)phenols | Flavonoids | Flavanones | Neohesperidin | 0 ± 0 |
| (Poly)phenols | Flavonoids | Flavanones | Pinocembrin | 0 ± 0 |
| (Poly)phenols | Flavonoids | Flavanones | Poncirin | 0.1 ± 0 |
| (Poly)phenols | Flavonoids | Flavanones | Hesperetin | 0 ± 0 |
| (Poly)phenols | Flavonoids | Flavanones | Sakuranetin | 0 ± 0 |
| (Poly)phenols | Flavonoids | Flavones | Total | 17.7 ± 6.7 |
| (Poly)phenols | Flavonoids | Flavones | 6-Hydroxyluteolin | 0 ± 0 |
| (Poly)phenols | Flavonoids | Flavones | 6-Hydroxyluteolin 7-*O*-rhamnoside | 0 ± 0 |
| (Poly)phenols | Flavonoids | Flavones | 7,3',4'-Trihydroxyflavone | 0 ± 0 |
| (Poly)phenols | Flavonoids | Flavones | 7,4'-Dihydroxyflavone | 0 ± 0 |
| (Poly)phenols | Flavonoids | Flavones | Apigenin | 0.1 ± 0 |
| (Poly)phenols | Flavonoids | Flavones | Apigenin 6,8-*C*-arabinoside-*C*-glucoside | 4.3 ± 1.5 |
| (Poly)phenols | Flavonoids | Flavones | Apigenin 6,8-*C*-galactoside-*C*-arabinoside | 6.4 ± 2.3 |
| (Poly)phenols | Flavonoids | Flavones | Apigenin 6,8-di-*C*-glucoside | 1.4 ± 0.8 |
| (Poly)phenols | Flavonoids | Flavones | Apigenin 6-*C*-glucoside | 0 ± 0 |
| (Poly)phenols | Flavonoids | Flavones | Apigenin 7-*O*-(6''-malonyl-apiosyl-glucoside) | 0 ± 0 |
| (Poly)phenols | Flavonoids | Flavones | Apigenin 7-*O*-apiosyl-glucoside | 0 ± 0 |
| (Poly)phenols | Flavonoids | Flavones | Apigenin 7-*O*-diglucuronide | 0 ± 0 |
| (Poly)phenols | Flavonoids | Flavones | Apigenin 7-*O*-glucoside | 0.1 ± 0.1 |
| (Poly)phenols | Flavonoids | Flavones | Apigenin 7-*O*-glucuronide | 0.1 ± 0.1 |
| (Poly)phenols | Flavonoids | Flavones | Baicalein | 0 ± 0 |
| (Poly)phenols | Flavonoids | Flavones | Chrysin | 0 ± 0 |
| (Poly)phenols | Flavonoids | Flavones | Isorhoifolin | 0.1 ± 0.1 |
| (Poly)phenols | Flavonoids | Flavones | Luteolin | 0.6 ± 0.4 |
| (Poly)phenols | Flavonoids | Flavones | Luteolin 6-*C*-glucoside | 0 ± 0 |
| (Poly)phenols | Flavonoids | Flavones | Luteolin 7-*O*-(2-apiosyl-6-malonyl)-glucoside | 0.2 ± 0.1 |
| (Poly)phenols | Flavonoids | Flavones | Luteolin 7-*O*-(2-apiosyl-glucoside) | 0 ± 0 |
| (Poly)phenols | Flavonoids | Flavones | Luteolin 7-*O*-diglucuronide | 0 ± 0 |
| (Poly)phenols | Flavonoids | Flavones | Luteolin 7-*O*-glucoside | 0.2 ± 0.1 |
| (Poly)phenols | Flavonoids | Flavones | Luteolin 7-*O*-glucuronide | 0.1 ± 0.1 |
| (Poly)phenols | Flavonoids | Flavones | Luteolin 7-*O*-malonyl-glucoside | 0 ± 0 |
| (Poly)phenols | Flavonoids | Flavones | Luteolin 7-*O*-rutinoside | 0.4 ± 1.2 |
| (Poly)phenols | Flavonoids | Flavones | Rhoifolin | 0 ± 0 |
| (Poly)phenols | Flavonoids | Flavones | Rhoifolin 4'-*O*-glucoside | 0 ± 0 |

| Family | Class | Subclass | Compound | Intake (mg/day) Mean ± SD |
|---|---|---|---|---|
| (Poly)phenols | Flavonoids | Flavones | Scutellarein | 0 ± 0 |
| (Poly)phenols | Flavonoids | Flavones | 5,6-Dihydroxy-7,8,3',4'-tetramethoxyflavone | 0 ± 0 |
| (Poly)phenols | Flavonoids | Flavones | Chrysoeriol 7-*O*-(6"-malonyl-apiosyl-glucoside) | 0 ± 0 |
| (Poly)phenols | Flavonoids | Flavones | Chrysoeriol 7-*O*-(6"-malonyl-glucoside) | 0 ± 0 |
| (Poly)phenols | Flavonoids | Flavones | Chrysoeriol 7-*O*-apiosyl-glucoside | 0 ± 0 |
| (Poly)phenols | Flavonoids | Flavones | Chrysoeriol 7-*O*-glucoside | 0 ± 0 |
| (Poly)phenols | Flavonoids | Flavones | Cirsilineol | 0 ± 0 |
| (Poly)phenols | Flavonoids | Flavones | Cirsimaritin | 0 ± 0 |
| (Poly)phenols | Flavonoids | Flavones | Diosmin | 0.1 ± 0.1 |
| (Poly)phenols | Flavonoids | Flavones | Eupatorin | 0 ± 0 |
| (Poly)phenols | Flavonoids | Flavones | Gardenin B | 0 ± 0 |
| (Poly)phenols | Flavonoids | Flavones | Geraldone | 0 ± 0 |
| (Poly)phenols | Flavonoids | Flavones | Hispidulin | 0 ± 0.1 |
| (Poly)phenols | Flavonoids | Flavones | Jaceosidin | 0 ± 0 |
| (Poly)phenols | Flavonoids | Flavones | Neodiosmin | 0 ± 0 |
| (Poly)phenols | Flavonoids | Flavones | Nepetin | 0 ± 0 |
| (Poly)phenols | Flavonoids | Flavones | Nobiletin | 0 ± 0 |
| (Poly)phenols | Flavonoids | Flavones | Pebrellin | 0 ± 0 |
| (Poly)phenols | Flavonoids | Flavones | Sinensetin | 0 ± 0 |
| (Poly)phenols | Flavonoids | Flavones | Tangeretin | 0 ± 0 |
| (Poly)phenols | Flavonoids | Flavones | Tetramethylscutellarein | 0 ± 0 |
| (Poly)phenols | Flavonoids | Isoflavones | Total | 1.4 ± 1.5 |
| (Poly)phenols | Flavonoids | Isoflavones | 6"-*O*-Acetyldaidzin | 0 ± 0.1 |
| (Poly)phenols | Flavonoids | Isoflavones | 6"-*O*-Acetylgenistin | 0.1 ± 0.1 |
| (Poly)phenols | Flavonoids | Isoflavones | 6"-*O*-Malonyldaidzin | 0.1 ± 0.1 |
| (Poly)phenols | Flavonoids | Isoflavones | 6"-*O*-Malonylgenistin | 0.2 ± 0.3 |
| (Poly)phenols | Flavonoids | Isoflavones | Daidzein | 0 ± 0 |
| (Poly)phenols | Flavonoids | Isoflavones | Daidzin | 0.3 ± 0.3 |
| (Poly)phenols | Flavonoids | Isoflavones | Genistein | 0.1 ± 0.1 |
| (Poly)phenols | Flavonoids | Isoflavones | Genistin | 0.4 ± 0.4 |
| (Poly)phenols | Flavonoids | Isoflavones | 6"-*O*-Acetylglycitin | 0 ± 0 |
| (Poly)phenols | Flavonoids | Isoflavones | 6"-*O*-Malonylglycitin | 0 ± 0.1 |
| (Poly)phenols | Flavonoids | Isoflavones | Biochanin A | 0 ± 0 |
| (Poly)phenols | Flavonoids | Isoflavones | Formononetin | 0 ± 0 |
| (Poly)phenols | Flavonoids | Isoflavones | Glycitein | 0 ± 0 |
| (Poly)phenols | Flavonoids | Isoflavones | Glycitin | 0 ± 0.1 |
| (Poly)phenols | Flavonoids | Anthocyanins | Total | 17.1 ± 13 |
| (Poly)phenols | Flavonoids | Anthocyanins | Cyanidin | 0 ± 0 |
| (Poly)phenols | Flavonoids | Anthocyanins | Cyanidin 3,5-*O*-diglucoside | 0 ± 0 |
| (Poly)phenols | Flavonoids | Anthocyanins | Cyanidin 3-*O*-(6"-acetyl-galactoside) | 0 ± 0 |
| (Poly)phenols | Flavonoids | Anthocyanins | Cyanidin 3-*O*-(6"-acetyl-glucoside) | 0 ± 0 |

| Family | Class | Subclass | Compound | Intake (mg/day) Mean ± SD |
|---|---|---|---|---|
| (Poly)phenols | Flavonoids | Anthocyanins | Cyanidin 3-*O*-(6"-caffeoyl-glucoside) | 0 ± 0 |
| (Poly)phenols | Flavonoids | Anthocyanins | Cyanidin 3-*O*-(6"-dioxalyl-glucoside) | 0 ± 0 |
| (Poly)phenols | Flavonoids | Anthocyanins | Cyanidin 3-O-(6"-malonyl-3"-glucosyl-glucoside) | 0 ± 0 |
| (Poly)phenols | Flavonoids | Anthocyanins | Cyanidin 3-*O*-(6"-malonyl-glucoside) | 0 ± 0 |
| (Poly)phenols | Flavonoids | Anthocyanins | Cyanidin 3-*O*-(6"-*p*-coumaroyl-glucoside) | 0 ± 0 |
| (Poly)phenols | Flavonoids | Anthocyanins | Cyanidin 3-*O*-(6"-succinyl-glucoside) | 0.1 ± 0 |
| (Poly)phenols | Flavonoids | Anthocyanins | Cyanidin 3-*O*-arabinoside | 0 ± 0 |
| (Poly)phenols | Flavonoids | Anthocyanins | Cyanidin 3-*O*-galactoside | 0.1 ± 0.1 |
| (Poly)phenols | Flavonoids | Anthocyanins | Cyanidin 3-*O*-glucoside | 1.1 ± 0.5 |
| (Poly)phenols | Flavonoids | Anthocyanins | Cyanidin 3-*O*-glucosyl-rutinoside | 0.2 ± 0.2 |
| (Poly)phenols | Flavonoids | Anthocyanins | Cyanidin 3-*O*-rutinoside | 3.9 ± 1.9 |
| (Poly)phenols | Flavonoids | Anthocyanins | Cyanidin 3-*O*-sambubioside | 0 ± 0.1 |
| (Poly)phenols | Flavonoids | Anthocyanins | Cyanidin 3-*O*-sambubioside 5-*O*-glucoside | 0 ± 0 |
| (Poly)phenols | Flavonoids | Anthocyanins | Cyanidin 3-*O*-sophoroside | 0.3 ± 0.2 |
| (Poly)phenols | Flavonoids | Anthocyanins | Cyanidin 3-*O*-xyloside | 0 ± 0 |
| (Poly)phenols | Flavonoids | Anthocyanins | Cyanidin 3-*O*-xylosyl-rutinoside | 0 ± 0 |
| (Poly)phenols | Flavonoids | Anthocyanins | Delphinidin 3,5-*O*-diglucoside | 0 ± 0 |
| (Poly)phenols | Flavonoids | Anthocyanins | Delphinidin 3-*O*-(6"-acetyl-galactoside) | 0 ± 0 |
| (Poly)phenols | Flavonoids | Anthocyanins | Delphinidin 3-*O*-(6"-acetyl-glucoside) | 0.1 ± 0.1 |
| (Poly)phenols | Flavonoids | Anthocyanins | Delphinidin 3-*O*-(6"-*p*-coumaroyl-glucoside) | 0 ± 0 |
| (Poly)phenols | Flavonoids | Anthocyanins | Delphinidin 3-*O*-arabinoside | 0.1 ± 0.1 |
| (Poly)phenols | Flavonoids | Anthocyanins | Delphinidin 3-*O*-feruloyl-glucoside | 0 ± 0 |
| (Poly)phenols | Flavonoids | Anthocyanins | Delphinidin 3-*O*-galactoside | 0.1 ± 0.1 |
| (Poly)phenols | Flavonoids | Anthocyanins | Delphinidin 3-*O*-glucoside | 0.4 ± 0.3 |
| (Poly)phenols | Flavonoids | Anthocyanins | Delphinidin 3-*O*-glucosyl-glucoside | 0 ± 0 |
| (Poly)phenols | Flavonoids | Anthocyanins | Delphinidin 3-*O*-rutinoside | 0.5 ± 1 |
| (Poly)phenols | Flavonoids | Anthocyanins | Delphinidin 3-*O*-sambubioside | 0 ± 0 |
| (Poly)phenols | Flavonoids | Anthocyanins | Delphinidin 3-*O*-xyloside | 0 ± 0 |
| (Poly)phenols | Flavonoids | Anthocyanins | Malvidin 3,5-*O*-diglucoside | 0 ± 0 |
| (Poly)phenols | Flavonoids | Anthocyanins | Malvidin 3-*O*-(6"-acetyl-galactoside) | 0 ± 0 |
| (Poly)phenols | Flavonoids | Anthocyanins | Malvidin 3-*O*-(6"-acetyl-glucoside) | 0.6 ± 0.6 |
| (Poly)phenols | Flavonoids | Anthocyanins | Malvidin 3-*O*-(6"-caffeoyl-glucoside) | 0 ± 0 |
| (Poly)phenols | Flavonoids | Anthocyanins | Malvidin 3-*O*-(6"-*p*-coumaroyl-glucoside) | 0.2 ± 0.2 |
| (Poly)phenols | Flavonoids | Anthocyanins | Malvidin 3-*O*-arabinoside | 0.1 ± 0.1 |
| (Poly)phenols | Flavonoids | Anthocyanins | Malvidin 3-*O*-galactoside | 0.1 ± 0.1 |

| Family | Class | Subclass | Compound | Intake (mg/day) Mean ± SD |
|---|---|---|---|---|
| (Poly)phenols | Flavonoids | Anthocyanins | Malvidin 3-*O*-glucoside | 1.8 ± 1.6 |
| (Poly)phenols | Flavonoids | Anthocyanins | Pelargonidin | 0.2 ± 0.1 |
| (Poly)phenols | Flavonoids | Anthocyanins | Pelargonidin 3,5-*O*-diglucoside | 0 ± 0 |
| (Poly)phenols | Flavonoids | Anthocyanins | Pelargonidin 3-*O*-(6"-malonyl-glucoside) | 0.2 ± 0.1 |
| (Poly)phenols | Flavonoids | Anthocyanins | Pelargonidin 3-*O*-(6"-succinyl-glucoside) | 0.5 ± 0.3 |
| (Poly)phenols | Flavonoids | Anthocyanins | Pelargonidin 3-*O*-arabinoside | 0 ± 0 |
| (Poly)phenols | Flavonoids | Anthocyanins | Pelargonidin 3-*O*-galactoside | 0 ± 0 |
| (Poly)phenols | Flavonoids | Anthocyanins | Pelargonidin 3-*O*-glucoside | 2.1 ± 1.2 |
| (Poly)phenols | Flavonoids | Anthocyanins | Pelargonidin 3-*O*-glucosyl-rutinoside | 0 ± 0 |
| (Poly)phenols | Flavonoids | Anthocyanins | Pelargonidin 3-*O*-rutinoside | 0.1 ± 0 |
| (Poly)phenols | Flavonoids | Anthocyanins | Pelargonidin 3-*O*-sambubioside | 0 ± 0 |
| (Poly)phenols | Flavonoids | Anthocyanins | Pelargonidin 3-*O*-sophoroside | 0 ± 0 |
| (Poly)phenols | Flavonoids | Anthocyanins | Peonidin | 0 ± 0 |
| (Poly)phenols | Flavonoids | Anthocyanins | Peonidin 3-*O*-(6"-acetyl-galactoside) | 0 ± 0 |
| (Poly)phenols | Flavonoids | Anthocyanins | Peonidin 3-*O*-(6"-acetyl-glucoside) | 0.1 ± 0.1 |
| (Poly)phenols | Flavonoids | Anthocyanins | Peonidin 3-*O*-(6"-*p*-coumaroyl-glucoside) | 0 ± 0 |
| (Poly)phenols | Flavonoids | Anthocyanins | Peonidin 3-*O*-arabinoside | 0 ± 0 |
| (Poly)phenols | Flavonoids | Anthocyanins | Peonidin 3-*O*-galactoside | 0 ± 0 |
| (Poly)phenols | Flavonoids | Anthocyanins | Peonidin 3-*O*-glucoside | 0.2 ± 0.1 |
| (Poly)phenols | Flavonoids | Anthocyanins | Peonidin 3-*O*-rutinoside | 0.2 ± 0.1 |
| (Poly)phenols | Flavonoids | Anthocyanins | Petunidin 3,5-*O*-diglucoside | 0 ± 0 |
| (Poly)phenols | Flavonoids | Anthocyanins | Petunidin 3-*O*-(6"-acetyl-galactoside) | 0 ± 0 |
| (Poly)phenols | Flavonoids | Anthocyanins | Petunidin 3-*O*-(6"-acetyl-glucoside) | 0.1 ± 0.1 |
| (Poly)phenols | Flavonoids | Anthocyanins | Petunidin 3-*O*-(6"-*p*-coumaroyl-glucoside) | 0.1 ± 0.1 |
| (Poly)phenols | Flavonoids | Anthocyanins | Petunidin 3-*O*-arabinoside | 0 ± 0 |
| (Poly)phenols | Flavonoids | Anthocyanins | Petunidin 3-*O*-galactoside | 0.1 ± 0.1 |
| (Poly)phenols | Flavonoids | Anthocyanins | Petunidin 3-*O*-glucoside | 0.3 ± 0.2 |
| (Poly)phenols | Flavonoids | Anthocyanins | Petunidin 3-*O*-rhamnoside | 0 ± 0 |
| (Poly)phenols | Flavonoids | Anthocyanins | Petunidin 3-*O*-rutinoside | 0 ± 0 |
| (Poly)phenols | Flavonoids | Anthocyanins | Pigment A | 0.2 ± 0.2 |
| (Poly)phenols | Flavonoids | Anthocyanins | Pinotin A | 0 ± 0 |
| (Poly)phenols | Flavonoids | Anthocyanins | Vitisin A | 0 ± 0 |
| (Poly)phenols | Phenolic acids | Total | - | 772.4 ± 314.4 |
| (Poly)phenols | Phenolic acids | Cinnamic acids | Total | 743.1 ± 312 |
| (Poly)phenols | Phenolic acids | Cinnamic acids | Caffeoyl tartaric acid | 1.2 ± 0.7 |
| (Poly)phenols | Phenolic acids | Cinnamic acids | *p*-Coumaric acid | 0.5 ± 0.2 |
| (Poly)phenols | Phenolic acids | Cinnamic acids | *p*-Coumaroyl tartaric acid | 0.3 ± 0.2 |

| Family | Class | Subclass | Compound | Intake (mg/day) Mean ± SD |
|---|---|---|---|---|
| (Poly)phenols | Phenolic acids | Cinnamic acids | Ferulic acid | 6.4 ± 1.8 |
| (Poly)phenols | Phenolic acids | Cinnamic acids | 2,5-di-*S*-Glutathionyl caftaric acid | 0.2 ± 0.2 |
| (Poly)phenols | Phenolic acids | Cinnamic acids | 2-*S*-Glutathionyl caftaric acid | 0 ± 0 |
| (Poly)phenols | Phenolic acids | Cinnamic acids | 3,4-Dicaffeoylquinic acid | 8.8 ± 3.9 |
| (Poly)phenols | Phenolic acids | Cinnamic acids | 3,5-Dicaffeoylquinic acid | 5.5 ± 2.4 |
| (Poly)phenols | Phenolic acids | Cinnamic acids | 3-Caffeoylquinic acid | 160.3 ± 72.9 |
| (Poly)phenols | Phenolic acids | Cinnamic acids | 3-*p*-Coumaroylquinic acid | 0.9 ± 0.5 |
| (Poly)phenols | Phenolic acids | Cinnamic acids | 4,5-Dicaffeoylquinic acid | 6.9 ± 3.1 |
| (Poly)phenols | Phenolic acids | Cinnamic acids | 4-Caffeoylquinic acid | 181.6 ± 83.8 |
| (Poly)phenols | Phenolic acids | Cinnamic acids | 4-*p*-Coumaroylquinic acid | 1.4 ± 1 |
| (Poly)phenols | Phenolic acids | Cinnamic acids | 5-Caffeoylquinic acid | 257.4 ± 101.2 |
| (Poly)phenols | Phenolic acids | Cinnamic acids | 5-*p*-Coumaroylquinic acid | 0.1 ± 0.1 |
| (Poly)phenols | Phenolic acids | Cinnamic acids | Caffeic acid | 2.4 ± 0.6 |
| (Poly)phenols | Phenolic acids | Cinnamic acids | Caffeic acid 4-*O*-glucoside | 0 ± 0 |
| (Poly)phenols | Phenolic acids | Cinnamic acids | Caffeic acid ethyl ester | 0 ± 0 |
| (Poly)phenols | Phenolic acids | Cinnamic acids | Caffeoyl aspartic acid | 0.3 ± 0.2 |
| (Poly)phenols | Phenolic acids | Cinnamic acids | Caffeoyl glucose | 0 ± 0 |
| (Poly)phenols | Phenolic acids | Cinnamic acids | Chicoric acid | 0.6 ± 1.2 |
| (Poly)phenols | Phenolic acids | Cinnamic acids | Cinnamic acid | 0.1 ± 0.1 |
| (Poly)phenols | Phenolic acids | Cinnamic acids | Cinnamoyl glucose | 0 ± 0 |
| (Poly)phenols | Phenolic acids | Cinnamic acids | Hydroxycaffeic acid | 0 ± 0 |
| (Poly)phenols | Phenolic acids | Cinnamic acids | *m*-Coumaric acid | 0.1 ± 0.1 |
| (Poly)phenols | Phenolic acids | Cinnamic acids | *o*-Coumaric acid | 0.5 ± 0.6 |
| (Poly)phenols | Phenolic acids | Cinnamic acids | *p*-Coumaric acid 4-*O*-glucoside | 0 ± 0 |
| (Poly)phenols | Phenolic acids | Cinnamic acids | *p*-Coumaric acid ethyl ester | 0 ± 0 |
| (Poly)phenols | Phenolic acids | Cinnamic acids | *p*-Coumaroyl glucose | 0.3 ± 0.1 |

| Family | Class | Subclass | Compound | Intake (mg/day) Mean ± SD |
|---|---|---|---|---|
| (Poly)phenols | Phenolic acids | Cinnamic acids | *p*-Coumaroyl glycolic acid | 0 ± 0 |
| (Poly)phenols | Phenolic acids | Cinnamic acids | *p*-Coumaroyl malic acid | 0 ± 0 |
| (Poly)phenols | Phenolic acids | Cinnamic acids | *p*-Coumaroyl tartaric acid glucosidic ester | 0 ± 0 |
| (Poly)phenols | Phenolic acids | Cinnamic acids | *p*-Coumaroyl tyrosine | 0 ± 0 |
| (Poly)phenols | Phenolic acids | Cinnamic acids | *p*-Coumaroylquinic acid | 0.6 ± 0.3 |
| (Poly)phenols | Phenolic acids | Cinnamic acids | Rosmarinic acid | 0.9 ± 1.9 |
| (Poly)phenols | Phenolic acids | Cinnamic acids | Verbascoside | 0.4 ± 0.4 |
| (Poly)phenols | Phenolic acids | Cinnamic acids | 1,2,2'-Triferuloylgentiobiose | 0 ± 0 |
| (Poly)phenols | Phenolic acids | Cinnamic acids | 1,2,2'-Trisinapoylgentiobiose | 0 ± 0 |
| (Poly)phenols | Phenolic acids | Cinnamic acids | 1,2-Diferuloylgentiobiose | 0.1 ± 0.1 |
| (Poly)phenols | Phenolic acids | Cinnamic acids | 1,2'-Disinapoyl-2-feruloylgentiobiose | 0 ± 0 |
| (Poly)phenols | Phenolic acids | Cinnamic acids | 1,2-Disinapoylgentiobiose | 0.1 ± 0 |
| (Poly)phenols | Phenolic acids | Cinnamic acids | 1-Sinapoyl-2,2'-diferuloylgentiobiose | 0 ± 0 |
| (Poly)phenols | Phenolic acids | Cinnamic acids | 1-Sinapoyl-2-feruloylgentiobiose | 0 ± 0 |
| (Poly)phenols | Phenolic acids | Cinnamic acids | 24-Methylcholestanol ferulate | 0 ± 0 |
| (Poly)phenols | Phenolic acids | Cinnamic acids | 24-Methylcholesterol ferulate | 0 ± 0 |
| (Poly)phenols | Phenolic acids | Cinnamic acids | 24-Methylenecholestanol ferulate | 0 ± 0 |
| (Poly)phenols | Phenolic acids | Cinnamic acids | 24-Methyllathosterol ferulate | 0 ± 0 |
| (Poly)phenols | Phenolic acids | Cinnamic acids | 3,4-Diferuloylquinic acid | 0 ± 0 |
| (Poly)phenols | Phenolic acids | Cinnamic acids | 3,5-Diferuloylquinic acid | 0 ± 0 |
| (Poly)phenols | Phenolic acids | Cinnamic acids | 3-Feruloylquinic acid | 12.5 ± 5.8 |
| (Poly)phenols | Phenolic acids | Cinnamic acids | 3-Sinapoylquinic acid | 0 ± 0 |
| (Poly)phenols | Phenolic acids | Cinnamic acids | 4-Feruloylquinic acid | 25.9 ± 12.1 |
| (Poly)phenols | Phenolic acids | Cinnamic acids | 4-Sinapoylquinic acid | 0 ± 0 |
| (Poly)phenols | Phenolic acids | Cinnamic acids | 5-Feruloylquinic acid | 35 ± 16.4 |
| (Poly)phenols | Phenolic acids | Cinnamic acids | 5-Sinapoylquinic acid | 0 ± 0 |

| Family | Class | Subclass | Compound | Intake (mg/day) Mean ± SD |
|---|---|---|---|---|
| (Poly)phenols | Phenolic acids | Cinnamic acids | Ferulic acid 4-*O*-glucoside | 0 ± 0 |
| (Poly)phenols | Phenolic acids | Cinnamic acids | Feruloyl glucose | 0.1 ± 0 |
| (Poly)phenols | Phenolic acids | Cinnamic acids | Feruloyl tartaric acid | 0 ± 0 |
| (Poly)phenols | Phenolic acids | Cinnamic acids | Isoferulic acid | 0 ± 0 |
| (Poly)phenols | Phenolic acids | Cinnamic acids | Schottenol ferulate | 0 ± 0 |
| (Poly)phenols | Phenolic acids | Cinnamic acids | Sinapic acid | 0.5 ± 0.2 |
| (Poly)phenols | Phenolic acids | Cinnamic acids | Sinapine | 0 ± 0 |
| (Poly)phenols | Phenolic acids | Cinnamic acids | Sitosterol ferulate | 0 ± 0 |
| (Poly)phenols | Phenolic acids | Cinnamic acids | Stigmastanol ferulate | 0.1 ± 0.1 |
| (Poly)phenols | Phenolic acids | Cinnamic acids | 5-5'-Dehydrodiferulic acid | 0.3 ± 0.2 |
| (Poly)phenols | Phenolic acids | Cinnamic acids | 5-8'-Benzofuran dehydrodiferulic acid | 0.3 ± 0.3 |
| (Poly)phenols | Phenolic acids | Cinnamic acids | 5-8'-Dehydrodiferulic acid | 0.4 ± 0.3 |
| (Poly)phenols | Phenolic acids | Cinnamic acids | 8-*O*-4'-Dehydrodiferulic acid | 0.5 ± 0.4 |
| (Poly)phenols | Phenolic acids | Phenylpropanoic acids | Total | 0 ± 0 |
| (Poly)phenols | Phenolic acids | Phenylpropanoic acids | Dihydrocaffeic acid | 0 ± 0 |
| (Poly)phenols | Phenolic acids | Phenylpropanoic acids | Dihydro-*p*-coumaric acid | 0 ± 0 |
| (Poly)phenols | Phenolic acids | Phenylacetic acids | Total | 0.2 ± 0.1 |
| (Poly)phenols | Phenolic acids | Phenylacetic acids | 3,4-Dihydroxyphenylacetic acid | 0 ± 0 |
| (Poly)phenols | Phenolic acids | Phenylacetic acids | 4-Hydroxyphenylacetic acid | 0.1 ± 0.1 |
| (Poly)phenols | Phenolic acids | Phenylacetic acids | Homovanillic acid | 0.1 ± 0 |
| (Poly)phenols | Phenolic acids | Phenylacetic acids | Homoveratric acid | 0 ± 0 |
| (Poly)phenols | Phenolic acids | Phenylacetic acids | Methoxyphenylacetic acid | 0 ± 0 |
| (Poly)phenols | Phenolic acids | Benzoic acids | Total | 29 ± 17.9 |
| (Poly)phenols | Phenolic acids | Benzoic acids | 2,3-Dihydroxybenzoic acid | 0 ± 0 |
| (Poly)phenols | Phenolic acids | Benzoic acids | 2,4-Dihydroxybenzoic acid | 0 ± 0 |
| (Poly)phenols | Phenolic acids | Benzoic acids | 2,6-Dihydroxybenzoic acid | 0.1 ± 0.1 |
| (Poly)phenols | Phenolic acids | Benzoic acids | 2-Hydroxybenzoic acid | 0.2 ± 0.2 |
| (Poly)phenols | Phenolic acids | Benzoic acids | 3,5-Dihydroxybenzoic acid | 0 ± 0 |
| (Poly)phenols | Phenolic acids | Benzoic acids | 3-Hydroxybenzoic acid | 0 ± 0 |

| Family | Class | Subclass | Compound | Intake (mg/day) Mean ± SD |
|---|---|---|---|---|
| (Poly)phenols | Phenolic acids | Benzoic acids | 4-Hydroxybenzoic acid | 1.2 ± 0.8 |
| (Poly)phenols | Phenolic acids | Benzoic acids | 4-Hydroxybenzoic acid 4-*O*-glucoside | 0.1 ± 0 |
| (Poly)phenols | Phenolic acids | Benzoic acids | 5-*O*-Galloylquinic acid | 14.2 ± 12.9 |
| (Poly)phenols | Phenolic acids | Benzoic acids | Benzoic acid | 0 ± 0 |
| (Poly)phenols | Phenolic acids | Benzoic acids | Gallic acid | 9.2 ± 5 |
| (Poly)phenols | Phenolic acids | Benzoic acids | Gallic acid 3-*O*-gallate | 0.3 ± 0.2 |
| (Poly)phenols | Phenolic acids | Benzoic acids | Gallic acid 4-*O*-glucoside | 0 ± 0 |
| (Poly)phenols | Phenolic acids | Benzoic acids | Gallic acid ethyl ester | 0.3 ± 0.2 |
| (Poly)phenols | Phenolic acids | Benzoic acids | Galloyl glucose | 0 ± 0 |
| (Poly)phenols | Phenolic acids | Benzoic acids | Gentisic acid | 0.4 ± 0.2 |
| (Poly)phenols | Phenolic acids | Benzoic acids | Protocatechuic acid | 0.7 ± 0.6 |
| (Poly)phenols | Phenolic acids | Benzoic acids | Protocatechuic acid 4-*O*-glucoside | 0 ± 0 |
| (Poly)phenols | Phenolic acids | Benzoic acids | Syringic acid | 0.5 ± 0.2 |
| (Poly)phenols | Phenolic acids | Benzoic acids | Vanillic acid | 0.3 ± 0.1 |
| (Poly)phenols | Phenolic acids | Benzoic acids | Guaiacol | 0.5 ± 0.2 |
| (Poly)phenols | Phenolic acids | Miscellaneous phenolic acids | Total | 0 ± 0 |
| (Poly)phenols | Phenolic acids | Miscellaneous phenolic acids | Avenanthramide 2f | 0 ± 0 |
| (Poly)phenols | Phenolic acids | Miscellaneous phenolic acids | Avenanthramide 2c | 0 ± 0 |
| (Poly)phenols | Phenolic acids | Miscellaneous phenolic acids | Avenanthramide 2p | 0 ± 0 |
| (Poly)phenols | Phenolic acids | Miscellaneous phenolic acids | Avenanthramide K | 0 ± 0 |
| (Poly)phenols | Tannins | Total | - | 230.8 ± 48.9 |
| (Poly)phenols | Tannins | Proanthocyanidins & theaflavins | Total | 225.2 ± 48 |
| (Poly)phenols | Tannins | Proanthocyanidins & theaflavins | (-)-Epicatechin-(2a-7)(4a-8)-epicatechin 3-*O*-galactoside | 0.2 ± 0.1 |
| (Poly)phenols | Tannins | Proanthocyanidins & theaflavins | Cinnamtannin A2 | 1.7 ± 1 |
| (Poly)phenols | Tannins | Proanthocyanidins & theaflavins | Procyanidin dimer B1 | 10 ± 3.9 |
| (Poly)phenols | Tannins | Proanthocyanidins & theaflavins | Procyanidin dimer B2 | 11.7 ± 4.6 |
| (Poly)phenols | Tannins | Proanthocyanidins & theaflavins | Procyanidin dimer B3 | 2.8 ± 1.7 |
| (Poly)phenols | Tannins | Proanthocyanidins & theaflavins | Procyanidin dimer B4 | 3.6 ± 2.4 |

| Family | Class | Subclass | Compound | Intake (mg/day) Mean ± SD |
|---|---|---|---|---|
| (Poly)phenols | Tannins | Proanthocyanidins & theaflavins | Procyanidin dimer B5 | 0.4 ± 0.1 |
| (Poly)phenols | Tannins | Proanthocyanidins & theaflavins | Procyanidin dimer B7 | 2.1 ± 0.7 |
| (Poly)phenols | Tannins | Proanthocyanidins & theaflavins | Procyanidin trimer C1 | 9.2 ± 8 |
| (Poly)phenols | Tannins | Proanthocyanidins & theaflavins | Procyanidin trimer C2 | 0 ± 0 |
| (Poly)phenols | Tannins | Proanthocyanidins & theaflavins | Procyanidin trimer EEC | 0.7 ± 0.2 |
| (Poly)phenols | Tannins | Proanthocyanidins & theaflavins | Procyanidin trimer T2 | 0.2 ± 0.2 |
| (Poly)phenols | Tannins | Proanthocyanidins & theaflavins | Prodelphinidin dimer B3 | 2.1 ± 1.9 |
| (Poly)phenols | Tannins | Proanthocyanidins & theaflavins | Prodelphinidin trimer C-GC-C | 0 ± 0 |
| (Poly)phenols | Tannins | Proanthocyanidins & theaflavins | Prodelphinidin trimer GC-C-C | 0 ± 0 |
| (Poly)phenols | Tannins | Proanthocyanidins & theaflavins | Prodelphinidin trimer GC-GC-C | 0 ± 0 |
| (Poly)phenols | Tannins | Proanthocyanidins & theaflavins | 02 mers | 17 ± 5.4 |
| (Poly)phenols | Tannins | Proanthocyanidins & theaflavins | 03 mers | 15.1 ± 5.3 |
| (Poly)phenols | Tannins | Proanthocyanidins & theaflavins | 04 mers | 5.4 ± 3.2 |
| (Poly)phenols | Tannins | Proanthocyanidins & theaflavins | 04-06 mers | 41.6 ± 9 |
| (Poly)phenols | Tannins | Proanthocyanidins & theaflavins | 07-10 mers | 28.6 ± 5.4 |
| (Poly)phenols | Tannins | Proanthocyanidins & theaflavins | polymers (>10 mers) | 68.8 ± 13.8 |
| (Poly)phenols | Tannins | Proanthocyanidins & theaflavins | Theaflavin | 1 ± 1.1 |

| Family | Class | Subclass | Compound | Intake (mg/day) Mean ± SD |
|---|---|---|---|---|
| (Poly)phenols | Tannins | Proanthocyanidins & theaflavins | Theaflavin 3,3'-*O*-digallate | 1 ± 1.1 |
| (Poly)phenols | Tannins | Proanthocyanidins & theaflavins | Theaflavin 3'-*O*-gallate | 1 ± 1.1 |
| (Poly)phenols | Tannins | Proanthocyanidins & theaflavins | Theaflavin 3-*O*-gallate | 1 ± 1.1 |
| (Poly)phenols | Tannins | Gallotannins | Total | 0 ± 0 |
| (Poly)phenols | Tannins | Phlorotannins | Total | 0 ± 0 |
| (Poly)phenols | Tannins | Ellagitannins | Total | 5.6 ± 3.8 |
| (Poly)phenols | Tannins | Ellagitannins | Ellagic acid | 4.4 ± 3.7 |
| (Poly)phenols | Tannins | Ellagitannins | Ellagic acid acetyl-arabinoside | 0 ± 0 |
| (Poly)phenols | Tannins | Ellagitannins | Ellagic acid acetyl-xyloside | 0 ± 0 |
| (Poly)phenols | Tannins | Ellagitannins | Ellagic acid arabinoside | 0 ± 0 |
| (Poly)phenols | Tannins | Ellagitannins | Ellagic acid glucoside | 0.1 ± 0.1 |
| (Poly)phenols | Tannins | Ellagitannins | Valoneic acid dilactone | 1.4 ± 0.8 |
| (Poly)phenols | Tannins | Ellagitannins | Lambertianin C | 0.2 ± 0.2 |
| (Poly)phenols | Tannins | Ellagitannins | Punicalagin | 0.1 ± 0.3 |
| (Poly)phenols | Tannins | Ellagitannins | Sanguiin H-6 | 0.5 ± 0.4 |
| (Poly)phenols | Miscellaneous (poly)phenols | Total | - | 53.4 ± 1.7 |
| (Poly)phenols | Miscellaneous (poly)phenols | Lignans | Total | 12 ± 0.6 |
| (Poly)phenols | Miscellaneous (poly)phenols | Lignans | 1-Acetoxypinoresinol | 0.1 ± 0.1 |
| (Poly)phenols | Miscellaneous (poly)phenols | Lignans | 7-Hydroxymatairesinol | 0 ± 0 |
| (Poly)phenols | Miscellaneous (poly)phenols | Lignans | 7-Hydroxysecoisolariciresinol | 0 ± 0 |
| (Poly)phenols | Miscellaneous (poly)phenols | Lignans | 7-Oxomatairesinol | 0 ± 0 |
| (Poly)phenols | Miscellaneous (poly)phenols | Lignans | Anhydro-secoisolariciresinol | 0 ± 0 |
| (Poly)phenols | Miscellaneous (poly)phenols | Lignans | Arctigenin | 0 ± 0 |
| (Poly)phenols | Miscellaneous (poly)phenols | Lignans | Conidendrin | 0 ± 0 |
| (Poly)phenols | Miscellaneous (poly)phenols | Lignans | Cyclolariciresinol | 0 ± 0 |
| (Poly)phenols | Miscellaneous (poly)phenols | Lignans | Dimethylmatairesinol | 0 ± 0 |
| (Poly)phenols | Miscellaneous (poly)phenols | Lignans | Episesamin | 0 ± 0.1 |
| (Poly)phenols | Miscellaneous (poly)phenols | Lignans | Episesaminol | 0 ± 0 |
| (Poly)phenols | Miscellaneous (poly)phenols | Lignans | Isohydroxymatairesinol | 0 ± 0 |

| Family | Class | Subclass | Compound | Intake (mg/day) Mean ± SD |
|---|---|---|---|---|
| (Poly)phenols | Miscellaneous (poly)phenols | Lignans | Isolariciresinol | 0 ± 0 |
| (Poly)phenols | Miscellaneous (poly)phenols | Lignans | Lariciresinol | 0.3 ± 0 |
| (Poly)phenols | Miscellaneous (poly)phenols | Lignans | Lariciresinol-sesquilignan | 0 ± 0 |
| (Poly)phenols | Miscellaneous (poly)phenols | Lignans | Matairesinol | 0.1 ± 0 |
| (Poly)phenols | Miscellaneous (poly)phenols | Lignans | Medioresinol | 0 ± 0 |
| (Poly)phenols | Miscellaneous (poly)phenols | Lignans | Nortrachelogenin | 0 ± 0 |
| (Poly)phenols | Miscellaneous (poly)phenols | Lignans | Pinoresinol | 0.2 ± 0.1 |
| (Poly)phenols | Miscellaneous (poly)phenols | Lignans | Secoisolariciresinol | 0.5 ± 0.3 |
| (Poly)phenols | Miscellaneous (poly)phenols | Lignans | Secoisolariciresinol-sesquilignan | 0 ± 0 |
| (Poly)phenols | Miscellaneous (poly)phenols | Lignans | Sesamin | 0 ± 0 |
| (Poly)phenols | Miscellaneous (poly)phenols | Lignans | Sesaminol | 0.2 ± 0.2 |
| (Poly)phenols | Miscellaneous (poly)phenols | Lignans | Sesamol | 0 ± 0 |
| (Poly)phenols | Miscellaneous (poly)phenols | Lignans | Sesamolin | 0.2 ± 0.2 |
| (Poly)phenols | Miscellaneous (poly)phenols | Lignans | Sesamolinol | 0 ± 0 |
| (Poly)phenols | Miscellaneous (poly)phenols | Lignans | Syringaresinol | 0.3 ± 0.1 |
| (Poly)phenols | Miscellaneous (poly)phenols | Lignans | Trachelogenin | 0 ± 0 |
| (Poly)phenols | Miscellaneous (poly)phenols | Stilbenes | Total | 0.8 ± 0.6 |
| (Poly)phenols | Miscellaneous (poly)phenols | Stilbenes | *d*-Viniferin | 0 ± 0 |
| (Poly)phenols | Miscellaneous (poly)phenols | Stilbenes | *e*-Viniferin | 0 ± 0 |
| (Poly)phenols | Miscellaneous (poly)phenols | Stilbenes | Pallidol | 0 ± 0 |
| (Poly)phenols | Miscellaneous (poly)phenols | Stilbenes | Piceatannol | 0 ± 0 |
| (Poly)phenols | Miscellaneous (poly)phenols | Stilbenes | Piceatannol 3-*O*-glucoside | 0.2 ± 0.2 |
| (Poly)phenols | Miscellaneous (poly)phenols | Stilbenes | Pinosylvin | 0 ± 0 |
| (Poly)phenols | Miscellaneous (poly)phenols | Stilbenes | Pterostilbene | 0 ± 0 |
| (Poly)phenols | Miscellaneous (poly)phenols | Stilbenes | Resveratrol | 0.1 ± 0 |
| (Poly)phenols | Miscellaneous (poly)phenols | Stilbenes | Resveratrol 3-*O*-glucoside | 0.2 ± 0.1 |

| Family | Class | Subclass | Compound | Intake (mg/day) Mean ± SD |
|---|---|---|---|---|
| (Poly)phenols | Miscellaneous (poly)phenols | Stilbenes | Resveratrol 5-*O*-glucoside | 0 ± 0 |
| (Poly)phenols | Miscellaneous (poly)phenols | Coumarins & furanocoumarins | Total | 0.3 ± 0.2 |
| (Poly)phenols | Miscellaneous (poly)phenols | Coumarins & furanocoumarins | Bergapten | 0.1 ± 0.1 |
| (Poly)phenols | Miscellaneous (poly)phenols | Coumarins & furanocoumarins | Isopimpinellin | 0 ± 0 |
| (Poly)phenols | Miscellaneous (poly)phenols | Coumarins & furanocoumarins | Psoralen | 0 ± 0.1 |
| (Poly)phenols | Miscellaneous (poly)phenols | Coumarins & furanocoumarins | Xanthotoxin | 0 ± 0 |
| (Poly)phenols | Miscellaneous (poly)phenols | Coumarins & furanocoumarins | 4-Hydroxycoumarin | 0.2 ± 0.1 |
| (Poly)phenols | Miscellaneous (poly)phenols | Coumarins & furanocoumarins | Coumarin | 0 ± 0 |
| (Poly)phenols | Miscellaneous (poly)phenols | Coumarins & furanocoumarins | Esculetin | 0 ± 0 |
| (Poly)phenols | Miscellaneous (poly)phenols | Coumarins & furanocoumarins | Esculin | 0 ± 0 |
| (Poly)phenols | Miscellaneous (poly)phenols | Coumarins & furanocoumarins | Mellein | 0 ± 0 |
| (Poly)phenols | Miscellaneous (poly)phenols | Coumarins & furanocoumarins | Scopoletin | 0 ± 0 |
| (Poly)phenols | Miscellaneous (poly)phenols | Coumarins & furanocoumarins | Umbelliferone | 0 ± 0 |
| (Poly)phenols | Miscellaneous (poly)phenols | Chalcones | Total | 1.9 ± 1.4 |
| (Poly)phenols | Miscellaneous (poly)phenols | Chalcones | Butein | 0 ± 0 |
| (Poly)phenols | Miscellaneous (poly)phenols | Chalcones | 3-Hydroxyphloretin 2'-*O*-glucoside | 0 ± 0 |
| (Poly)phenols | Miscellaneous (poly)phenols | Chalcones | 3-Hydroxyphloretin 2'-*O*-xylosyl-glucoside | 0 ± 0 |
| (Poly)phenols | Miscellaneous (poly)phenols | Chalcones | Phloretin | 0 ± 0 |
| (Poly)phenols | Miscellaneous (poly)phenols | Chalcones | Phloretin 2'-*O*-xylosyl-glucoside | 0.7 ± 0.5 |
| (Poly)phenols | Miscellaneous (poly)phenols | Chalcones | Phloridzin | 1.2 ± 0.9 |

| Family | Class | Subclass | Compound | Intake (mg/day) Mean ± SD |
|---|---|---|---|---|
| (Poly)phenols | Miscellaneous (poly)phenols | Prenylflavonoids | Total | 0 ± 0 |
| (Poly)phenols | Miscellaneous (poly)phenols | Prenylflavonoids | Xanthohumol | 0 ± 0 |
| (Poly)phenols | Miscellaneous (poly)phenols | Prenylflavonoids | 6-Geranylnaringenin | 0 ± 0 |
| (Poly)phenols | Miscellaneous (poly)phenols | Prenylflavonoids | 6-Prenylnaringenin | 0 ± 0 |
| (Poly)phenols | Miscellaneous (poly)phenols | Prenylflavonoids | 8-Prenylnaringenin | 0 ± 0 |
| (Poly)phenols | Miscellaneous (poly)phenols | Prenylflavonoids | Isoxanthohumol | 0 ± 0 |
| (Poly)phenols | Miscellaneous (poly)phenols | Alkylresorcinols | Total | 29.8 ± 9.3 |
| (Poly)phenols | Miscellaneous (poly)phenols | Alkylresorcinols | 3-Methylcatechol | 0.3 ± 0.1 |
| (Poly)phenols | Miscellaneous (poly)phenols | Alkylresorcinols | 4-Ethylcatechol | 0.4 ± 0.2 |
| (Poly)phenols | Miscellaneous (poly)phenols | Alkylresorcinols | 4-Ethylphenol | 0 ± 0 |
| (Poly)phenols | Miscellaneous (poly)phenols | Alkylresorcinols | 4-Methylcatechol | 0.1 ± 0.1 |
| (Poly)phenols | Miscellaneous (poly)phenols | Alkylresorcinols | 4-Vinylphenol | 0 ± 0 |
| (Poly)phenols | Miscellaneous (poly)phenols | Alkylresorcinols | 5-Heneicosenylresorcinol | 0.9 ± 1.3 |
| (Poly)phenols | Miscellaneous (poly)phenols | Alkylresorcinols | 5-Heneicosylresorcinol | 10.9 ± 3.1 |
| (Poly)phenols | Miscellaneous (poly)phenols | Alkylresorcinols | 5-Heptadecylresorcinol | 2.7 ± 2 |
| (Poly)phenols | Miscellaneous (poly)phenols | Alkylresorcinols | 5-Nonadecenylresorcinol | 0.9 ± 0.4 |
| (Poly)phenols | Miscellaneous (poly)phenols | Alkylresorcinols | 5-Nonadecylresorcinol | 9.9 ± 3.1 |
| (Poly)phenols | Miscellaneous (poly)phenols | Alkylresorcinols | 5-Pentacosenylresorcinol | 0.1 ± 0.1 |
| (Poly)phenols | Miscellaneous (poly)phenols | Alkylresorcinols | 5-Pentacosylresorcinol | 1.2 ± 0.6 |
| (Poly)phenols | Miscellaneous (poly)phenols | Alkylresorcinols | 5-Pentadecylresorcinol | 0 ± 0 |
| (Poly)phenols | Miscellaneous (poly)phenols | Alkylresorcinols | 5-Tricosenylresorcinol | 0.2 ± 0.3 |
| (Poly)phenols | Miscellaneous (poly)phenols | Alkylresorcinols | 5-Tricosylresorcinol | 2.3 ± 0.9 |
| (Poly)phenols | Miscellaneous (poly)phenols | Miscellaneous | Total | 18.8 ± 6 |
| (Poly)phenols | Miscellaneous (poly)phenols | Miscellaneous | Todolactol A | 0 ± 0 |
| (Poly)phenols | Miscellaneous (poly)phenols | Miscellaneous | 4-Ethylguaiacol | 2 ± 0.9 |
| (Poly)phenols | Miscellaneous (poly)phenols | Miscellaneous | 4-Vinylguaiacol | 1.6 ± 0.7 |

| Family | Class | Subclass | Compound | Intake (mg/day) Mean ± SD |
|---|---|---|---|---|
| (Poly)phenols | Miscellaneous (poly)phenols | Miscellaneous | 4-Vinylsyringol | 0.1 ± 0.3 |
| (Poly)phenols | Miscellaneous (poly)phenols | Miscellaneous | Bisdemethoxycurcumin | 0 ± 0 |
| (Poly)phenols | Miscellaneous (poly)phenols | Miscellaneous | Curcumin | 0 ± 0 |
| (Poly)phenols | Miscellaneous (poly)phenols | Miscellaneous | Demethoxycurcumin | 0 ± 0 |
| (Poly)phenols | Miscellaneous (poly)phenols | Miscellaneous | 4-Hydroxybenzaldehyde | 0 ± 0 |
| (Poly)phenols | Miscellaneous (poly)phenols | Miscellaneous | Gallic aldehyde | 0 ± 0 |
| (Poly)phenols | Miscellaneous (poly)phenols | Miscellaneous | *p*-Anisaldehyde | 0 ± 0 |
| (Poly)phenols | Miscellaneous (poly)phenols | Miscellaneous | Protocatechuic aldehyde | 0 ± 0 |
| (Poly)phenols | Miscellaneous (poly)phenols | Miscellaneous | Syringaldehyde | 0.1 ± 0.1 |
| (Poly)phenols | Miscellaneous (poly)phenols | Miscellaneous | Vanillin | 0 ± 0 |
| (Poly)phenols | Miscellaneous (poly)phenols | Miscellaneous | 2,3-Dihydroxy-1-guaiacylpropanone | 0 ± 0 |
| (Poly)phenols | Miscellaneous (poly)phenols | Miscellaneous | 3-Methoxyacetophenone | 0 ± 0 |
| (Poly)phenols | Miscellaneous (poly)phenols | Miscellaneous | Ferulaldehyde | 0 ± 0 |
| (Poly)phenols | Miscellaneous (poly)phenols | Miscellaneous | Sinapaldehyde | 0 ± 0 |
| (Poly)phenols | Miscellaneous (poly)phenols | Miscellaneous | 6-Gingerol | 0 ± 0 |
| (Poly)phenols | Miscellaneous (poly)phenols | Miscellaneous | 2-Methoxy-5-prop-1-enylphenol | 0 ± 0 |
| (Poly)phenols | Miscellaneous (poly)phenols | Miscellaneous | Acetyl eugenol | 0.1 ± 0.1 |
| (Poly)phenols | Miscellaneous (poly)phenols | Miscellaneous | Anethole | 0 ± 0 |
| (Poly)phenols | Miscellaneous (poly)phenols | Miscellaneous | Eugenol | 0.3 ± 0.4 |
| (Poly)phenols | Miscellaneous (poly)phenols | Miscellaneous | 1,4-Naphtoquinone | 0 ± 0 |
| (Poly)phenols | Miscellaneous (poly)phenols | Miscellaneous | Juglone | 0 ± 0 |
| (Poly)phenols | Miscellaneous (poly)phenols | Miscellaneous | 3,4-Dihydroxyphenylglycol | 0 ± 0 |
| (Poly)phenols | Miscellaneous (poly)phenols | Miscellaneous | Arbutin | 0.1 ± 0.1 |
| (Poly)phenols | Miscellaneous (poly)phenols | Miscellaneous | Catechol | 1.2 ± 0.6 |
| (Poly)phenols | Miscellaneous (poly)phenols | Miscellaneous | Coumestrol | 0 ± 0 |
| (Poly)phenols | Miscellaneous (poly)phenols | Miscellaneous | Phenol | 0.4 ± 0.2 |

| Family | Class | Subclass | Compound | Intake (mg/day) Mean ± SD |
|---|---|---|---|---|
| (Poly)phenols | Miscellaneous (poly)phenols | Miscellaneous | Phlorin | 0.8 ± 0.4 |
| (Poly)phenols | Miscellaneous (poly)phenols | Miscellaneous | Pyrogallol | 1.6 ± 0.8 |
| (Poly)phenols | Miscellaneous (poly)phenols | Miscellaneous | Capsaicin | 2.7 ± 2 |
| (Poly)phenols | Miscellaneous (poly)phenols | Miscellaneous | Dihydrocapsaicin | 1.4 ± 1.1 |
| (Poly)phenols | Miscellaneous (poly)phenols | Miscellaneous | Carnosic acid | 0.3 ± 0.5 |
| (Poly)phenols | Miscellaneous (poly)phenols | Miscellaneous | Carnosol | 0 ± 0 |
| (Poly)phenols | Miscellaneous (poly)phenols | Miscellaneous | Epirosmanol | 0 ± 0 |
| (Poly)phenols | Miscellaneous (poly)phenols | Miscellaneous | Rosmadial | 0 ± 0 |
| (Poly)phenols | Miscellaneous (poly)phenols | Miscellaneous | Rosmanol | 0.1 ± 0.1 |
| (Poly)phenols | Miscellaneous (poly)phenols | Miscellaneous | 3,4-DHPEA-AC | 0.1 ± 0.1 |
| (Poly)phenols | Miscellaneous (poly)phenols | Miscellaneous | 3,4-DHPEA-EA | 0.4 ± 0.4 |
| (Poly)phenols | Miscellaneous (poly)phenols | Miscellaneous | 3,4-DHPEA-EDA | 1.3 ± 0.9 |
| (Poly)phenols | Miscellaneous (poly)phenols | Miscellaneous | Demethyloleuropein | 0 ± 0 |
| (Poly)phenols | Miscellaneous (poly)phenols | Miscellaneous | Hydroxytyrosol | 0.2 ± 0.1 |
| (Poly)phenols | Miscellaneous (poly)phenols | Miscellaneous | Hydroxytyrosol 4-*O*-glucoside | 0 ± 0 |
| (Poly)phenols | Miscellaneous (poly)phenols | Miscellaneous | Ligstroside | 0.1 ± 0 |
| (Poly)phenols | Miscellaneous (poly)phenols | Miscellaneous | Ligstroside-aglycone | 0.4 ± 0.5 |
| (Poly)phenols | Miscellaneous (poly)phenols | Miscellaneous | Oleoside 11-methylester | 0 ± 0 |
| (Poly)phenols | Miscellaneous (poly)phenols | Miscellaneous | Oleoside dimethylester | 0 ± 0 |
| (Poly)phenols | Miscellaneous (poly)phenols | Miscellaneous | Oleuropein | 0 ± 0 |
| (Poly)phenols | Miscellaneous (poly)phenols | Miscellaneous | Oleuropein-aglycone | 0.8 ± 1 |
| (Poly)phenols | Miscellaneous (poly)phenols | Miscellaneous | *p*-HPEA-AC | 0 ± 0 |
| (Poly)phenols | Miscellaneous (poly)phenols | Miscellaneous | *p*-HPEA-EA | 0.3 ± 0.2 |
| (Poly)phenols | Miscellaneous (poly)phenols | Miscellaneous | *p*-HPEA-EDA | 0.8 ± 0.6 |
| (Poly)phenols | Miscellaneous (poly)phenols | Miscellaneous | Tyrosol | 1.1 ± 0.6 |

**Table S2: average intake of terpenoid family, classes, subclasses and individual compounds in the European adult population.**

| Family | Class | Subclass | Compound | Intake (mg/day) Mean ± SD |
|---|---|---|---|---|
| Terpenoids | Total | - | - | 2061.2 ± 557.4 |
| Terpenoids | Tetraterpenoids | Total | - | 46.2 ± 17.8 |
| Terpenoids | Tetraterpenoids | Carotenes | Total | 11.9 ± 2.4 |
| Terpenoids | Tetraterpenoids | Carotenes | β-carotene | 3.2 ± 0.7 |
| Terpenoids | Tetraterpenoids | Carotenes | γ-carotene | 0 ± 0 |
| Terpenoids | Tetraterpenoids | Carotenes | ζ-carotene | 0.1 ± 0 |
| Terpenoids | Tetraterpenoids | Carotenes | α-carotene | 0.6 ± 0.3 |
| Terpenoids | Tetraterpenoids | Carotenes | Lycopene | 4.2 ± 1 |
| Terpenoids | Tetraterpenoids | Carotenes | Phytoene | 2.2 ± 0.6 |
| Terpenoids | Tetraterpenoids | Carotenes | Phytofluene | 1.5 ± 0.6 |
| Terpenoids | Tetraterpenoids | Xanthophylls | Total | 34.3 ± 18 |
| Terpenoids | Tetraterpenoids | Xanthophylls | Antheraxanthin | 0.9 ± 0.5 |
| Terpenoids | Tetraterpenoids | Xanthophylls | Auroxanthin | 0.3 ± 0.2 |
| Terpenoids | Tetraterpenoids | Xanthophylls | Capsanthin | 13.5 ± 8.2 |
| Terpenoids | Tetraterpenoids | Xanthophylls | Capsanthin-3,6-epoxide | 1.6 ± 0.9 |
| Terpenoids | Tetraterpenoids | Xanthophylls | Capsanthin-5,6-epoxide | 1.7 ± 1.1 |
| Terpenoids | Tetraterpenoids | Xanthophylls | Capsanthone | 0.4 ± 0.3 |
| Terpenoids | Tetraterpenoids | Xanthophylls | Capsorubin | 2.5 ± 1.5 |
| Terpenoids | Tetraterpenoids | Xanthophylls | Cryptocapsin | 0.4 ± 0.2 |
| Terpenoids | Tetraterpenoids | Xanthophylls | α-cryptoxanthin | 0.1 ± 0.1 |
| Terpenoids | Tetraterpenoids | Xanthophylls | β-cryptoxanthin | 0.2 ± 0.1 |
| Terpenoids | Tetraterpenoids | Xanthophylls | Cucurbitaxanthin | 3.9 ± 2.4 |
| Terpenoids | Tetraterpenoids | Xanthophylls | Karpoxanthin | 0.7 ± 0.4 |
| Terpenoids | Tetraterpenoids | Xanthophylls | Lactucaxanthin | 0 ± 0 |
| Terpenoids | Tetraterpenoids | Xanthophylls | Latoxanthin | 0.1 ± 0.1 |
| Terpenoids | Tetraterpenoids | Xanthophylls | Lutein | 2.3 ± 0.9 |
| Terpenoids | Tetraterpenoids | Xanthophylls | Zeaxanthin | 2.8 ± 1.6 |
| Terpenoids | Tetraterpenoids | Xanthophylls | Neoxanthin | 0.4 ± 0.1 |
| Terpenoids | Tetraterpenoids | Xanthophylls | Mutatoxanthin | 0 ± 0 |
| Terpenoids | Tetraterpenoids | Xanthophylls | Violaxanthin | 1.6 ± 0.4 |
| Terpenoids | Tetraterpenoids | Xanthophylls | Cycloviolaxanthin | 0.7 ± 0.4 |
| Terpenoids | Tetraterpenoids | Xanthophylls | Zeinoxanthin | 0 ± 0 |
| Terpenoids | Tetraterpenoids | Xanthophylls | Luteoxanthin | 0.1 ± 0 |
| Terpenoids | Triterpenoids | Total | - | 553 ± 306.1 |
| Terpenoids | Triterpenoids | Phytosterols and phytostanols | Total | 73.6 ± 34.7 |
| Terpenoids | Triterpenoids | Phytosterols and phytostanols | Avenasterol | 7.9 ± 1.8 |
| Terpenoids | Triterpenoids | Phytosterols and phytostanols | Brassicasterol | 14.2 ± 6 |
| Terpenoids | Triterpenoids | Phytosterols and phytostanols | Campesterol | 35.4 ± 6.8 |

| Family | Class | Subclass | Compound | Intake (mg/day) Mean ± SD |
|---|---|---|---|---|
| Terpenoids | Triterpenoids | Phytosterols and phytostanols | Citrostadienol | 2 ± 1.4 |
| Terpenoids | Triterpenoids | Phytosterols and phytostanols | Clerosterol | 0.1 ± 0.1 |
| Terpenoids | Triterpenoids | Phytosterols and phytostanols | Cycloartenol | 7.4 ± 1.7 |
| Terpenoids | Triterpenoids | Phytosterols and phytostanols | Daucosterol | 0 ± 0 |
| Terpenoids | Triterpenoids | Phytosterols and phytostanols | Ergosterol | 0.7 ± 0.2 |
| Terpenoids | Triterpenoids | Phytosterols and phytostanols | 7-Ergostaenol | 0 ± 0 |
| Terpenoids | Triterpenoids | Phytosterols and phytostanols | 5,7-Ergostadienol | 0 ± 0 |
| Terpenoids | Triterpenoids | Phytosterols and phytostanols | 7,22-Ergostadienol | 0 ± 0 |
| Terpenoids | Triterpenoids | Phytosterols and phytostanols | Indosterol | 0.2 ± 0.2 |
| Terpenoids | Triterpenoids | Phytosterols and phytostanols | Sitosterol | 162.1 ± 18.6 |
| Terpenoids | Triterpenoids | Phytosterols and phytostanols | Sitosterol-glucoside | 7.1 ± 3.1 |
| Terpenoids | Triterpenoids | Phytosterols and phytostanols | α-Spinasterol | 16.4 ± 4.2 |
| Terpenoids | Triterpenoids | Phytosterols and phytostanols | Stigmastadiene | 0 ± 0 |
| Terpenoids | Triterpenoids | Phytosterols and phytostanols | Stigmasterol | 18.5 ± 3.2 |
| Terpenoids | Triterpenoids | Phytosterols and phytostanols | δ-7-Stigmastenol | 2.3 ± 1.3 |
| Terpenoids | Triterpenoids | Phytosterols and phytostanols | δ-5,23-Stigmastadienol | 0 ± 0 |
| Terpenoids | Triterpenoids | Phytosterols and phytostanols | δ-5,24-Stigmastadienol | 0.4 ± 0.3 |
| Terpenoids | Triterpenoids | Phytosterols and phytostanols | 7,22,25-Stigmastatrienol | 0 ± 0.1 |
| Terpenoids | Triterpenoids | Phytosterols and phytostanols | Campestanol | 7.7 ± 1.3 |
| Terpenoids | Triterpenoids | Phytosterols and phytostanols | Cholestanol | 0 ± 0 |
| Terpenoids | Triterpenoids | Phytosterols and phytostanols | Cycloartanol | 5.3 ± 1.5 |
| Terpenoids | Triterpenoids | Phytosterols and phytostanols | 24-Methylenecycloartanol | 9.4 ± 4.5 |
| Terpenoids | Triterpenoids | Phytosterols and phytostanols | Sitostanol | 8.9 ± 1.8 |
| Terpenoids | Triterpenoids | Saponins | Total | 123.7 ± 51.6 |
| Terpenoids | Triterpenoids | Saponins | Avenacin C | 0 ± 0 |
| Terpenoids | Triterpenoids | Saponins | Avenacin D | 0 ± 0 |
| Terpenoids | Triterpenoids | Saponins | Avenacin E | 0 ± 0 |

| Family | Class | Subclass | Compound | Intake (mg/day) Mean ± SD |
|---|---|---|---|---|
| Terpenoids | Triterpenoids | Saponins | Avenacin F | 0 ± 0 |
| Terpenoids | Triterpenoids | Saponins | Avenacin G | 0 ± 0 |
| Terpenoids | Triterpenoids | Saponins | Avenacin H | 0 ± 0 |
| Terpenoids | Triterpenoids | Saponins | Avenacin I | 0 ± 0 |
| Terpenoids | Triterpenoids | Saponins | Avenacin J | 0 ± 0 |
| Terpenoids | Triterpenoids | Saponins | Avenacin K | 0 ± 0 |
| Terpenoids | Triterpenoids | Saponins | Avenacin A1 | 0 ± 0 |
| Terpenoids | Triterpenoids | Saponins | Avenacin A2 | 0 ± 0 |
| Terpenoids | Triterpenoids | Saponins | Avenacin B1 | 0 ± 0 |
| Terpenoids | Triterpenoids | Saponins | Avenacin B2 | 0 ± 0 |
| Terpenoids | Triterpenoids | Saponins | Avenacoside A | 0.3 ± 0.3 |
| Terpenoids | Triterpenoids | Saponins | Avenacoside B | 0.1 ± 0.1 |
| Terpenoids | Triterpenoids | Saponins | Apioglycyrrhizin | 0 ± 0 |
| Terpenoids | Triterpenoids | Saponins | β-glycyrrhetinic acid | 0 ± 0.1 |
| Terpenoids | Triterpenoids | Saponins | Corosolic acid | 2.5 ± 1.4 |
| Terpenoids | Triterpenoids | Saponins | Diosgenin | 93.4 ± 50.1 |
| Terpenoids | Triterpenoids | Saponins | Chlorogenin | 0.1 ± 0 |
| Terpenoids | Triterpenoids | Saponins | Tigogenin | 0 ± 0 |
| Terpenoids | Triterpenoids | Saponins | Erythrodiol | 0 ± 0 |
| Terpenoids | Triterpenoids | Saponins | Ginsenoside RA | 0 ± 0 |
| Terpenoids | Triterpenoids | Saponins | Ginsenoside RB | 0 ± 0.1 |
| Terpenoids | Triterpenoids | Saponins | Ginsenoside RC | 0 ± 0 |
| Terpenoids | Triterpenoids | Saponins | Ginsenoside RD | 0 ± 0 |
| Terpenoids | Triterpenoids | Saponins | Ginsenoside RE | 0 ± 0.1 |
| Terpenoids | Triterpenoids | Saponins | Ginsenoside RF | 0 ± 0 |
| Terpenoids | Triterpenoids | Saponins | Ginsenoside RG | 0 ± 0 |
| Terpenoids | Triterpenoids | Saponins | Ginsenoside RH1 | 0 ± 0 |
| Terpenoids | Triterpenoids | Saponins | Ginsenoside RO | 0 ± 0 |
| Terpenoids | Triterpenoids | Saponins | α-Glycyrrhetinic acid | 0 ± 0 |
| Terpenoids | Triterpenoids | Saponins | Glycyrrhizin | 0.5 ± 1.1 |
| Terpenoids | Triterpenoids | Saponins | Hederagenin | 5.2 ± 1.7 |
| Terpenoids | Triterpenoids | Saponins | Maslinic acid | 6.7 ± 2.4 |
| Terpenoids | Triterpenoids | Saponins | Notoginsenoside R-1 | 0 ± 0 |
| Terpenoids | Triterpenoids | Saponins | Oleanolic acid | 8.5 ± 2.9 |
| Terpenoids | Triterpenoids | Saponins | Panaxadiol | 0 ± 0 |
| Terpenoids | Triterpenoids | Saponins | Panaxatriol | 0 ± 0 |
| Terpenoids | Triterpenoids | Saponins | Phytolaccagenic acid | 0 ± 0 |
| Terpenoids | Triterpenoids | Saponins | Deoxyphytolaccagenic acid | 0 ± 0 |
| Terpenoids | Triterpenoids | Saponins | Quinquenoside R-1 | 0 ± 0 |
| Terpenoids | Triterpenoids | Saponins | Serjanic acid | 0 ± 0 |
| Terpenoids | Triterpenoids | Saponins | Soyasaponin A | 0 ± 0 |
| Terpenoids | Triterpenoids | Saponins | Soyasaponin Ag | 0 ± 0 |
| Terpenoids | Triterpenoids | Saponins | Soyasaponin I | 3.4 ± 4 |
| Terpenoids | Triterpenoids | Saponins | Soyasaponin II | 0.3 ± 0.3 |

| Family | Class | Subclass | Compound | Intake (mg/day) Mean ± SD |
|---|---|---|---|---|
| Terpenoids | Triterpenoids | Saponins | Soyasaponin III | 0 ± 0 |
| Terpenoids | Triterpenoids | Saponins | Soyasaponin IV | 0 ± 0 |
| Terpenoids | Triterpenoids | Saponins | Soyasaponin V | 0.1 ± 0.1 |
| Terpenoids | Triterpenoids | Saponins | Soyasaponin VI | 1.3 ± 1.2 |
| Terpenoids | Triterpenoids | Saponins | Soyasapogenol A | 0 ± 0 |
| Terpenoids | Triterpenoids | Saponins | Soyasapogenol B | 1.3 ± 1.5 |
| Terpenoids | Triterpenoids | Saponins | Uralsaponin B | 0 ± 0 |
| Terpenoids | Triterpenoids | Saponins | Uvaol | 0 ± 0 |
| Terpenoids | Triterpenoids | Limonoids | Total | 1.4 ± 0.5 |
| Terpenoids | Triterpenoids | Limonoids | Deacetyl nomilin glucoside | 0 ± 0 |
| Terpenoids | Triterpenoids | Limonoids | Deacetyl nomilinic acid glucoside | 0 ± 0 |
| Terpenoids | Triterpenoids | Limonoids | Ichangin | 0 ± 0 |
| Terpenoids | Triterpenoids | Limonoids | Limonin | 0.3 ± 0.1 |
| Terpenoids | Triterpenoids | Limonoids | Limonin glucoside | 0 ± 0 |
| Terpenoids | Triterpenoids | Limonoids | Limonin-*d*-glycopyranoside | 0.2 ± 0.2 |
| Terpenoids | Triterpenoids | Limonoids | Limonoate A-ring lactone | 0.6 ± 0.3 |
| Terpenoids | Triterpenoids | Limonoids | Nomilin | 0.1 ± 0 |
| Terpenoids | Triterpenoids | Limonoids | Nomilin glucoside | 0 ± 0 |
| Terpenoids | Triterpenoids | Limonoids | Nomilinic acid glucoside | 0 ± 0 |
| Terpenoids | Triterpenoids | Limonoids | Nomilinoate A-ring lactone | 0.1 ± 0 |
| Terpenoids | Triterpenoids | Limonoids | Obacunone | 0.2 ± 0.1 |
| Terpenoids | Triterpenoids | Limonoids | Obacunone glucoside | 0 ± 0 |
| Terpenoids | Triterpenoids | Limonoids | Obacunoic acid | 0 ± 0 |
| Terpenoids | Triterpenoids | Ursanes | Total | 18.1 ± 9.9 |
| Terpenoids | Triterpenoids | Ursanes | Alphitolic acid | 0 ± 0 |
| Terpenoids | Triterpenoids | Ursanes | α-Amyrin | 0.5 ± 0.3 |
| Terpenoids | Triterpenoids | Ursanes | β-Amyrin | 0.1 ± 0.1 |
| Terpenoids | Triterpenoids | Ursanes | Ursolic acid | 17.4 ± 9.9 |
| Terpenoids | Triterpenoids | Ursanes | 2α-Hydroxyursolic acid | 0 ± 0 |
| Terpenoids | Triterpenoids | Ursanes | Ursonic acid | 0 ± 0 |
| Terpenoids | Triterpenoids | Miscellaneous triterpenoids | Total | 103.8 ± 53.8 |
| Terpenoids | Triterpenoids | Miscellaneous triterpenoids | Cucurbitacin E | 6.3 ± 8.4 |
| Terpenoids | Triterpenoids | Miscellaneous triterpenoids | Cucurbitacin E Glucoside | 10 ± 10.1 |
| Terpenoids | Triterpenoids | Miscellaneous triterpenoids | Cucurbitacin I | 7.6 ± 7.8 |
| Terpenoids | Triterpenoids | Miscellaneous triterpenoids | Cucurbitacin I glucoside | 0.1 ± 0.1 |
| Terpenoids | Triterpenoids | Miscellaneous triterpenoids | 22-deoxocucurbitoside B | 0.2 ± 0.1 |
| Terpenoids | Triterpenoids | Miscellaneous triterpenoids | α-Boswellic acid | 0.9 ± 0.5 |
| Terpenoids | Triterpenoids | Miscellaneous triterpenoids | Betulin | 23.9 ± 7.1 |

| Family | Class | Subclass | Compound | Intake (mg/day) Mean ± SD |
|---|---|---|---|---|
| Terpenoids | Triterpenoids | Miscellaneous triterpenoids | Betulinic acid | 1.6 ± 0.5 |
| Terpenoids | Triterpenoids | Miscellaneous triterpenoids | Betulonic acid | 0 ± 0 |
| Terpenoids | Triterpenoids | Miscellaneous triterpenoids | Ceanothic acid | 0 ± 0 |
| Terpenoids | Triterpenoids | Miscellaneous triterpenoids | Lupeol | 0.7 ± 0.2 |
| Terpenoids | Triterpenoids | Miscellaneous triterpenoids | Pomolic acid | 1.4 ± 0.7 |
| Terpenoids | Triterpenoids | Miscellaneous triterpenoids | Squalene | 50.7 ± 41.8 |
| Terpenoids | Triterpenoids | Miscellaneous triterpenoids | Tormentic acid | 0.4 ± 0.2 |
| Terpenoids | Diterpenoids | Diterpenoids | Total | 14.8 ± 15 |
| Terpenoids | Diterpenoids | Diterpenoids | Cafestol | 6.4 ± 12.8 |
| Terpenoids | Diterpenoids | Diterpenoids | Kahweol | 3 ± 2.7 |
| Terpenoids | Diterpenoids | Diterpenoids | Capsianoside A | 1.5 ± 0.9 |
| Terpenoids | Diterpenoids | Diterpenoids | Capsianoside B | 0.1 ± 0.1 |
| Terpenoids | Diterpenoids | Diterpenoids | Capsianoside C | 0.7 ± 0.5 |
| Terpenoids | Diterpenoids | Diterpenoids | Capsianoside D | 0.3 ± 0.2 |
| Terpenoids | Diterpenoids | Diterpenoids | Capsianoside E | 0.2 ± 0.1 |
| Terpenoids | Diterpenoids | Diterpenoids | Capsianoside F | 0.1 ± 0 |
| Terpenoids | Diterpenoids | Diterpenoids | Capsianoside I | 0.2 ± 0.1 |
| Terpenoids | Diterpenoids | Diterpenoids | Capsianoside II | 1 ± 0.6 |
| Terpenoids | Diterpenoids | Diterpenoids | Capsianoside III | 0.6 ± 0.4 |
| Terpenoids | Diterpenoids | Diterpenoids | Capsianoside IV | 0.1 ± 0.1 |
| Terpenoids | Diterpenoids | Diterpenoids | Capsianoside V | 0 ± 0 |
| Terpenoids | Diterpenoids | Diterpenoids | Cinnzeylanine | 0 ± 0 |
| Terpenoids | Diterpenoids | Diterpenoids | Cinnzeylanol | 0 ± 0 |
| Terpenoids | Diterpenoids | Diterpenoids | Phytol | 0.2 ± 0.1 |
| Terpenoids | Diterpenoids | Diterpenoids | 16-*O*-methylcafestol | 0.4 ± 0.5 |
| Terpenoids | Sesquiterpenoids | Sesquiterpenoids | Total | 103 ± 49.5 |
| Terpenoids | Sesquiterpenoids | Sesquiterpenoids | Aromadendrene | 0 ± 0 |
| Terpenoids | Sesquiterpenoids | Sesquiterpenoids | Aromadendrine | 0 ± 0 |
| Terpenoids | Sesquiterpenoids | Sesquiterpenoids | α-bergamotene | 34.2 ± 17.6 |
| Terpenoids | Sesquiterpenoids | Sesquiterpenoids | α-bisabolene | 2.1 ± 1.8 |
| Terpenoids | Sesquiterpenoids | Sesquiterpenoids | α-bulnesene | 0 ± 0 |
| Terpenoids | Sesquiterpenoids | Sesquiterpenoids | α-cadinene | 0 ± 0 |
| Terpenoids | Sesquiterpenoids | Sesquiterpenoids | α-cadinol | 0 ± 0 |
| Terpenoids | Sesquiterpenoids | Sesquiterpenoids | α-cubebene | 0.1 ± 0 |
| Terpenoids | Sesquiterpenoids | Sesquiterpenoids | α-farnesene | 0 ± 0 |
| Terpenoids | Sesquiterpenoids | Sesquiterpenoids | α-gurjunene | 0 ± 0 |
| Terpenoids | Sesquiterpenoids | Sesquiterpenoids | α-muurolene | 0 ± 0 |
| Terpenoids | Sesquiterpenoids | Sesquiterpenoids | α-santalene | 0 ± 0 |
| Terpenoids | Sesquiterpenoids | Sesquiterpenoids | α-selinene | 0 ± 0.1 |

| Family | Class | Subclass | Compound | Intake (mg/day) Mean ± SD |
|---|---|---|---|---|
| Terpenoids | Sesquiterpenoids | Sesquiterpenoids | α-sinensal | 0.1 ± 0.1 |
| Terpenoids | Sesquiterpenoids | Sesquiterpenoids | α-ylangene | 0 ± 0 |
| Terpenoids | Sesquiterpenoids | Sesquiterpenoids | Curcumene | 0.3 ± 0.3 |
| Terpenoids | Sesquiterpenoids | Sesquiterpenoids | Artemorin | 0 ± 0.1 |
| Terpenoids | Sesquiterpenoids | Sesquiterpenoids | β-bisabolene | 2.6 ± 1.1 |
| Terpenoids | Sesquiterpenoids | Sesquiterpenoids | β-bisabolol | 0 ± 0 |
| Terpenoids | Sesquiterpenoids | Sesquiterpenoids | β-cubebene | 0.8 ± 0.4 |
| Terpenoids | Sesquiterpenoids | Sesquiterpenoids | β-eudesmol | 0 ± 0 |
| Terpenoids | Sesquiterpenoids | Sesquiterpenoids | β-gurjunene | 0 ± 0 |
| Terpenoids | Sesquiterpenoids | Sesquiterpenoids | β-selinene | 0 ± 0 |
| Terpenoids | Sesquiterpenoids | Sesquiterpenoids | β-sesquiphellandrene | 0.2 ± 0.1 |
| Terpenoids | Sesquiterpenoids | Sesquiterpenoids | Calamenene | 0 ± 0 |
| Terpenoids | Sesquiterpenoids | Sesquiterpenoids | Capsidiol | 0.3 ± 0.2 |
| Terpenoids | Sesquiterpenoids | Sesquiterpenoids | Caryophyllene | 4.9 ± 2 |
| Terpenoids | Sesquiterpenoids | Sesquiterpenoids | Copaene | 0.1 ± 0 |
| Terpenoids | Sesquiterpenoids | Sesquiterpenoids | Costunolide | 0.2 ± 0.3 |
| Terpenoids | Sesquiterpenoids | Sesquiterpenoids | δ-cadinene | 0.2 ± 0.1 |
| Terpenoids | Sesquiterpenoids | Sesquiterpenoids | Elemene | 0.3 ± 0.1 |
| Terpenoids | Sesquiterpenoids | Sesquiterpenoids | Elemol | 0 ± 0 |
| Terpenoids | Sesquiterpenoids | Sesquiterpenoids | Eremophilene | 0 ± 0 |
| Terpenoids | Sesquiterpenoids | Sesquiterpenoids | β-farnesene | 0.2 ± 0.1 |
| Terpenoids | Sesquiterpenoids | Sesquiterpenoids | Farnesal | 0 ± 0 |
| Terpenoids | Sesquiterpenoids | Sesquiterpenoids | Farnesol | 0 ± 0 |
| Terpenoids | Sesquiterpenoids | Sesquiterpenoids | γ-bisabolene | 4.3 ± 2.3 |
| Terpenoids | Sesquiterpenoids | Sesquiterpenoids | γ-cadinene | 0.1 ± 0.1 |
| Terpenoids | Sesquiterpenoids | Sesquiterpenoids | γ-eudesmol | 0 ± 0 |
| Terpenoids | Sesquiterpenoids | Sesquiterpenoids | γ-muurolene | 49.2 ± 26.4 |
| Terpenoids | Sesquiterpenoids | Sesquiterpenoids | γ-muurolol | 0 ± 0 |
| Terpenoids | Sesquiterpenoids | Sesquiterpenoids | γ-selinene | 0.2 ± 0.1 |
| Terpenoids | Sesquiterpenoids | Sesquiterpenoids | Germacrene-D | 0 ± 0 |
| Terpenoids | Sesquiterpenoids | Sesquiterpenoids | 6-Gingediol | 0 ± 0 |
| Terpenoids | Sesquiterpenoids | Sesquiterpenoids | Ginsenol | 0 ± 0 |
| Terpenoids | Sesquiterpenoids | Sesquiterpenoids | Humulene | 1.2 ± 1.3 |
| Terpenoids | Sesquiterpenoids | Sesquiterpenoids | Nerolidol | 0 ± 0 |
| Terpenoids | Sesquiterpenoids | Sesquiterpenoids | Reynosin | 0 ± 0 |
| Terpenoids | Sesquiterpenoids | Sesquiterpenoids | Santamarin | 0 ± 0 |
| Terpenoids | Sesquiterpenoids | Sesquiterpenoids | Verlotorin | 0 ± 0 |
| Terpenoids | Sesquiterpenoids | Sesquiterpenoids | Zingiberene | 0 ± 0 |
| Terpenoids | Sesquiterpenoids | Sesquiterpenoids | Zingiberenol | 0 ± 0 |
| Terpenoids | Sesquiterpenoids | Sesquiterpenoids | Zingiberol | 0.4 ± 0.4 |
| Terpenoids | Sesquiterpenoids | Sesquiterpenoids | Lactucin | 0 ± 0 |
| Terpenoids | Sesquiterpenoids | Sesquiterpenoids | 113,13-Dihydrolactucin | 0 ± 0 |
| Terpenoids | Sesquiterpenoids | Sesquiterpenoids | β-ionone | 0 ± 0 |
| Terpenoids | Sesquiterpenoids | Sesquiterpenoids | Bicycloelemene | 0 ± 0 |

| Family | Class | Subclass | Compound | Intake (mg/day) Mean ± SD |
|---|---|---|---|---|
| Terpenoids | Sesquiterpenoids | Sesquiterpenoids | Bicyclogermacrene | 0 ± 0 |
| Terpenoids | Sesquiterpenoids | Sesquiterpenoids | Deoxylactucin | 0.5 ± 0.4 |
| Terpenoids | Sesquiterpenoids | Sesquiterpenoids | Dihydrolactucopicrin | 0 ± 0 |
| Terpenoids | Sesquiterpenoids | Sesquiterpenoids | Lactuside C | 0 ± 0 |
| Terpenoids | Sesquiterpenoids | Sesquiterpenoids | Lactucopicrin | 0.1 ± 0.2 |
| Terpenoids | Sesquiterpenoids | Sesquiterpenoids | Valencene | 0 ± 0 |
| Terpenoids | Monoterpenoids | Monoterpenoids | Total | 1344.1 ± 518.1 |
| Terpenoids | Monoterpenoids | Monoterpenoids | α-Terpinene | 1.3 ± 0.5 |
| Terpenoids | Monoterpenoids | Monoterpenoids | α-Terpineol | 2.7 ± 1.7 |
| Terpenoids | Monoterpenoids | Monoterpenoids | α-Thujene | 0.3 ± 0.2 |
| Terpenoids | Monoterpenoids | Monoterpenoids | α-Thujone | 0.2 ± 0.3 |
| Terpenoids | Monoterpenoids | Monoterpenoids | β-Terpinene | 0 ± 0 |
| Terpenoids | Monoterpenoids | Monoterpenoids | β-Terpineol | 0.8 ± 0.4 |
| Terpenoids | Monoterpenoids | Monoterpenoids | β-Thujone | 0.2 ± 0.3 |
| Terpenoids | Monoterpenoids | Monoterpenoids | γ-Terpinene | 11.2 ± 6.1 |
| Terpenoids | Monoterpenoids | Monoterpenoids | δ-3-Carene | 0.9 ± 0.3 |
| Terpenoids | Monoterpenoids | Monoterpenoids | Camphene | 1.2 ± 0.7 |
| Terpenoids | Monoterpenoids | Monoterpenoids | Carvacrol | 0 ± 0.1 |
| Terpenoids | Monoterpenoids | Monoterpenoids | Carvone | 0.9 ± 0.6 |
| Terpenoids | Monoterpenoids | Monoterpenoids | Geranial | 7.7 ± 3.1 |
| Terpenoids | Monoterpenoids | Monoterpenoids | Limonene | 830 ± 335.7 |
| Terpenoids | Monoterpenoids | Monoterpenoids | Linalool | 25.2 ± 10 |
| Terpenoids | Monoterpenoids | Monoterpenoids | Myrcene | 17.8 ± 7.4 |
| Terpenoids | Monoterpenoids | Monoterpenoids | *p*-Cymene | 312.5 ± 138.7 |
| Terpenoids | Monoterpenoids | Monoterpenoids | Sabinene | 5.9 ± 2 |
| Terpenoids | Monoterpenoids | Monoterpenoids | Thymol | 0 ± 0 |
| Terpenoids | Monoterpenoids | Monoterpenoids | Dihydrocarvone | 0.1 ± 0.1 |
| Terpenoids | Monoterpenoids | Monoterpenoids | Sabinene hydrate | 0.1 ± 0 |
| Terpenoids | Monoterpenoids | Monoterpenoids | 1,4-Cineole | 1.2 ± 1 |
| Terpenoids | Monoterpenoids | Monoterpenoids | 1,8-Cineole | 1.4 ± 0.8 |
| Terpenoids | Monoterpenoids | Monoterpenoids | Borneol | 0.4 ± 0.3 |
| Terpenoids | Monoterpenoids | Monoterpenoids | Camphor | 0 ± 0 |
| Terpenoids | Monoterpenoids | Monoterpenoids | Carvene | 0 ± 0 |
| Terpenoids | Monoterpenoids | Monoterpenoids | Carveol | 0.1 ± 0.1 |
| Terpenoids | Monoterpenoids | Monoterpenoids | Myrtenol | 0 ± 0 |
| Terpenoids | Monoterpenoids | Monoterpenoids | Citronellal | 1.3 ± 0.8 |
| Terpenoids | Monoterpenoids | Monoterpenoids | Citronellol | 0.3 ± 0.2 |
| Terpenoids | Monoterpenoids | Monoterpenoids | Dihydrocarveol | 0 ± 0.1 |
| Terpenoids | Monoterpenoids | Monoterpenoids | Fenchol | 0.8 ± 0.6 |
| Terpenoids | Monoterpenoids | Monoterpenoids | Fenchone | 0 ± 0 |
| Terpenoids | Monoterpenoids | Monoterpenoids | Geraniol | 66.8 ± 35.7 |
| Terpenoids | Monoterpenoids | Monoterpenoids | Isomenthol | 0 ± 0.1 |
| Terpenoids | Monoterpenoids | Monoterpenoids | Isomenthone | 0.2 ± 0.4 |
| Terpenoids | Monoterpenoids | Monoterpenoids | Isothymol | 0 ± 0 |

| Family | Class | Subclass | Compound | Intake (mg/day) Mean ± SD |
|---|---|---|---|---|
| Terpenoids | Monoterpenoids | Monoterpenoids | Menthol | 1 ± 2.6 |
| Terpenoids | Monoterpenoids | Monoterpenoids | Myrtenal | 0 ± 0 |
| Terpenoids | Monoterpenoids | Monoterpenoids | Neomenthol | 0.1 ± 0.3 |
| Terpenoids | Monoterpenoids | Monoterpenoids | Neomenthone | 0.1 ± 0.3 |
| Terpenoids | Monoterpenoids | Monoterpenoids | Neral | 3.5 ± 1.4 |
| Terpenoids | Monoterpenoids | Monoterpenoids | Nerol | 0.1 ± 0 |
| Terpenoids | Monoterpenoids | Monoterpenoids | Ocimene | 0.1 ± 0 |
| Terpenoids | Monoterpenoids | Monoterpenoids | Perillene | 0 ± 0 |
| Terpenoids | Monoterpenoids | Monoterpenoids | Phellandrene | 2.7 ± 1.2 |
| Terpenoids | Monoterpenoids | Monoterpenoids | Pinene | 12 ± 5.4 |
| Terpenoids | Monoterpenoids | Monoterpenoids | Pinocarvone | 0 ± 0 |
| Terpenoids | Monoterpenoids | Monoterpenoids | Piperitenone | 0 ± 0 |
| Terpenoids | Monoterpenoids | Monoterpenoids | Piperitone | 0.2 ± 0.4 |
| Terpenoids | Monoterpenoids | Monoterpenoids | Sabinol | 0 ± 0 |
| Terpenoids | Monoterpenoids | Monoterpenoids | Terpinolene | 25.8 ± 13.2 |

**Table S3: average intake of *N*-containing compound family, classes, subclasses and individual compounds in the European adult population.**

| Family | Class | Subclass | Compound | Intake (mg/day) Mean ± SD |
|---|---|---|---|---|
| *N*-containing compounds | Total | - | - | 1092.1 ± 392.1 |
| *N*-containing compounds | Alkaloids | Total | - | 582.4 ± 283.3 |
| *N*-containing compounds | Alkaloids | Purines and pyrimidines | Total | 344.4 ± 169.9 |
| *N*-containing compounds | Alkaloids | Purines and pyrimidines | Convicine | 0.3 ± 0.6 |
| *N*-containing compounds | Alkaloids | Purines and pyrimidines | *N*-Methylpyridinium | 2.9 ± 2.1 |
| *N*-containing compounds | Alkaloids | Purines and pyrimidines | Theophylline | 0.1 ± 0.3 |
| *N*-containing compounds | Alkaloids | Purines and pyrimidines | Vicine | 0.8 ± 1.6 |
| *N*-containing compounds | Alkaloids | Purines and pyrimidines | Caffeine | 289.8 ± 165.9 |
| *N*-containing compounds | Alkaloids | Purines and pyrimidines | Theobromine | 50.5 ± 19.1 |
| *N*-containing compounds | Alkaloids | Betalains | Total | 4.8 ± 6.8 |
| *N*-containing compounds | Alkaloids | Betalains | Amaranthine | 0 ± 0 |
| *N*-containing compounds | Alkaloids | Betalains | iso-Amaranthine | 0 ± 0 |
| *N*-containing compounds | Alkaloids | Betalains | Betanin | 4.8 ± 6.8 |

| Family | Class | Subclass | Compound | Intake (mg/day) Mean ± SD |
|---|---|---|---|---|
| *N*-containing compounds | Alkaloids | Betalains | Indicaxanthin | 0 ± 0.1 |
| *N*-containing compounds | Alkaloids | Pyrrolidines and piperidines | Total | 16.1 ± 19.4 |
| *N*-containing compounds | Alkaloids | Pyrrolidines and piperidines | *D*-Fagomine | 0 ± 0 |
| *N*-containing compounds | Alkaloids | Pyrrolidines and piperidines | 3,4-Di-epi-fagomine | 0 ± 0 |
| *N*-containing compounds | Alkaloids | Pyrrolidines and piperidines | *N*,*N*-dimethylpiperidinium | 0 ± 0 |
| *N*-containing compounds | Alkaloids | Pyrrolidines and piperidines | Piperine | 16.1 ± 19.4 |
| *N*-containing compounds | Alkaloids | Pyrrolidines and piperidines | *N*-nitrosopyrrolidine | 0 ± 0 |
| *N*-containing compounds | Alkaloids | Pyrrolidines and piperidines | *N*-nitrosopiperidine | 0 ± 0 |
| *N*-containing compounds | Alkaloids | Pyrrolidines and piperidines | 1-(2,4-Decadienoyl)-Pyrrolidine | 0 ± 0 |
| *N*-containing compounds | Alkaloids | Pyrrolidines and piperidines | 1-(2,4-Dodecadienoyl)-Pyrrolidine | 0 ± 0 |
| *N*-containing compounds | Alkaloids | Pyrrolidines and piperidines | *N*-Trans-Feruloyl-Piperidine | 0 ± 0 |
| *N*-containing compounds | Alkaloids | Steroidal alkaloids | Total | 3.3 ± 0.9 |
| *N*-containing compounds | Alkaloids | Steroidal alkaloids | Carpaine | 0 ± 0 |
| *N*-containing compounds | Alkaloids | Steroidal alkaloids | α-Chaconine | 1.9 ± 0.5 |
| *N*-containing compounds | Alkaloids | Steroidal alkaloids | α-Solanine | 1.3 ± 0.4 |
| *N*-containing compounds | Alkaloids | Steroidal alkaloids | Tomatine | 0.2 ± 0 |
| *N*-containing compounds | Alkaloids | Steroidal alkaloids | Dehydrotomatine | 0 ± 0 |
| *N*-containing compounds | Alkaloids | Miscellaneous alkaloids | Total | 213.7 ± 109.8 |
| *N*-containing compounds | Alkaloids | Miscellaneous alkaloids | Albine | 0 ± 0 |
| *N*-containing compounds | Alkaloids | Miscellaneous alkaloids | Anisodine | 0 ± 0 |
| *N*-containing compounds | Alkaloids | Miscellaneous alkaloids | Atropine | 0 ± 0 |
| *N*-containing compounds | Alkaloids | Miscellaneous alkaloids | Apoatropine | 0 ± 0 |
| *N*-containing compounds | Alkaloids | Miscellaneous alkaloids | α-Hydroxymethyl Atropine | 0 ± 0 |
| *N*-containing compounds | Alkaloids | Miscellaneous alkaloids | Apohyoscine | 0 ± 0 |
| *N*-containing compounds | Alkaloids | Miscellaneous alkaloids | Calystegine A3 | 18 ± 5.2 |
| *N*-containing compounds | Alkaloids | Miscellaneous alkaloids | Calystegine A5 | 0.2 ± 0.1 |

| Family | Class | Subclass | Compound | Intake (mg/day) Mean ± SD |
|---|---|---|---|---|
| *N*-containing compounds | Alkaloids | Miscellaneous alkaloids | Calystegine B1 | 0.1 ± 0.1 |
| *N*-containing compounds | Alkaloids | Miscellaneous alkaloids | Calystegine B2 | 3.5 ± 0.9 |
| *N*-containing compounds | Alkaloids | Miscellaneous alkaloids | Calystegine B3 | 0.1 ± 0 |
| *N*-containing compounds | Alkaloids | Miscellaneous alkaloids | Calystegine B4 | 0.3 ± 0.1 |
| *N*-containing compounds | Alkaloids | Miscellaneous alkaloids | Convolamine | 0 ± 0 |
| *N*-containing compounds | Alkaloids | Miscellaneous alkaloids | Convolidine | 0 ± 0 |
| *N*-containing compounds | Alkaloids | Miscellaneous alkaloids | Convolvine | 0 ± 0 |
| *N*-containing compounds | Alkaloids | Miscellaneous alkaloids | Fillalbin | 0 ± 0 |
| *N*-containing compounds | Alkaloids | Miscellaneous alkaloids | Harman | 0 ± 0 |
| *N*-containing compounds | Alkaloids | Miscellaneous alkaloids | Norharman | 0.1 ± 0.1 |
| *N*-containing compounds | Alkaloids | Miscellaneous alkaloids | Homatropine | 0 ± 0 |
| *N*-containing compounds | Alkaloids | Miscellaneous alkaloids | Anisodamine | 0 ± 0 |
| *N*-containing compounds | Alkaloids | Miscellaneous alkaloids | Isoxanthopterin | 0 ± 0 |
| *N*-containing compounds | Alkaloids | Miscellaneous alkaloids | Littorine | 0 ± 0 |
| *N*-containing compounds | Alkaloids | Miscellaneous alkaloids | Lupanine | 0 ± 0 |
| *N*-containing compounds | Alkaloids | Miscellaneous alkaloids | 13a-hydroxylupanine | 0 ± 0 |
| *N*-containing compounds | Alkaloids | Miscellaneous alkaloids | Lupinine | 0 ± 0 |
| *N*-containing compounds | Alkaloids | Miscellaneous alkaloids | Noratropine | 0 ± 0 |
| *N*-containing compounds | Alkaloids | Miscellaneous alkaloids | Nortropinone | 0 ± 0 |
| *N*-containing compounds | Alkaloids | Miscellaneous alkaloids | 2-Acetyl-pyridine | 0 ± 0 |
| *N*-containing compounds | Alkaloids | Miscellaneous alkaloids | Multiflorine | 0 ± 0 |
| *N*-containing compounds | Alkaloids | Miscellaneous alkaloids | 3α-Phenylacetoxytropane | 0 ± 0 |
| *N*-containing compounds | Alkaloids | Miscellaneous alkaloids | Scopine | 0 ± 0 |
| *N*-containing compounds | Alkaloids | Miscellaneous alkaloids | Scopoline | 0 ± 0 |
| *N*-containing compounds | Alkaloids | Miscellaneous alkaloids | Scopolamine | 0 ± 0 |
| *N*-containing compounds | Alkaloids | Miscellaneous alkaloids | Scopolamine acetate | 0 ± 0 |

| Family | Class | Subclass | Compound | Intake (mg/day) Mean ± SD |
|---|---|---|---|---|
| *N*-containing compounds | Alkaloids | Miscellaneous alkaloids | Norscopolamine | 0 ± 0 |
| *N*-containing compounds | Alkaloids | Miscellaneous alkaloids | Sparteine | 0 ± 0 |
| *N*-containing compounds | Alkaloids | Miscellaneous alkaloids | Tropine | 0 ± 0 |
| *N*-containing compounds | Alkaloids | Miscellaneous alkaloids | Tropinone | 0 ± 0 |
| *N*-containing compounds | Alkaloids | Miscellaneous alkaloids | 6-Hydroxytropinone | 0 ± 0 |
| *N*-containing compounds | Alkaloids | Miscellaneous alkaloids | Trigonelline | 191.4 ± 110.5 |
| *N*-containing compounds | Alkaloids | Miscellaneous alkaloids | MTCA | 0 ± 0 |
| *N*-containing compounds | Glucosinolates and isothiocyanates | Total | - | 30.8 ± 10.3 |
| *N*-containing compounds | Glucosinolates and isothiocyanates | Glucosinolates | Total | 24.2 ± 8.2 |
| *N*-containing compounds | Glucosinolates and isothiocyanates | Glucosinolates | Glucoraphanin | 2.6 ± 1.8 |
| *N*-containing compounds | Glucosinolates and isothiocyanates | Glucosinolates | 4-hydroxyglucobrassicin | 0.4 ± 0.2 |
| *N*-containing compounds | Glucosinolates and isothiocyanates | Glucosinolates | 4-Methoxyglucobrassicin | 1.4 ± 0.8 |
| *N*-containing compounds | Glucosinolates and isothiocyanates | Glucosinolates | Glucoiberin | 1.9 ± 1 |
| *N*-containing compounds | Glucosinolates and isothiocyanates | Glucosinolates | Glucoerucin | 0.8 ± 0.4 |
| *N*-containing compounds | Glucosinolates and isothiocyanates | Glucosinolates | Gluconapin | 1.2 ± 0.9 |
| *N*-containing compounds | Glucosinolates and isothiocyanates | Glucosinolates | Progoitrin | 1.2 ± 0.6 |
| *N*-containing compounds | Glucosinolates and isothiocyanates | Glucosinolates | Glucoapoleiferin | 0 ± 0 |
| *N*-containing compounds | Glucosinolates and isothiocyanates | Glucosinolates | Progoitrin isomero S | 0.1 ± 0.1 |
| *N*-containing compounds | Glucosinolates and isothiocyanates | Glucosinolates | Glucobrassicin | 3.9 ± 1.6 |
| *N*-containing compounds | Glucosinolates and isothiocyanates | Glucosinolates | Neoglucobrassicin | 0.9 ± 0.4 |
| *N*-containing compounds | Glucosinolates and isothiocyanates | Glucosinolates | Gluconapoleiferin | 0.2 ± 0.1 |
| *N*-containing compounds | Glucosinolates and isothiocyanates | Glucosinolates | Glucoalyssin | 0.2 ± 0.1 |
| *N*-containing compounds | Glucosinolates and isothiocyanates | Glucosinolates | Glucoiberverin | 0.1 ± 0 |
| *N*-containing compounds | Glucosinolates and isothiocyanates | Glucosinolates | Glucoibervirin | 0.2 ± 0.1 |
| *N*-containing compounds | Glucosinolates and isothiocyanates | Glucosinolates | Glucoibeverin | 0 ± 0 |
| *N*-containing compounds | Glucosinolates and isothiocyanates | Glucosinolates | Glucobrassicanapin | 2.6 ± 1.5 |

| Family | Class | Subclass | Compound | Intake (mg/day) Mean ± SD |
|---|---|---|---|---|
| *N*-containing compounds | Glucosinolates and isothiocyanates | Glucosinolates | Gluconasturtiin | 0.7 ± 0.4 |
| *N*-containing compounds | Glucosinolates and isothiocyanates | Glucosinolates | Sinigrin | 1.7 ± 0.9 |
| *N*-containing compounds | Glucosinolates and isothiocyanates | Glucosinolates | Glucoraphenin | 0.1 ± 0 |
| *N*-containing compounds | Glucosinolates and isothiocyanates | Glucosinolates | Glucocapparin | 3.2 ± 3.1 |
| *N*-containing compounds | Glucosinolates and isothiocyanates | Glucosinolates | Glucoberteroin | 0 ± 0.1 |
| *N*-containing compounds | Glucosinolates and isothiocyanates | Glucosinolates | Mercaptobutylglucosinolate | 0.8 ± 0.8 |
| *N*-containing compounds | Glucosinolates and isothiocyanates | Isothiocyanates | Total | 6.6 ± 3.3 |
| *N*-containing compounds | Glucosinolates and isothiocyanates | Isothiocyanates | 3-Butenyl isothiocyanate | 0 ± 0 |
| *N*-containing compounds | Glucosinolates and isothiocyanates | Isothiocyanates | 4-methylsulphinylbutyl isothiocyanate | 0.1 ± 0 |
| *N*-containing compounds | Glucosinolates and isothiocyanates | Isothiocyanates | Isobutyl-isothiocyanate | 0 ± 0 |
| *N*-containing compounds | Glucosinolates and isothiocyanates | Isothiocyanates | Benzyl-isothiocyanate | 0 ± 0 |
| *N*-containing compounds | Glucosinolates and isothiocyanates | Isothiocyanates | Allyl isothiocyanate | 1 ± 0.9 |
| *N*-containing compounds | Glucosinolates and isothiocyanates | Isothiocyanates | Iberin | 0 ± 0 |
| *N*-containing compounds | Glucosinolates and isothiocyanates | Isothiocyanates | Phenylethyl Isothiocyanate | 0.2 ± 0.2 |
| *N*-containing compounds | Glucosinolates and isothiocyanates | Isothiocyanates | Iberverin | 0 ± 0 |
| *N*-containing compounds | Glucosinolates and isothiocyanates | Isothiocyanates | Sulforaphane | 5.3 ± 3.3 |
| *N*-containing compounds | Miscellaneous *N*-containing compounds | Total | - | 478.9 ± 178.1 |
| *N*-containing compounds | Miscellaneous *N*-containing compounds | Amines | Total | 28.6 ± 2.8 |
| *N*-containing compounds | Miscellaneous *N*-containing compounds | Amines | Agmatine | 1.1 ± 0.6 |
| *N*-containing compounds | Miscellaneous *N*-containing compounds | Amines | Cadaverine | 2.1 ± 0.4 |
| *N*-containing compounds | Miscellaneous *N*-containing compounds | Amines | Dopamine | 1.1 ± 0.4 |
| *N*-containing compounds | Miscellaneous *N*-containing compounds | Amines | Ethanolamine | 0 ± 0 |
| *N*-containing compounds | Miscellaneous *N*-containing compounds | Amines | Ethylamine | 0.2 ± 0.1 |
| *N*-containing compounds | Miscellaneous *N*-containing compounds | Amines | Histamine | 0.7 ± 0.2 |
| *N*-containing compounds | Miscellaneous *N*-containing compounds | Amines | Melatonin | 0 ± 0 |
| *N*-containing compounds | Miscellaneous *N*-containing compounds | Amines | Methylamine | 0 ± 0 |

| Family | Class | Subclass | Compound | Intake (mg/day) Mean ± SD |
|---|---|---|---|---|
| *N*-containing compounds | Miscellaneous *N*-containing compounds | Amines | Norepinephrine | 0 ± 0 |
| *N*-containing compounds | Miscellaneous *N*-containing compounds | Amines | Phenylethylamine | 0.3 ± 0.1 |
| *N*-containing compounds | Miscellaneous *N*-containing compounds | Amines | Putrescine | 10.5 ± 1.4 |
| *N*-containing compounds | Miscellaneous *N*-containing compounds | Amines | Serotonin | 1.4 ± 0.3 |
| *N*-containing compounds | Miscellaneous *N*-containing compounds | Amines | Spermidine | 7 ± 0.8 |
| *N*-containing compounds | Miscellaneous *N*-containing compounds | Amines | Spermine | 1.6 ± 0.3 |
| *N*-containing compounds | Miscellaneous *N*-containing compounds | Amines | Tryptamine | 0.2 ± 0.1 |
| *N*-containing compounds | Miscellaneous *N*-containing compounds | Amines | Tyramine | 2.2 ± 0.5 |
| *N*-containing compounds | Miscellaneous *N*-containing compounds | Non-protein amino acids | Total | 380.6 ± 166.1 |
| *N*-containing compounds | Miscellaneous *N*-containing compounds | Non-protein amino acids | Betaine | 152.8 ± 33.8 |
| *N*-containing compounds | Miscellaneous *N*-containing compounds | Non-protein amino acids | Betonicine | 4.4 ± 2.3 |
| *N*-containing compounds | Miscellaneous *N*-containing compounds | Non-protein amino acids | Citrulline | 15.9 ± 13.2 |
| *N*-containing compounds | Miscellaneous *N*-containing compounds | Non-protein amino acids | Ergothioneine | 0.1 ± 0.1 |
| *N*-containing compounds | Miscellaneous *N*-containing compounds | Non-protein amino acids | Proline betaine | 32.8 ± 11 |
| *N*-containing compounds | Miscellaneous *N*-containing compounds | Non-protein amino acids | Spinacine | 0 ± 0 |
| *N*-containing compounds | Miscellaneous *N*-containing compounds | Non-protein amino acids | Ornithine | 0 ± 0 |
| *N*-containing compounds | Miscellaneous *N*-containing compounds | Non-protein amino acids | Taurine | 2.6 ± 0.9 |
| *N*-containing compounds | Miscellaneous *N*-containing compounds | Non-protein amino acids | Theanine | 171.9 ± 159.5 |
| *N*-containing compounds | Miscellaneous *N*-containing compounds | Cyanogens | Total | 3.1 ± 1.3 |
| *N*-containing compounds | Miscellaneous *N*-containing compounds | Cyanogens | Amygdalin | 2.2 ± 1.4 |
| *N*-containing compounds | Miscellaneous *N*-containing compounds | Cyanogens | Dhurrin | 0 ± 0 |
| *N*-containing compounds | Miscellaneous *N*-containing compounds | Cyanogens | Linamarin | 0 ± 0 |
| *N*-containing compounds | Miscellaneous *N*-containing compounds | Cyanogens | Linustatin | 0.2 ± 0.2 |
| *N*-containing compounds | Miscellaneous *N*-containing compounds | Cyanogens | Lotaustralin | 0.5 ± 0.4 |
| *N*-containing compounds | Miscellaneous *N*-containing compounds | Cyanogens | Neolinustatin | 0.3 ± 0.3 |
| *N*-containing compounds | Miscellaneous *N*-containing compounds | Cyanogens | Prunasin | 0 ± 0 |

| Family | Class | Subclass | Compound | Intake (mg/day) Mean ± SD |
|---|---|---|---|---|
| *N*-containing compounds | Miscellaneous *N*-containing compounds | Miscellaneous | Total | 66.6 ± 24.3 |
| *N*-containing compounds | Miscellaneous *N*-containing compounds | Miscellaneous | Cyclo(pro-leu) | 0 ± 0 |
| *N*-containing compounds | Miscellaneous *N*-containing compounds | Miscellaneous | Cyclo(pro-phe) | 0 ± 0 |
| *N*-containing compounds | Miscellaneous *N*-containing compounds | Miscellaneous | Cyclo(pro-pro) | 0 ± 0 |
| *N*-containing compounds | Miscellaneous *N*-containing compounds | Miscellaneous | Cyclo(pro-ile) | 0 ± 0 |
| *N*-containing compounds | Miscellaneous *N*-containing compounds | Miscellaneous | Cyclo(pro-val) | 0.1 ± 0.1 |
| *N*-containing compounds | Miscellaneous *N*-containing compounds | Miscellaneous | Cyclo(phe-leu) | 0 ± 0 |
| *N*-containing compounds | Miscellaneous *N*-containing compounds | Miscellaneous | Cyclo(phe-ile) | 0 ± 0 |
| *N*-containing compounds | Miscellaneous *N*-containing compounds | Miscellaneous | Cyclo(phe-val) | 0 ± 0 |
| *N*-containing compounds | Miscellaneous *N*-containing compounds | Miscellaneous | Cyclo(phe-ala) | 0 ± 0 |
| *N*-containing compounds | Miscellaneous *N*-containing compounds | Miscellaneous | Cyclo(ile-val) | 0 ± 0 |
| *N*-containing compounds | Miscellaneous *N*-containing compounds | Miscellaneous | Cyclo(leu-val) | 0 ± 0 |
| *N*-containing compounds | Miscellaneous *N*-containing compounds | Miscellaneous | Cyclo(leu-leu) | 0 ± 0 |
| *N*-containing compounds | Miscellaneous *N*-containing compounds | Miscellaneous | Cyclo(leu-ile) | 0 ± 0 |
| *N*-containing compounds | Miscellaneous *N*-containing compounds | Miscellaneous | Cyclo(phe-phe) | 0 ± 0 |
| *N*-containing compounds | Miscellaneous *N*-containing compounds | Miscellaneous | Cyclo(ile-ala) | 0 ± 0 |
| *N*-containing compounds | Miscellaneous *N*-containing compounds | Miscellaneous | Cyclo(leu-ala) | 0 ± 0 |
| *N*-containing compounds | Miscellaneous *N*-containing compounds | Miscellaneous | Cyclo(phe-tyr) | 0 ± 0 |
| *N*-containing compounds | Miscellaneous *N*-containing compounds | Miscellaneous | 2-Methylpyrazine | 1.4 ± 1.5 |
| *N*-containing compounds | Miscellaneous *N*-containing compounds | Miscellaneous | Trimethylpyrazine | 1.2 ± 0.9 |
| *N*-containing compounds | Miscellaneous *N*-containing compounds | Miscellaneous | 2-Isobutyl-3-methoxypyrazine | 0 ± 0 |
| *N*-containing compounds | Miscellaneous *N*-containing compounds | Miscellaneous | *N*-caproylhistamine | 0 ± 0 |
| *N*-containing compounds | Miscellaneous *N*-containing compounds | Miscellaneous | *N*-caproylhistidinol | 0 ± 0 |
| *N*-containing compounds | Miscellaneous *N*-containing compounds | Miscellaneous | *N*-caprylhistidinol | 0 ± 0 |
| *N*-containing compounds | Miscellaneous *N*-containing compounds | Miscellaneous | 2,5-dimethylpyrazine | 1.6 ± 1.7 |
| *N*-containing compounds | Miscellaneous *N*-containing compounds | Miscellaneous | 2,6-dimethylpyrazine | 0.5 ± 0.5 |

| Family | Class | Subclass | Compound | Intake (mg/day) Mean ± SD |
|---|---|---|---|---|
| *N*-containing compounds | Miscellaneous *N*-containing compounds | Miscellaneous | (-)-Salsolinol | 0.5 ± 0.1 |
| *N*-containing compounds | Miscellaneous *N*-containing compounds | Miscellaneous | DIBOA | 0.6 ± 0.6 |
| *N*-containing compounds | Miscellaneous *N*-containing compounds | Miscellaneous | DIMBOA | 0 ± 0 |
| *N*-containing compounds | Miscellaneous *N*-containing compounds | Miscellaneous | BOA | 0.5 ± 0.6 |
| *N*-containing compounds | Miscellaneous *N*-containing compounds | Miscellaneous | HBOA | 0.1 ± 0.1 |
| *N*-containing compounds | Miscellaneous *N*-containing compounds | Miscellaneous | HMBOA | 0 ± 0 |
| *N*-containing compounds | Miscellaneous *N*-containing compounds | Miscellaneous | MBOA | 0 ± 0 |
| *N*-containing compounds | Miscellaneous *N*-containing compounds | Miscellaneous | Synephrine | 59.7 ± 22.4 |

**Table S4: average intake of miscellaneous phytochemical family, classes, subclasses and individual compounds in the European adult population.**

| Family | Class | Subclass | Compound | Intake (mg/day) Mean ± SD |
|---|---|---|---|---|
| Miscellaneous phytochemicals | Total | - | - | 724.9 ± 71.6 |
| Miscellaneous phytochemicals | Phytates | Total | - | 636.6 ± 71.6 |
| Miscellaneous phytochemicals | Phytates | Phyates | Phytic acid | 636.6 ± 71.6 |
| Miscellaneous phytochemicals | Phytoprostanes | Phytoprostanes | Total | 0.1 ± 0.1 |
| Miscellaneous phytochemicals | Phytoprostanes | Phytoprostanes | 16-B1 PhytoP | 0 ± 0 |
| Miscellaneous phytochemicals | Phytoprostanes | Phytoprostanes | 9-D1t PhytoP | 0 ± 0 |
| Miscellaneous phytochemicals | Phytoprostanes | Phytoprostanes | F1 PhytoP | 0.1 ± 0 |
| Miscellaneous phytochemicals | Phytoprostanes | Phytoprostanes | 9-L1 PhytoP | 0 ± 0 |
| Miscellaneous phytochemicals | Thiosulfinates | Thiosulfinates | Total | 85.8 ± 34.1 |
| Miscellaneous phytochemicals | Thiosulfinates | Thiosulfinates | Ajoene | 0.2 ± 0.1 |
| Miscellaneous phytochemicals | Thiosulfinates | Thiosulfinates | Allicin | 10.9 ± 6.3 |
| Miscellaneous phytochemicals | Thiosulfinates | Thiosulfinates | Alliin | 5.6 ± 3.2 |
| Miscellaneous phytochemicals | Thiosulfinates | Thiosulfinates | Allin | 9.7 ± 5.6 |

| Family | Class | Subclass | Compound | Intake (mg/day) Mean ± SD |
|---|---|---|---|---|
| Miscellaneous phytochemicals | Thiosulfinates | Thiosulfinates | Allyl-2-Propenethiosulfinate | 5 ± 2.9 |
| Miscellaneous phytochemicals | Thiosulfinates | Thiosulfinates | Allyl-Methyl-Thiosulfinates | 1.7 ± 1 |
| Miscellaneous phytochemicals | Thiosulfinates | Thiosulfinates | Allyl-Propyl-Disulfide | 0.1 ± 0 |
| Miscellaneous phytochemicals | Thiosulfinates | Thiosulfinates | Allyl-Methyl-Disulfide | 0 ± 0 |
| Miscellaneous phytochemicals | Thiosulfinates | Thiosulfinates | Allyl-Methyl-Sulfide | 0 ± 0 |
| Miscellaneous phytochemicals | Thiosulfinates | Thiosulfinates | Allyl-Methyl-Tetrasulfide | 0 ± 0 |
| Miscellaneous phytochemicals | Thiosulfinates | Thiosulfinates | Allyl-Methyl-Trisulfide | 0.3 ± 0.2 |
| Miscellaneous phytochemicals | Thiosulfinates | Thiosulfinates | Allyl-Propyl-Trisulfide | 0.1 ± 0.1 |
| Miscellaneous phytochemicals | Thiosulfinates | Thiosulfinates | Cycloalliin | 1.8 ± 1 |
| Miscellaneous phytochemicals | Thiosulfinates | Thiosulfinates | Diallyl-Disulfide | 0.2 ± 0.1 |
| Miscellaneous phytochemicals | Thiosulfinates | Thiosulfinates | Diallyl-Sulfide | 0 ± 0 |
| Miscellaneous phytochemicals | Thiosulfinates | Thiosulfinates | Diallyl-Tetrasulfide | 0.1 ± 0.1 |
| Miscellaneous phytochemicals | Thiosulfinates | Thiosulfinates | Diallyl-Trisulfide | 0.7 ± 0.4 |
| Miscellaneous phytochemicals | Thiosulfinates | Thiosulfinates | Dimethyl-Disulfide | 0 ± 0 |
| Miscellaneous phytochemicals | Thiosulfinates | Thiosulfinates | Dimethyl-Trisulfide | 0 ± 0 |
| Miscellaneous phytochemicals | Thiosulfinates | Thiosulfinates | Dipropylsulfide | 0.1 ± 0.1 |
| Miscellaneous phytochemicals | Thiosulfinates | Thiosulfinates | Dipropyltetrasulfide | 0 ± 0 |
| Miscellaneous phytochemicals | Thiosulfinates | Thiosulfinates | γ-Glutamyl-*S*-Allylcysteine | 7.2 ± 4.1 |
| Miscellaneous phytochemicals | Thiosulfinates | Thiosulfinates | γ-Glutamyl-*S*-Allyl-Mercaptocysteine | 0 ± 0 |
| Miscellaneous phytochemicals | Thiosulfinates | Thiosulfinates | Isoalliin | 1.4 ± 0.8 |
| Miscellaneous phytochemicals | Thiosulfinates | Thiosulfinates | Methyl methanethiosulfinate | 0.1 ± 0.1 |
| Miscellaneous phytochemicals | Thiosulfinates | Thiosulfinates | *S*-Allyl-Cysteine | 0 ± 0 |
| Miscellaneous phytochemicals | Thiosulfinates | Thiosulfinates | *S*-Allylmercaptocysteine | 0 ± 0 |
| Miscellaneous phytochemicals | Thiosulfinates | Thiosulfinates | *S*-(2-propenyl)-L-cysteine sulphoxide | 6.7 ± 3.9 |
| Miscellaneous phytochemicals | Thiosulfinates | Thiosulfinates | *S*-Methyl-L-cysteine sulfoxide | 23.2 ± 5.9 |

| Family | Class | Subclass | Compound | Intake (mg/day) Mean ± SD |
|---|---|---|---|---|
| Miscellaneous phytochemicals | Thiosulfinates | Thiosulfinates | *S*-Propyl-L-cysteine Sulphoxide | 10.3 ± 2.8 |
| Miscellaneous phytochemicals | Miscellaneous | Miscellaneous | Total | 2.4 ± 1.2 |
| Miscellaneous phytochemicals | Miscellaneous | Miscellaneous | Estragole | 0.1 ± 0.1 |
| Miscellaneous phytochemicals | Miscellaneous | Miscellaneous | Falcarindiol | 0.7 ± 0.8 |
| Miscellaneous phytochemicals | Miscellaneous | Miscellaneous | Falcarinol | 0.7 ± 0.4 |
| Miscellaneous phytochemicals | Miscellaneous | Miscellaneous | Myristicin | 0.1 ± 0 |
| Miscellaneous phytochemicals | Miscellaneous | Miscellaneous | Oleocanthal | 1 ± 0.6 |
| Miscellaneous phytochemicals | Miscellaneous | Miscellaneous | Panaxydol | 0 ± 0 |
| Miscellaneous phytochemicals | Miscellaneous | Miscellaneous | Safrole | 0 ± 0 |

**Table S5: phytochemical intake distributions of (poly)phenols, terpenoids, *N*-containing compounds and miscellaneous phytochemicals in 26 European countries.**

| Country | Family | Class | Average intake (mg/day) | 5th percentile intake (mg/day) | 95th percentile intake (mg/day) |
|---|---|---|---|---|---|
| Austria | (Poly)phenols | Flavonoids | 187.7 | 0.0 | 913.9 |
| Belgium | (Poly)phenols | Flavonoids | 183.9 | 0.0 | 949.4 |
| Bosnia and Herzegovina | (Poly)phenols | Flavonoids | 114.3 | 0.0 | 535.2 |
| Croatia | (Poly)phenols | Flavonoids | 116.5 | 0.0 | 492.0 |
| Cyprus | (Poly)phenols | Flavonoids | 153.6 | 0.0 | 873.9 |
| Czechia | (Poly)phenols | Flavonoids | 211.2 | 0.0 | 756.3 |
| Denmark | (Poly)phenols | Flavonoids | 232.9 | 0.4 | 1052.3 |
| Estonia | (Poly)phenols | Flavonoids | 210.6 | 0.0 | 901.1 |
| Finland | (Poly)phenols | Flavonoids | 237.8 | 0.0 | 1332.9 |
| France | (Poly)phenols | Flavonoids | 227.7 | 0.0 | 1258.6 |
| Germany | (Poly)phenols | Flavonoids | 248.3 | 0.0 | 1011.7 |
| Greece | (Poly)phenols | Flavonoids | 108.3 | 0.0 | 482.8 |
| Hungary | (Poly)phenols | Flavonoids | 216.6 | 0.0 | 873.8 |
| Ireland | (Poly)phenols | Flavonoids | 331.3 | 0.0 | 1067.7 |
| Italy | (Poly)phenols | Flavonoids | 146.4 | 0.0 | 838.2 |
| Latvia | (Poly)phenols | Flavonoids | 240.9 | 0.0 | 1006.3 |
| Montenegro | (Poly)phenols | Flavonoids | 118.1 | 0.0 | 634.1 |
| Netherlands | (Poly)phenols | Flavonoids | 336.1 | 0.0 | 1778.3 |

| Country | Family | Class | Average intake (mg/day) | $5^{th}$ percentile intake (mg/day) | $95^{th}$ percentile intake (mg/day) |
|---|---|---|---|---|---|
| Poland | (Poly)phenols | Flavonoids | 221.4 | 0.0 | 679.8 |
| Portugal | (Poly)phenols | Flavonoids | 134.6 | 0.0 | 567.7 |
| Romania | (Poly)phenols | Flavonoids | 143.1 | 0.1 | 710.4 |
| Serbia | (Poly)phenols | Flavonoids | 152.4 | 0.0 | 708.0 |
| Slovenia | (Poly)phenols | Flavonoids | 113.6 | 0.0 | 399.9 |
| Spain | (Poly)phenols | Flavonoids | 143.6 | 0.0 | 728.0 |
| Sweden | (Poly)phenols | Flavonoids | 180.1 | 0.0 | 810.8 |
| United Kingdom | (Poly)phenols | Flavonoids | 326.6 | 0.0 | 1199.2 |
| Austria | (Poly)phenols | Phenolic acids | 724.5 | 0.0 | 2273.2 |
| Belgium | (Poly)phenols | Phenolic acids | 686.3 | 0.0 | 2374.7 |
| Bosnia and Herzegovina | (Poly)phenols | Phenolic acids | 658.1 | 0.0 | 3022.0 |
| Croatia | (Poly)phenols | Phenolic acids | 542.2 | 0.0 | 2309.7 |
| Cyprus | (Poly)phenols | Phenolic acids | 967.9 | 0.0 | 4516.3 |
| Czechia | (Poly)phenols | Phenolic acids | 1105.8 | 0.0 | 3517.6 |
| Denmark | (Poly)phenols | Phenolic acids | 1551.0 | 4.1 | 4205.3 |
| Estonia | (Poly)phenols | Phenolic acids | 918.9 | 0.0 | 2575.1 |
| Finland | (Poly)phenols | Phenolic acids | 1150.4 | 0.0 | 3450.3 |
| France | (Poly)phenols | Phenolic acids | 837.1 | 0.0 | 3400.2 |
| Germany | (Poly)phenols | Phenolic acids | 1252.3 | 0.0 | 3687.2 |
| Greece | (Poly)phenols | Phenolic acids | 1050.6 | 0.0 | 3896.9 |
| Hungary | (Poly)phenols | Phenolic acids | 1023.8 | 0.0 | 5522.0 |
| Ireland | (Poly)phenols | Phenolic acids | 611.8 | 0.0 | 2531.9 |
| Italy | (Poly)phenols | Phenolic acids | 519.9 | 0.0 | 1770.0 |
| Latvia | (Poly)phenols | Phenolic acids | 872.3 | 0.0 | 3007.7 |
| Montenegro | (Poly)phenols | Phenolic acids | 690.7 | 0.0 | 2856.5 |
| Netherlands | (Poly)phenols | Phenolic acids | 1070.1 | 0.0 | 3730.0 |
| Poland | (Poly)phenols | Phenolic acids | 473.5 | 0.0 | 1689.0 |
| Portugal | (Poly)phenols | Phenolic acids | 325.1 | 0.0 | 1104.1 |
| Romania | (Poly)phenols | Phenolic acids | 543.9 | 0.0 | 1958.5 |
| Serbia | (Poly)phenols | Phenolic acids | 863.4 | 0.0 | 3268.8 |
| Slovenia | (Poly)phenols | Phenolic acids | 380.8 | 0.0 | 1160.7 |
| Spain | (Poly)phenols | Phenolic acids | 361.6 | 0.0 | 2201.3 |
| Sweden | (Poly)phenols | Phenolic acids | 337.4 | 0.0 | 1617.6 |
| United Kingdom | (Poly)phenols | Phenolic acids | 562.7 | 0.0 | 2266.2 |
| Austria | (Poly)phenols | Tannins | 293.4 | 0.0 | 1156.3 |
| Belgium | (Poly)phenols | Tannins | 297.9 | 0.0 | 1435.4 |
| Bosnia and Herzegovina | (Poly)phenols | Tannins | 190.3 | 0.0 | 867.7 |
| Croatia | (Poly)phenols | Tannins | 222.9 | 0.0 | 722.4 |

| Country | Family | Class | Average intake (mg/day) | 5th percentile intake (mg/day) | 95th percentile intake (mg/day) |
|---|---|---|---|---|---|
| Cyprus | (Poly)phenols | Tannins | 155.2 | 0.0 | 654.5 |
| Czechia | (Poly)phenols | Tannins | 180.0 | 0.0 | 602.0 |
| Denmark | (Poly)phenols | Tannins | 306.7 | 0.0 | 1193.5 |
| Estonia | (Poly)phenols | Tannins | 281.0 | 0.0 | 1067.6 |
| Finland | (Poly)phenols | Tannins | 175.5 | 0.0 | 948.4 |
| France | (Poly)phenols | Tannins | 284.2 | 0.0 | 1427.1 |
| Germany | (Poly)phenols | Tannins | 316.8 | 0.0 | 1009.3 |
| Greece | (Poly)phenols | Tannins | 165.4 | 0.0 | 601.3 |
| Hungary | (Poly)phenols | Tannins | 187.2 | 0.0 | 628.1 |
| Ireland | (Poly)phenols | Tannins | 207.4 | 0.0 | 811.2 |
| Italy | (Poly)phenols | Tannins | 204.9 | 0.0 | 1067.8 |
| Latvia | (Poly)phenols | Tannins | 231.6 | 0.0 | 949.4 |
| Montenegro | (Poly)phenols | Tannins | 211.9 | 0.0 | 1106.8 |
| Netherlands | (Poly)phenols | Tannins | 274.4 | 0.0 | 1200.5 |
| Poland | (Poly)phenols | Tannins | 156.7 | 0.0 | 696.6 |
| Portugal | (Poly)phenols | Tannins | 206.0 | 0.0 | 818.3 |
| Romania | (Poly)phenols | Tannins | 237.2 | 0.0 | 1170.3 |
| Serbia | (Poly)phenols | Tannins | 252.4 | 0.0 | 1115.7 |
| Slovenia | (Poly)phenols | Tannins | 227.6 | 0.0 | 574.5 |
| Spain | (Poly)phenols | Tannins | 218.5 | 0.0 | 1068.1 |
| Sweden | (Poly)phenols | Tannins | 227.8 | 0.0 | 1284.4 |
| United Kingdom | (Poly)phenols | Tannins | 287.5 | 0.0 | 1336.4 |
| Austria | (Poly)phenols | Miscellaneous (poly)phenols | 52.0 | 0.0 | 225.7 |
| Belgium | (Poly)phenols | Miscellaneous (poly)phenols | 47.9 | 0.0 | 177.6 |
| Bosnia and Herzegovina | (Poly)phenols | Miscellaneous (poly)phenols | 51.8 | 0.0 | 192.5 |
| Croatia | (Poly)phenols | Miscellaneous (poly)phenols | 33.3 | 0.0 | 143.9 |
| Cyprus | (Poly)phenols | Miscellaneous (poly)phenols | 61.3 | 0.0 | 327.5 |
| Czechia | (Poly)phenols | Miscellaneous (poly)phenols | 46.6 | 0.0 | 171.6 |
| Denmark | (Poly)phenols | Miscellaneous (poly)phenols | 91.3 | 3.4 | 255.6 |
| Estonia | (Poly)phenols | Miscellaneous (poly)phenols | 44.2 | 0.0 | 159.9 |
| Finland | (Poly)phenols | Miscellaneous (poly)phenols | 51.1 | 0.0 | 207.3 |
| France | (Poly)phenols | Miscellaneous (poly)phenols | 53.2 | 0.0 | 209.2 |
| Germany | (Poly)phenols | Miscellaneous (poly)phenols | 61.4 | 0.0 | 288.5 |

| Country | Family | Class | Average intake (mg/day) | 5th percentile intake (mg/day) | 95th percentile intake (mg/day) |
|---|---|---|---|---|---|
| Greece | (Poly)phenols | Miscellaneous (poly)phenols | 59.6 | 1.3 | 248.0 |
| Hungary | (Poly)phenols | Miscellaneous (poly)phenols | 59.9 | 0.0 | 258.9 |
| Ireland | (Poly)phenols | Miscellaneous (poly)phenols | 71.4 | 0.0 | 340.8 |
| Italy | (Poly)phenols | Miscellaneous (poly)phenols | 52.6 | 3.0 | 221.6 |
| Latvia | (Poly)phenols | Miscellaneous (poly)phenols | 55.2 | 0.0 | 213.7 |
| Montenegro | (Poly)phenols | Miscellaneous (poly)phenols | 53.4 | 0.0 | 196.4 |
| Netherlands | (Poly)phenols | Miscellaneous (poly)phenols | 55.9 | 0.0 | 208.5 |
| Poland | (Poly)phenols | Miscellaneous (poly)phenols | 36.1 | 0.0 | 172.2 |
| Portugal | (Poly)phenols | Miscellaneous (poly)phenols | 48.0 | 0.3 | 196.0 |
| Romania | (Poly)phenols | Miscellaneous (poly)phenols | 57.9 | 0.0 | 226.4 |
| Serbia | (Poly)phenols | Miscellaneous (poly)phenols | 66.0 | 0.0 | 223.5 |
| Slovenia | (Poly)phenols | Miscellaneous (poly)phenols | 41.7 | 0.0 | 176.4 |
| Spain | (Poly)phenols | Miscellaneous (poly)phenols | 42.6 | 0.0 | 187.2 |
| Sweden | (Poly)phenols | Miscellaneous (poly)phenols | 37.3 | 0.0 | 175.4 |
| United Kingdom | (Poly)phenols | Miscellaneous (poly)phenols | 56.6 | 0.0 | 263.3 |
| Austria | Terpenoids | Tetraterpenoids | 72.0 | 0.0 | 336.5 |
| Belgium | Terpenoids | Tetraterpenoids | 40.5 | 0.0 | 219.9 |
| Bosnia and Herzegovina | Terpenoids | Tetraterpenoids | 52.8 | 0.0 | 330.2 |
| Croatia | Terpenoids | Tetraterpenoids | 45.4 | 0.0 | 268.5 |
| Cyprus | Terpenoids | Tetraterpenoids | 25.6 | 0.0 | 132.5 |
| Czechia | Terpenoids | Tetraterpenoids | 40.6 | 0.0 | 194.1 |
| Denmark | Terpenoids | Tetraterpenoids | 47.8 | 0.6 | 149.2 |
| Estonia | Terpenoids | Tetraterpenoids | 31.6 | 0.0 | 145.1 |
| Finland | Terpenoids | Tetraterpenoids | 43.3 | 0.0 | 196.1 |
| France | Terpenoids | Tetraterpenoids | 35.4 | 0.0 | 180.7 |
| Germany | Terpenoids | Tetraterpenoids | 34.0 | 0.0 | 222.3 |
| Greece | Terpenoids | Tetraterpenoids | 45.2 | 0.3 | 215.4 |
| Hungary | Terpenoids | Tetraterpenoids | 104.1 | 0.0 | 466.6 |
| Ireland | Terpenoids | Tetraterpenoids | 42.0 | 0.0 | 185.2 |
| Italy | Terpenoids | Tetraterpenoids | 42.4 | 0.2 | 267.5 |
| Latvia | Terpenoids | Tetraterpenoids | 41.4 | 0.0 | 191.6 |

| Country | Family | Class | Average intake (mg/day) | 5$^{th}$ percentile intake (mg/day) | 95$^{th}$ percentile intake (mg/day) |
|---|---|---|---|---|---|
| Montenegro | Terpenoids | Tetraterpenoids | 65.7 | 0.0 | 373.3 |
| Netherlands | Terpenoids | Tetraterpenoids | 37.0 | 0.0 | 189.7 |
| Poland | Terpenoids | Tetraterpenoids | 26.2 | 0.0 | 138.8 |
| Portugal | Terpenoids | Tetraterpenoids | 27.7 | 0.1 | 129.5 |
| Romania | Terpenoids | Tetraterpenoids | 54.5 | 0.0 | 262.2 |
| Serbia | Terpenoids | Tetraterpenoids | 82.3 | 0.0 | 387.0 |
| Slovenia | Terpenoids | Tetraterpenoids | 47.1 | 0.0 | 216.9 |
| Spain | Terpenoids | Tetraterpenoids | 36.4 | 0.0 | 160.8 |
| Sweden | Terpenoids | Tetraterpenoids | 45.0 | 0.0 | 203.6 |
| United Kingdom | Terpenoids | Tetraterpenoids | 36.0 | 0.0 | 188.3 |
| Austria | Terpenoids | Triterpenoids | 641.0 | 0.0 | 2867.7 |
| Belgium | Terpenoids | Triterpenoids | 541.3 | 0.0 | 2288.1 |
| Bosnia and Herzegovina | Terpenoids | Triterpenoids | 465.9 | 0.0 | 1960.8 |
| Croatia | Terpenoids | Triterpenoids | 484.0 | 4.5 | 1911.9 |
| Cyprus | Terpenoids | Triterpenoids | 561.1 | 0.0 | 2245.9 |
| Czechia | Terpenoids | Triterpenoids | 455.3 | 0.0 | 1717.3 |
| Denmark | Terpenoids | Triterpenoids | 620.1 | 14.2 | 2212.5 |
| Estonia | Terpenoids | Triterpenoids | 546.9 | 0.0 | 1967.6 |
| Finland | Terpenoids | Triterpenoids | 601.8 | 0.0 | 2524.4 |
| France | Terpenoids | Triterpenoids | 683.4 | 0.0 | 2874.8 |
| Germany | Terpenoids | Triterpenoids | 558.5 | 0.0 | 2495.6 |
| Greece | Terpenoids | Triterpenoids | 613.0 | 35.1 | 2289.5 |
| Hungary | Terpenoids | Triterpenoids | 576.0 | 9.0 | 1977.9 |
| Ireland | Terpenoids | Triterpenoids | 542.7 | 0.0 | 2082.5 |
| Italy | Terpenoids | Triterpenoids | 673.3 | 38.2 | 2713.0 |
| Latvia | Terpenoids | Triterpenoids | 661.3 | 0.0 | 2380.4 |
| Montenegro | Terpenoids | Triterpenoids | 483.2 | 0.0 | 2050.7 |
| Netherlands | Terpenoids | Triterpenoids | 582.1 | 0.0 | 2651.8 |
| Poland | Terpenoids | Triterpenoids | 519.8 | 0.0 | 2149.1 |
| Portugal | Terpenoids | Triterpenoids | 626.9 | 8.8 | 2232.2 |
| Romania | Terpenoids | Triterpenoids | 462.0 | 0.1 | 1722.8 |
| Serbia | Terpenoids | Triterpenoids | 547.6 | 0.0 | 2330.7 |
| Slovenia | Terpenoids | Triterpenoids | 460.1 | 0.0 | 1817.5 |
| Spain | Terpenoids | Triterpenoids | 430.1 | 0.0 | 1640.8 |
| Sweden | Terpenoids | Triterpenoids | 569.8 | 0.0 | 2706.4 |
| United Kingdom | Terpenoids | Triterpenoids | 471.4 | 0.0 | 1960.2 |
| Austria | Terpenoids | Diterpenoids | 13.6 | 0.0 | 59.7 |
| Belgium | Terpenoids | Diterpenoids | 3.9 | 0.0 | 19.4 |

| Country | Family | Class | Average intake (mg/day) | 5th percentile intake (mg/day) | 95th percentile intake (mg/day) |
|---|---|---|---|---|---|
| Bosnia and Herzegovina | Terpenoids | Diterpenoids | 11.4 | 0.0 | 60.6 |
| Croatia | Terpenoids | Diterpenoids | 8.2 | 0.0 | 44.8 |
| Cyprus | Terpenoids | Diterpenoids | 23.6 | 0.0 | 116.2 |
| Czechia | Terpenoids | Diterpenoids | 61.7 | 0.0 | 204.6 |
| Denmark | Terpenoids | Diterpenoids | 5.5 | 0.0 | 16.6 |
| Estonia | Terpenoids | Diterpenoids | 8.6 | 0.0 | 29.0 |
| Finland | Terpenoids | Diterpenoids | 6.1 | 0.0 | 29.4 |
| France | Terpenoids | Diterpenoids | 5.8 | 0.0 | 23.3 |
| Germany | Terpenoids | Diterpenoids | 11.0 | 0.0 | 43.5 |
| Greece | Terpenoids | Diterpenoids | 55.5 | 0.0 | 192.7 |
| Hungary | Terpenoids | Diterpenoids | 40.3 | 0.0 | 228.3 |
| Ireland | Terpenoids | Diterpenoids | 17.5 | 0.0 | 73.0 |
| Italy | Terpenoids | Diterpenoids | 7.0 | 0.0 | 32.7 |
| Latvia | Terpenoids | Diterpenoids | 11.8 | 0.0 | 44.4 |
| Montenegro | Terpenoids | Diterpenoids | 13.7 | 0.0 | 68.5 |
| Netherlands | Terpenoids | Diterpenoids | 6.9 | 0.0 | 24.1 |
| Poland | Terpenoids | Diterpenoids | 2.6 | 0.0 | 15.8 |
| Portugal | Terpenoids | Diterpenoids | 4.7 | 0.0 | 16.3 |
| Romania | Terpenoids | Diterpenoids | 11.8 | 0.0 | 51.6 |
| Serbia | Terpenoids | Diterpenoids | 17.3 | 0.0 | 72.1 |
| Slovenia | Terpenoids | Diterpenoids | 7.6 | 0.0 | 28.3 |
| Spain | Terpenoids | Diterpenoids | 6.5 | 0.0 | 50.8 |
| Sweden | Terpenoids | Diterpenoids | 6.3 | 0.0 | 30.9 |
| United Kingdom | Terpenoids | Diterpenoids | 16.9 | 0.0 | 72.6 |
| Austria | Terpenoids | Sesquiterpenoids | 166.5 | 0.0 | 744.8 |
| Belgium | Terpenoids | Sesquiterpenoids | 118.0 | 0.0 | 474.4 |
| Bosnia and Herzegovina | Terpenoids | Sesquiterpenoids | 51.4 | 0.0 | 229.5 |
| Croatia | Terpenoids | Sesquiterpenoids | 71.5 | 0.0 | 284.3 |
| Cyprus | Terpenoids | Sesquiterpenoids | 65.9 | 0.0 | 354.0 |
| Czechia | Terpenoids | Sesquiterpenoids | 42.8 | 0.0 | 168.6 |
| Denmark | Terpenoids | Sesquiterpenoids | 219.9 | 0.0 | 782.6 |
| Estonia | Terpenoids | Sesquiterpenoids | 129.7 | 0.0 | 404.3 |
| Finland | Terpenoids | Sesquiterpenoids | 132.3 | 0.0 | 476.6 |
| France | Terpenoids | Sesquiterpenoids | 135.1 | 0.0 | 540.3 |
| Germany | Terpenoids | Sesquiterpenoids | 138.1 | 0.0 | 821.3 |
| Greece | Terpenoids | Sesquiterpenoids | 41.9 | 0.0 | 226.7 |
| Hungary | Terpenoids | Sesquiterpenoids | 99.9 | 0.0 | 363.6 |
| Ireland | Terpenoids | Sesquiterpenoids | 150.2 | 0.0 | 481.9 |
| Italy | Terpenoids | Sesquiterpenoids | 76.4 | 0.0 | 382.7 |

| Country | Family | Class | Average intake (mg/day) | 5th percentile intake (mg/day) | 95th percentile intake (mg/day) |
|---|---|---|---|---|---|
| Latvia | Terpenoids | Sesquiterpenoids | 209.9 | 0.0 | 611.4 |
| Montenegro | Terpenoids | Sesquiterpenoids | 55.0 | 0.0 | 157.7 |
| Netherlands | Terpenoids | Sesquiterpenoids | 90.9 | 0.0 | 504.5 |
| Poland | Terpenoids | Sesquiterpenoids | 100.6 | 0.0 | 327.2 |
| Portugal | Terpenoids | Sesquiterpenoids | 141.5 | 0.0 | 419.7 |
| Romania | Terpenoids | Sesquiterpenoids | 64.3 | 0.0 | 223.5 |
| Serbia | Terpenoids | Sesquiterpenoids | 67.8 | 0.0 | 232.2 |
| Slovenia | Terpenoids | Sesquiterpenoids | 63.4 | 0.0 | 213.4 |
| Spain | Terpenoids | Sesquiterpenoids | 51.7 | 0.0 | 156.4 |
| Sweden | Terpenoids | Sesquiterpenoids | 67.1 | 0.0 | 258.5 |
| United Kingdom | Terpenoids | Sesquiterpenoids | 125.8 | 0.0 | 497.4 |
| Austria | Terpenoids | Monoterpenoids | 1225.7 | 0.0 | 5694.4 |
| Belgium | Terpenoids | Monoterpenoids | 1971.0 | 0.0 | 11175.3 |
| Bosnia and Herzegovina | Terpenoids | Monoterpenoids | 906.0 | 0.0 | 4243.5 |
| Croatia | Terpenoids | Monoterpenoids | 850.3 | 0.0 | 4577.5 |
| Cyprus | Terpenoids | Monoterpenoids | 1212.1 | 0.0 | 7090.9 |
| Czechia | Terpenoids | Monoterpenoids | 524.7 | 0.0 | 3195.1 |
| Denmark | Terpenoids | Monoterpenoids | 1726.4 | 0.0 | 7160.8 |
| Estonia | Terpenoids | Monoterpenoids | 841.9 | 0.0 | 5618.2 |
| Finland | Terpenoids | Monoterpenoids | 1546.5 | 0.0 | 9130.0 |
| France | Terpenoids | Monoterpenoids | 1911.8 | 0.0 | 9194.5 |
| Germany | Terpenoids | Monoterpenoids | 2967.4 | 0.0 | 14333.3 |
| Greece | Terpenoids | Monoterpenoids | 1002.4 | 0.0 | 4295.5 |
| Hungary | Terpenoids | Monoterpenoids | 1316.0 | 0.0 | 5356.6 |
| Ireland | Terpenoids | Monoterpenoids | 1545.4 | 0.0 | 8727.5 |
| Italy | Terpenoids | Monoterpenoids | 1252.3 | 0.0 | 7329.0 |
| Latvia | Terpenoids | Monoterpenoids | 964.9 | 0.0 | 6026.5 |
| Montenegro | Terpenoids | Monoterpenoids | 1005.4 | 0.0 | 6069.6 |
| Netherlands | Terpenoids | Monoterpenoids | 2165.4 | 0.0 | 12908.5 |
| Poland | Terpenoids | Monoterpenoids | 1078.2 | 0.0 | 4026.3 |
| Portugal | Terpenoids | Monoterpenoids | 1592.4 | 0.0 | 9774.3 |
| Romania | Terpenoids | Monoterpenoids | 833.8 | 0.0 | 1796.5 |
| Serbia | Terpenoids | Monoterpenoids | 963.8 | 0.0 | 3540.2 |
| Slovenia | Terpenoids | Monoterpenoids | 1203.3 | 0.0 | 5330.0 |
| Spain | Terpenoids | Monoterpenoids | 1694.1 | 0.0 | 8409.7 |
| Sweden | Terpenoids | Monoterpenoids | 1289.0 | 0.0 | 7473.3 |
| United Kingdom | Terpenoids | Monoterpenoids | 1356.6 | 0.0 | 7188.4 |
| Austria | *N*-containing compounds | Alkaloids | 756.2 | 0.0 | 2616.6 |

| Country | Family | Class | Average intake (mg/day) | 5th percentile intake (mg/day) | 95th percentile intake (mg/day) |
|---|---|---|---|---|---|
| Belgium | *N*-containing compounds | Alkaloids | 373.3 | 0.0 | 1481.5 |
| Bosnia and Herzegovina | *N*-containing compounds | Alkaloids | 927.6 | 0.0 | 4013.9 |
| Croatia | *N*-containing compounds | Alkaloids | 566.1 | 0.0 | 2394.9 |
| Cyprus | *N*-containing compounds | Alkaloids | 532.3 | 0.0 | 2748.7 |
| Czechia | *N*-containing compounds | Alkaloids | 486.6 | 0.0 | 1582.4 |
| Denmark | *N*-containing compounds | Alkaloids | 771.3 | 0.4 | 2207.1 |
| Estonia | *N*-containing compounds | Alkaloids | 842.9 | 0.0 | 2527.6 |
| Finland | *N*-containing compounds | Alkaloids | 594.3 | 0.0 | 2407.8 |
| France | *N*-containing compounds | Alkaloids | 641.8 | 0.0 | 2852.2 |
| Germany | *N*-containing compounds | Alkaloids | 1134.7 | 0.0 | 3289.3 |
| Greece | *N*-containing compounds | Alkaloids | 405.2 | 0.0 | 1236.8 |
| Hungary | *N*-containing compounds | Alkaloids | 551.0 | 0.0 | 2191.0 |
| Ireland | *N*-containing compounds | Alkaloids | 262.1 | 0.0 | 1089.3 |
| Italy | *N*-containing compounds | Alkaloids | 451.1 | 0.0 | 1841.0 |
| Latvia | *N*-containing compounds | Alkaloids | 501.5 | 0.0 | 2084.1 |
| Montenegro | *N*-containing compounds | Alkaloids | 976.2 | 0.0 | 4000.3 |
| Netherlands | *N*-containing compounds | Alkaloids | 608.5 | 0.0 | 2606.0 |
| Poland | *N*-containing compounds | Alkaloids | 265.5 | 0.0 | 1050.1 |
| Portugal | *N*-containing compounds | Alkaloids | 398.3 | 0.0 | 1417.6 |
| Romania | *N*-containing compounds | Alkaloids | 744.1 | 0.0 | 2538.9 |
| Serbia | *N*-containing compounds | Alkaloids | 1282.4 | 0.0 | 4695.7 |
| Slovenia | *N*-containing compounds | Alkaloids | 314.4 | 0.0 | 956.2 |
| Spain | *N*-containing compounds | Alkaloids | 133.4 | 0.0 | 599.0 |
| Sweden | *N*-containing compounds | Alkaloids | 355.6 | 0.0 | 1771.0 |
| United Kingdom | *N*-containing compounds | Alkaloids | 266.4 | 0.0 | 1017.1 |

| Country | Family | Class | Average intake (mg/day) | 5th percentile intake (mg/day) | 95th percentile intake (mg/day) |
|---|---|---|---|---|---|
| Austria | *N*-containing compounds | Glucosinolates & isothiocyanates | 37.4 | 0.0 | 164.9 |
| Belgium | *N*-containing compounds | Glucosinolates & isothiocyanates | 31.8 | 0.0 | 167.8 |
| Bosnia and Herzegovina | *N*-containing compounds | Glucosinolates & isothiocyanates | 15.9 | 0.0 | 109.0 |
| Croatia | *N*-containing compounds | Glucosinolates & isothiocyanates | 27.0 | 0.0 | 137.4 |
| Cyprus | *N*-containing compounds | Glucosinolates & isothiocyanates | 28.3 | 0.0 | 203.5 |
| Czechia | *N*-containing compounds | Glucosinolates & isothiocyanates | 26.1 | 0.0 | 106.7 |
| Denmark | *N*-containing compounds | Glucosinolates & isothiocyanates | 27.0 | 0.0 | 120.6 |
| Estonia | *N*-containing compounds | Glucosinolates & isothiocyanates | 40.6 | 0.0 | 204.4 |
| Finland | *N*-containing compounds | Glucosinolates & isothiocyanates | 29.2 | 0.0 | 161.5 |
| France | *N*-containing compounds | Glucosinolates & isothiocyanates | 22.1 | 0.0 | 80.6 |
| Germany | *N*-containing compounds | Glucosinolates & isothiocyanates | 26.5 | 0.0 | 68.6 |
| Greece | *N*-containing compounds | Glucosinolates & isothiocyanates | 26.3 | 0.0 | 60.4 |
| Hungary | *N*-containing compounds | Glucosinolates & isothiocyanates | 35.2 | 0.0 | 169.2 |
| Ireland | *N*-containing compounds | Glucosinolates & isothiocyanates | 37.8 | 0.0 | 205.5 |
| Italy | *N*-containing compounds | Glucosinolates & isothiocyanates | 26.5 | 0.0 | 35.6 |
| Latvia | *N*-containing compounds | Glucosinolates & isothiocyanates | 53.4 | 0.0 | 212.8 |
| Montenegro | *N*-containing compounds | Glucosinolates & isothiocyanates | 21.5 | 0.0 | 107.5 |
| Netherlands | *N*-containing compounds | Glucosinolates & isothiocyanates | 55.6 | 0.0 | 342.0 |
| Poland | *N*-containing compounds | Glucosinolates & isothiocyanates | 34.0 | 0.0 | 143.3 |
| Portugal | *N*-containing compounds | Glucosinolates & isothiocyanates | 42.7 | 0.0 | 258.9 |
| Romania | *N*-containing compounds | Glucosinolates & isothiocyanates | 18.8 | 0.0 | 99.7 |
| Serbia | *N*-containing compounds | Glucosinolates & isothiocyanates | 30.6 | 0.0 | 135.6 |
| Slovenia | *N*-containing compounds | Glucosinolates & isothiocyanates | 22.6 | 0.0 | 93.6 |
| Spain | *N*-containing compounds | Glucosinolates & isothiocyanates | 12.1 | 0.0 | 17.6 |
| Sweden | *N*-containing compounds | Glucosinolates & isothiocyanates | 29.8 | 0.0 | 123.2 |

| **Country** | **Family** | **Class** | **Average intake (mg/day)** | **5th percentile intake (mg/day)** | **95th percentile intake (mg/day)** |
|---|---|---|---|---|---|
| United Kingdom | *N*-containing compounds | Glucosinolates & isothiocyanates | 41.9 | 0.0 | 239.9 |
| Austria | *N*-containing compounds | Miscellaneous *N*-containing compounds | 568.7 | 0.0 | 3657.9 |
| Belgium | *N*-containing compounds | Miscellaneous *N*-containing compounds | 540.8 | 0.0 | 2792.4 |
| Bosnia and Herzegovina | *N*-containing compounds | Miscellaneous *N*-containing compounds | 508.4 | 0.0 | 2810.5 |
| Croatia | *N*-containing compounds | Miscellaneous *N*-containing compounds | 381.2 | 0.0 | 2403.9 |
| Cyprus | *N*-containing compounds | Miscellaneous *N*-containing compounds | 293.4 | 0.0 | 1663.4 |
| Czechia | *N*-containing compounds | Miscellaneous *N*-containing compounds | 356.8 | 0.0 | 1440.5 |
| Denmark | *N*-containing compounds | Miscellaneous *N*-containing compounds | 396.9 | 9.4 | 1420.2 |
| Estonia | *N*-containing compounds | Miscellaneous *N*-containing compounds | 468.2 | 0.0 | 2634.7 |
| Finland | *N*-containing compounds | Miscellaneous *N*-containing compounds | 648.3 | 0.0 | 3723.9 |
| France | *N*-containing compounds | Miscellaneous *N*-containing compounds | 831.3 | 0.0 | 4953.0 |
| Germany | *N*-containing compounds | Miscellaneous *N*-containing compounds | 734.0 | 0.0 | 3675.0 |
| Greece | *N*-containing compounds | Miscellaneous *N*-containing compounds | 320.0 | 0.0 | 1286.3 |
| Hungary | *N*-containing compounds | Miscellaneous *N*-containing compounds | 557.5 | 0.0 | 2814.9 |
| Ireland | *N*-containing compounds | Miscellaneous *N*-containing compounds | 398.8 | 0.0 | 1363.1 |
| Italy | *N*-containing compounds | Miscellaneous *N*-containing compounds | 425.0 | 0.0 | 2828.6 |
| Latvia | *N*-containing compounds | Miscellaneous *N*-containing compounds | 898.1 | 0.0 | 4357.8 |
| Montenegro | *N*-containing compounds | Miscellaneous *N*-containing compounds | 404.5 | 0.0 | 2596.9 |
| Netherlands | *N*-containing compounds | Miscellaneous *N*-containing compounds | 790.9 | 0.0 | 4912.4 |
| Poland | *N*-containing compounds | Miscellaneous *N*-containing compounds | 371.8 | 0.0 | 1332.2 |
| Portugal | *N*-containing compounds | Miscellaneous *N*-containing compounds | 400.3 | 0.0 | 1433.4 |
| Romania | *N*-containing compounds | Miscellaneous *N*-containing compounds | 337.8 | 0.0 | 1940.5 |
| Serbia | *N*-containing compounds | Miscellaneous *N*-containing compounds | 595.4 | 0.0 | 3178.4 |
| Slovenia | *N*-containing compounds | Miscellaneous *N*-containing compounds | 299.5 | 0.0 | 1278.6 |
| Spain | *N*-containing compounds | Miscellaneous *N*-containing compounds | 287.2 | 0.0 | 1405.4 |

| Country | Family | Class | Average intake (mg/day) | 5th percentile intake (mg/day) | 95th percentile intake (mg/day) |
|---|---|---|---|---|---|
| Sweden | *N*-containing compounds | Miscellaneous *N*-containing compounds | 264.0 | 0.0 | 1304.3 |
| United Kingdom | *N*-containing compounds | Miscellaneous *N*-containing compounds | 372.5 | 0.0 | 1403.2 |
| Austria | Miscellaneous phytochemicals | Phytates | 664.4 | 0.0 | 2487.2 |
| Belgium | Miscellaneous phytochemicals | Phytates | 556.3 | 0.0 | 1847.3 |
| Bosnia and Herzegovina | Miscellaneous phytochemicals | Phytates | 596.5 | 0.0 | 2595.6 |
| Croatia | Miscellaneous phytochemicals | Phytates | 590.0 | 0.0 | 2265.9 |
| Cyprus | Miscellaneous phytochemicals | Phytates | 702.5 | 0.0 | 2818.6 |
| Czechia | Miscellaneous phytochemicals | Phytates | 562.3 | 0.0 | 2188.2 |
| Denmark | Miscellaneous phytochemicals | Phytates | 657.9 | 11.0 | 2472.3 |
| Estonia | Miscellaneous phytochemicals | Phytates | 674.2 | 0.0 | 2672.3 |
| Finland | Miscellaneous phytochemicals | Phytates | 702.9 | 0.0 | 3085.6 |
| France | Miscellaneous phytochemicals | Phytates | 618.2 | 0.0 | 2449.2 |
| Germany | Miscellaneous phytochemicals | Phytates | 723.3 | 0.0 | 3208.0 |
| Greece | Miscellaneous phytochemicals | Phytates | 532.2 | 0.0 | 2199.4 |
| Hungary | Miscellaneous phytochemicals | Phytates | 694.3 | 0.0 | 2548.2 |
| Ireland | Miscellaneous phytochemicals | Phytates | 758.0 | 0.0 | 3107.0 |
| Italy | Miscellaneous phytochemicals | Phytates | 649.4 | 0.0 | 2762.9 |
| Latvia | Miscellaneous phytochemicals | Phytates | 637.4 | 0.0 | 2438.1 |
| Montenegro | Miscellaneous phytochemicals | Phytates | 644.5 | 0.0 | 2628.3 |
| Netherlands | Miscellaneous phytochemicals | Phytates | 672.5 | 0.0 | 2749.6 |
| Poland | Miscellaneous phytochemicals | Phytates | 534.9 | 0.0 | 2318.1 |
| Portugal | Miscellaneous phytochemicals | Phytates | 624.0 | 0.0 | 2204.8 |
| Romania | Miscellaneous phytochemicals | Phytates | 561.9 | 0.1 | 1962.3 |
| Serbia | Miscellaneous phytochemicals | Phytates | 745.3 | 0.0 | 3232.8 |
| Slovenia | Miscellaneous phytochemicals | Phytates | 674.6 | 0.0 | 2538.3 |

| Country | Family | Class | Average intake (mg/day) | 5th percentile intake (mg/day) | 95th percentile intake (mg/day) |
|---|---|---|---|---|---|
| Spain | Miscellaneous phytochemicals | Phytates | 467.0 | 0.0 | 2151.5 |
| Sweden | Miscellaneous phytochemicals | Phytates | 607.6 | 0.0 | 2653.8 |
| United Kingdom | Miscellaneous phytochemicals | Phytates | 699.3 | 0.0 | 2835.1 |
| Austria | Miscellaneous phytochemicals | Phytoprostanes | 0.1 | 0.0 | 0.3 |
| Belgium | Miscellaneous phytochemicals | Phytoprostanes | 0.1 | 0.0 | 0.5 |
| Bosnia and Herzegovina | Miscellaneous phytochemicals | Phytoprostanes | 0.1 | 0.0 | 0.6 |
| Croatia | Miscellaneous phytochemicals | Phytoprostanes | 0.1 | 0.0 | 0.7 |
| Cyprus | Miscellaneous phytochemicals | Phytoprostanes | 0.2 | 0.0 | 0.9 |
| Czechia | Miscellaneous phytochemicals | Phytoprostanes | 0.1 | 0.0 | 0.2 |
| Denmark | Miscellaneous phytochemicals | Phytoprostanes | 0.1 | 0.0 | 0.4 |
| Estonia | Miscellaneous phytochemicals | Phytoprostanes | 0.1 | 0.0 | 0.2 |
| Finland | Miscellaneous phytochemicals | Phytoprostanes | 0.2 | 0.0 | 0.4 |
| France | Miscellaneous phytochemicals | Phytoprostanes | 0.2 | 0.0 | 1.1 |
| Germany | Miscellaneous phytochemicals | Phytoprostanes | 0.1 | 0.0 | 0.1 |
| Greece | Miscellaneous phytochemicals | Phytoprostanes | 0.3 | 0.0 | 0.7 |
| Hungary | Miscellaneous phytochemicals | Phytoprostanes | 0.2 | 0.0 | 0.9 |
| Ireland | Miscellaneous phytochemicals | Phytoprostanes | 0.2 | 0.0 | 0.6 |
| Italy | Miscellaneous phytochemicals | Phytoprostanes | 0.2 | 0.0 | 0.7 |
| Latvia | Miscellaneous phytochemicals | Phytoprostanes | 0.1 | 0.0 | 0.4 |
| Montenegro | Miscellaneous phytochemicals | Phytoprostanes | 0.1 | 0.0 | 0.4 |
| Netherlands | Miscellaneous phytochemicals | Phytoprostanes | 0.1 | 0.0 | 0.5 |
| Poland | Miscellaneous phytochemicals | Phytoprostanes | 0.1 | 0.0 | 0.3 |
| Portugal | Miscellaneous phytochemicals | Phytoprostanes | 0.2 | 0.0 | 0.7 |
| Romania | Miscellaneous phytochemicals | Phytoprostanes | 0.1 | 0.0 | 0.4 |
| Serbia | Miscellaneous phytochemicals | Phytoprostanes | 0.2 | 0.0 | 0.5 |

| Country | Family | Class | Average intake (mg/day) | 5th percentile intake (mg/day) | 95th percentile intake (mg/day) |
|---|---|---|---|---|---|
| Slovenia | Miscellaneous phytochemicals | Phytoprostanes | 0.1 | 0.0 | 0.3 |
| Spain | Miscellaneous phytochemicals | Phytoprostanes | 0.1 | 0.0 | 0.3 |
| Sweden | Miscellaneous phytochemicals | Phytoprostanes | 0.0 | 0.0 | 0.1 |
| United Kingdom | Miscellaneous phytochemicals | Phytoprostanes | 0.1 | 0.0 | 0.6 |
| Austria | Miscellaneous phytochemicals | Thiosulfinates | 76.7 | 0.0 | 354.6 |
| Belgium | Miscellaneous phytochemicals | Thiosulfinates | 61.7 | 0.0 | 289.7 |
| Bosnia and Herzegovina | Miscellaneous phytochemicals | Thiosulfinates | 62.5 | 0.0 | 276.6 |
| Croatia | Miscellaneous phytochemicals | Thiosulfinates | 91.4 | 0.0 | 335.9 |
| Cyprus | Miscellaneous phytochemicals | Thiosulfinates | 89.4 | 0.0 | 399.3 |
| Czechia | Miscellaneous phytochemicals | Thiosulfinates | 120.7 | 0.0 | 484.9 |
| Denmark | Miscellaneous phytochemicals | Thiosulfinates | 31.1 | 0.0 | 94.9 |
| Estonia | Miscellaneous phytochemicals | Thiosulfinates | 63.5 | 0.0 | 258.3 |
| Finland | Miscellaneous phytochemicals | Thiosulfinates | 47.5 | 0.0 | 219.8 |
| France | Miscellaneous phytochemicals | Thiosulfinates | 120.6 | 0.0 | 508.6 |
| Germany | Miscellaneous phytochemicals | Thiosulfinates | 54.6 | 0.0 | 250.4 |
| Greece | Miscellaneous phytochemicals | Thiosulfinates | 74.6 | 0.0 | 304.4 |
| Hungary | Miscellaneous phytochemicals | Thiosulfinates | 98.5 | 0.0 | 322.7 |
| Ireland | Miscellaneous phytochemicals | Thiosulfinates | 70.7 | 0.0 | 338.8 |
| Italy | Miscellaneous phytochemicals | Thiosulfinates | 35.6 | 0.0 | 137.3 |
| Latvia | Miscellaneous phytochemicals | Thiosulfinates | 133.3 | 0.0 | 601.7 |
| Montenegro | Miscellaneous phytochemicals | Thiosulfinates | 89.4 | 0.0 | 360.8 |
| Netherlands | Miscellaneous phytochemicals | Thiosulfinates | 63.2 | 0.0 | 311.3 |
| Poland | Miscellaneous phytochemicals | Thiosulfinates | 56.4 | 0.0 | 276.2 |
| Portugal | Miscellaneous phytochemicals | Thiosulfinates | 164.2 | 0.0 | 469.3 |
| Romania | Miscellaneous phytochemicals | Thiosulfinates | 131.0 | 1.5 | 599.3 |

| Country | Family | Class | Average intake (mg/day) | 5th percentile intake (mg/day) | 95th percentile intake (mg/day) |
|---|---|---|---|---|---|
| Serbia | Miscellaneous phytochemicals | Thiosulfinates | 72.3 | 0.0 | 340.1 |
| Slovenia | Miscellaneous phytochemicals | Thiosulfinates | 145.2 | 0.0 | 846.6 |
| Spain | Miscellaneous phytochemicals | Thiosulfinates | 112.9 | 0.0 | 329.3 |
| Sweden | Miscellaneous phytochemicals | Thiosulfinates | 86.5 | 0.0 | 357.4 |
| United Kingdom | Miscellaneous phytochemicals | Thiosulfinates | 76.4 | 0.0 | 357.6 |
| Austria | Miscellaneous phytochemicals | Miscellaneous | 2.4 | 0.0 | 10.9 |
| Belgium | Miscellaneous phytochemicals | Miscellaneous | 2.5 | 0.0 | 10.1 |
| Bosnia and Herzegovina | Miscellaneous phytochemicals | Miscellaneous | 2.0 | 0.0 | 8.5 |
| Croatia | Miscellaneous phytochemicals | Miscellaneous | 1.1 | 0.0 | 5.6 |
| Cyprus | Miscellaneous phytochemicals | Miscellaneous | 1.7 | 0.0 | 8.2 |
| Czechia | Miscellaneous phytochemicals | Miscellaneous | 2.3 | 0.0 | 7.2 |
| Denmark | Miscellaneous phytochemicals | Miscellaneous | 3.3 | 0.0 | 13.2 |
| Estonia | Miscellaneous phytochemicals | Miscellaneous | 1.4 | 0.0 | 4.7 |
| Finland | Miscellaneous phytochemicals | Miscellaneous | 3.3 | 0.0 | 18.2 |
| France | Miscellaneous phytochemicals | Miscellaneous | 2.4 | 0.0 | 10.9 |
| Germany | Miscellaneous phytochemicals | Miscellaneous | 1.6 | 0.0 | 11.7 |
| Greece | Miscellaneous phytochemicals | Miscellaneous | 2.4 | 0.2 | 8.6 |
| Hungary | Miscellaneous phytochemicals | Miscellaneous | 1.0 | 0.0 | 3.5 |
| Ireland | Miscellaneous phytochemicals | Miscellaneous | 5.7 | 0.0 | 32.9 |
| Italy | Miscellaneous phytochemicals | Miscellaneous | 3.4 | 0.5 | 10.9 |
| Latvia | Miscellaneous phytochemicals | Miscellaneous | 3.3 | 0.0 | 11.7 |
| Montenegro | Miscellaneous phytochemicals | Miscellaneous | 1.4 | 0.0 | 7.8 |
| Netherlands | Miscellaneous phytochemicals | Miscellaneous | 1.9 | 0.0 | 11.1 |
| Poland | Miscellaneous phytochemicals | Miscellaneous | 1.1 | 0.0 | 5.4 |
| Portugal | Miscellaneous phytochemicals | Miscellaneous | 2.4 | 0.1 | 8.5 |

| Country | Family | Class | Average intake (mg/day) | 5th percentile intake (mg/day) | 95th percentile intake (mg/day) |
|---|---|---|---|---|---|
| Romania | Miscellaneous phytochemicals | Miscellaneous | 4.0 | 0.0 | 16.3 |
| Serbia | Miscellaneous phytochemicals | Miscellaneous | 1.7 | 0.0 | 8.8 |
| Slovenia | Miscellaneous phytochemicals | Miscellaneous | 1.8 | 0.0 | 8.7 |
| Spain | Miscellaneous phytochemicals | Miscellaneous | 1.7 | 0.0 | 4.7 |
| Sweden | Miscellaneous phytochemicals | Miscellaneous | 2.4 | 0.0 | 12.7 |
| United Kingdom | Miscellaneous phytochemicals | Miscellaneous | 4.9 | 0.0 | 31.4 |

**Table S6: intake distribution (mg/day) of caffeine in Europe.**

| | Intake of caffeine (mg/day) | | |
|---|---|---|---|
| Country | Average | 5th percentile | 95th percentile |
| Austria | 369 | 0 | 1263 |
| Belgium | 148 | 0 | 549 |
| Bosnia and Herzegovina | 487 | 0 | 2031 |
| Croatia | 233 | 0 | 1011 |
| Cyprus | 262 | 0 | 1416 |
| Czechia | 242 | 0 | 819 |
| Denmark | 307 | 0 | 888 |
| Estonia | 436 | 0 | 1272 |
| Finland | 305 | 0 | 1252 |
| France | 322 | 0 | 1459 |
| Germany | 617 | 0 | 1771 |
| Greece | 189 | 0 | 618 |
| Hungary | 322 | 0 | 1295 |
| Ireland | 112 | 0 | 445 |
| Italy | 256 | 0 | 1008 |
| Latvia | 157 | 0 | 555 |
| Montenegro | 550 | 0 | 2130 |
| Netherlands | 307 | 0 | 1352 |
| Poland | 110 | 0 | 357 |
| Portugal | 193 | 0 | 697 |
| Romania | 427 | 0 | 1321 |
| Serbia | 707 | 0 | 2375 |
| Slovenia | 140 | 0 | 421 |
| Spain | 51 | 0 | 251 |
| Sweden | 162 | 0 | 835 |
| United Kingdom | 126 | 0 | 443 |

| | Intake of caffeine (mg/day) | | |
|---|---|---|---|
| Country | Average | 5th percentile | 95th percentile |
| Europe | 290 | 0 | 1071 |

**Table S7: descriptive characteristics of European country surveys.**

| Country | Survey start year | Survey name | Population Group | n of individuals | Food intake assessment method used |
|---|---|---|---|---|---|
| Austria | 2014 | EU Menu Austria: Food consumption data for Austrian adults | Adults | 2169 | 24-hours dietary recall |
| Belgium | 2014 | Belgian national food consumption survey in children, adolescents and adults | Adults | 1234 | Food record, 24-hours dietary recall |
| Bosnia and Herzegovina | 2017 | Bosnia-Herzegovinian Dietary Survey of adolescents, adults and pregnant women | Adults | 850 | 24-hours dietary recall |
| Croatia | 2021 | Croatian national food consumption survey on adolescents and adults from 10 to 99 years of age | Adults | 869 | 24-hours dietary recall |
| Cyprus | 2014 | National dietary survey of the adult population of Cyprus | Adults | 272 | 24-hours dietary recall |
| Czechia | 2003 | Czech National Food Consumption Survey | Adults | 1666 | 24-hours dietary recall |
| Denmark | 2005 | The Danish National Dietary survey 2005-2008 | Adults | 1739 | Food record |
| Estonia | 2013 | National Dietary Survey among 11-74 years old individuals in Estonia | Adults | 2124 | 24-hours dietary recall |
| Finland | 2017 | FINDIET 2017 | Adults | 1196 | 24-hours dietary recall |
| France | 2014 | The French national dietary survey (INCA3, 2014-2015) | Adults | 1773 | Food record, 24-hours dietary recall |
| Germany | 2007 | National Nutrition Survey II | Adults | 10419 | 24-hours dietary recall |
| Greece | 2014 | The EFSA-funded collection of dietary and related data in the general population aged 10-74 years in Greece | Adults | 260 | 24-hours dietary recall |
| Hungary | 2018 | Hungarian national food consumption survey | Adults | 529 | Food record, 24-hours dietary recall |
| Ireland | 2008 | National Adult Nutrition Survey | Adults | 1274 | Food record |
| Italy | 2018 | Italian national dietary survey on adult population from 10 up to 74 years old IV SCAI ADULT 2018-2020 | Adults | 726 | 24-hours dietary recall |
| Latvia | 2012 | Latvian National Dietary survey | Adults | 1080 | Food record, 24-hours dietary recall |

| **Country** | **Survey start year** | **Survey name** | **Population Group** | **n of individuals** | **Food intake assessment method used** |
|---|---|---|---|---|---|
| Montenegro | 2017 | Montenegrin National Dietary Survey on the general population | Adults | 697 | 24-hours dietary recall |
| Netherlands | 2012 | Dutch National Food Consumption Survey 2012-2016 (DNFCS) | Adults | 1478 | Food record, 24-hours dietary recall |
| Poland | 2019 | National Dietary Survey on the adult population | Adults | 1195 | 24-hours dietary recall |
| Portugal | 2015 | National Food, Nutrition and Physical Activity Survey of the Portuguese general population | Adults | 3102 | Food record, 24-hours dietary recall |
| Romania | 2019 | Romanian national food consumption survey for adolescents, adults and elderly | Adults | 740 | 24-hours dietary recall |
| Serbia | 2019 | Serbian Food Consumption Survey on adults | Adults | 1150 | 24-hours dietary recall |
| Slovenia | 2017 | Slovenian national food consumption survey | Adults | 385 | Food record, 24-hours dietary recall |
| Spain | 2013 | Spanish National dietary survey in adults, elderly and pregnant women | Adults | 536 | 24-hours dietary recall |
| Sweden | 2016 | RIKSMATEN ADOLESCENTS 2016 | Adults | 215 | Web-based dietary record |
| United Kingdom | 2008 | National Diet and Nutrition Survey - Years 1-3 | Adults | 1266 | Food record |
